# Mathematical model of COVID-19 spread in Turkey and South Africa: Theory, methods and applications

**DOI:** 10.1101/2020.05.08.20095588

**Authors:** Abdon Atangana, Seda İğret Araz

## Abstract

A comprehensive study about the spread of COVID-19 cases in Turkey and South Africa has been presented in this paper. An exhaustive statistical analysis encompassing arithmetic, geometric, harmonic means, standard deviation, skewness, variance, Pearson and Spearman correlation was derived from the data collected from Turkey and South Africa within the period of 11 March 2020 to 3 May 2020 and 05 March and 3 of May respectively. It was observed that in the case of Turkey, a negative Spearman correlation for the number of infected class and a positive Spearman correlation for both the number of deaths and recoveries were obtained. This implied that the daily infections could decrease, while the daily deaths and number of recovered people could increase under current conditions. In the case of South Africa, a negative Spearman correlation for both daily deaths and daily infected people was obtained, indicating that these numbers may decrease if the current conditions are maintained. The utilization of a statistical technique predicted the daily number of infected, recovered and dead people for each country; and three results were obtained for Turkey, namely an upper boundary, a prediction from current situation and lower boundary. The prediction shows that Turkey may register in the near future approximately more than 6000 new infections in a day as worst case scenario; and less than 300 cases in the perfect scenario. However, the country could register in the near future a daily number of 27000 people recovered from COVID-19 in the perfect scenario; and less than 5000 people in a worst scenario. Moreover, Turkey in a worst-case scenario could record a high number of approximately 200 deaths per day; and less than 150 deaths in a perfect scenario. Similarly, in the case of South Africa, the prediction results show that in the near future the country could register about 500 new infected cases daily and more than 25 deaths in the worst scenario; while in a perfect scenario less than 50 new infected and zero death cases could be recorded. The histograms of the daily number of newly infected, recovered and death showed a sign of lognormal and normal distribution, which is presented using the Bell curving method parameters estimation. A new mathematical model COVID-19 comprised of nine classes was suggested; of which a formula of the reproductive number, well-poseness of the solutions and the stability analysis were presented in details. The suggested model was further extended to the scope of nonlocal operators for each case; whereby the Atangana-Seda numerical method was used to provide numerical solutions, and simulations were performed for different non-integer numbers. Additionally, sections devoted to control optimal and others dedicated to compare cases between Turkey and South Africa with the aim to comprehend why there are less numbers of deaths and infected people in South Africa than Turkey were presented in details.

## 1 Introduction

It is a Thursday morning, 5 March in South Africa, everybody is busy with his daily routine, when the National Institute for Communicable Diseases confirmed the first positive case of COVID-19. A situation that was known for other countries now has become true and real in South Africa. How did we get here? The outbreak of COVID-19 started in China, Wuhan City, around December 2019; but within a short period the spread crossed over to some countries in Europe like United Kingdom, Italy, Spain and France. The first confirmed patient was a 38-year old male who visited Italy and arrived back in South Africa on March 1, 2020. The patient, after noticing symptoms of fever, malaise, a sore throat, cough and headache consulted a private general practitioner on March 3. From the 5 March to 15 March 2020 the number of infected people increased significantly, as a result on 15 March 2020, a national state of disaster was declared by the President of South Africa to mitigate the spread of COVID-19. This announcement was followed by measures including immediate travel restrictions and closure of schools from 18 March 2020. On 23 March, the South African government announced a country lockdown with effect on 26 March 2020. By the end of April, South Africa officially had 5647 confirmed cases. To date, South Africa is officially confirmed an African country with more confirmed cases, with 3471 active confirmed cases, 2073 recovered and 103 deaths due to COVID-19. In the case of Turkey, the first case of COVID-19 was confirmed and recorded on the 11 March 2020. Four days later, Turkey registered its first death caused by COVID-19. COVID-19 spread like wildfire in Turkey; in which by the 21 st of April the country had confirmed approximately 95591 cases of infected people, with 14918 number of recovered people and 2259 deaths recorded. The rapid spreading of COVID-19, has raised the total number of confirmed cases to 120200, of which 48900 have recovered and 3200 have died by the end of April 2020. In comparison to other European countries such as Iran, it is recorded that the total number of confirmed cases in Turkey surpassed it exceedingly; resulting in Turkey to be categorized as the most affected country in terms of numbers of confirmed cases within the settlement of the Middle East. Furthermore, Turkey’s total number of confirmed cases by the 20 April was also recorded to exceed that of China; even though there were some raised concerns that the total confirmed cases in China could have been underestimated. The consideration of these statistics prompted researchers from Turkey and South Africa to undertake research in different fields of science, technology and engineering in the last 3 months, since their future is left uncertain. As the virologists are focusing their attention in developing a vaccine that could be used to prevent the spread of the deadly virus; mathematicians rely on modelling techniques to produce multi-scenarios models that could be utilized to foresee the future [1-6]. Therefore, as mathematicians our role is to use and apply mathematical tools, particularly mathematical models, on suggested scenarios that could be helpful in predicting the future. In this paper, we present a detailed analysis of spread in both countries and structured the paper as follows: Section 2 presents a detailed statistical analysis of COVID-19 spread in Turkey. Then we present a detailed statistical analysis of COVID-19 spread in South Africa. Also after using the inverse problem approach and the Bell curving approach we present the parameter’s estimation, we present a comparative analysis between Turkey and South Africa. In Section 3, we suggest a new mathematical model of COVID-19 that takes into account nine classes, including, susceptible, infected with 5 sub-classes, recovered class, death and vaccinated. Then presents the positivity of solutions of the model as well as the reproduction number; and also deals with local and global asymptotic stability of disease free equilibrium and endemic points. In Section 4 we present an analysis of the suggested model with non-local operators. In section 5, we present numerical the suggested mathematical model for COVID-19 using Atangana-Seda scheme for fractional and fractal-fractional operators. In section 6, we present the optimal control of the disease. Finally we present a discussion, recommendations and conclusion respectively.

### 2 Statistical analysis of COVID-19 spread in Turkey and South Africa

To understand the impact of COVID-19, collection of numbers of daily new infected, recovered and deaths are performed all over the globe, such process follows a discrete approach. Thus, to understand and predict the impact of the Covid-19 on humans, statistics is associated with such collection, analysis, interpretation, organization and presentation. We shall recall that, this mathematical branch is wider applicable in numerous academic fields for example, natural and social science, business and government. Some important and useful statistical formula are means, variance, skewness, correlation, linear regression, Pearson’s correlation coeffcient, Spearman’s rank correlation test and many order. In this section, we present some formulas that will be used in this work for interpretation and prediction purposes. We define a data set whose values can be chosen as *x*_1_*,x*_2_*.…,x_n_*. We start with the arithmetic mean, 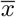, which states the mean of the *x*_1_*,x*_2_*.…,x_n_*. The arithmetic mean can be computed as

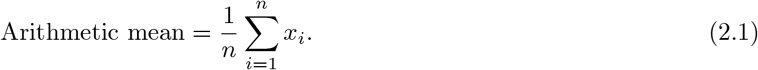

The formula of the geometric mean is

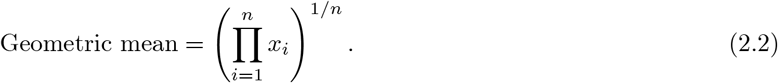

The formula of the harmonic mean is

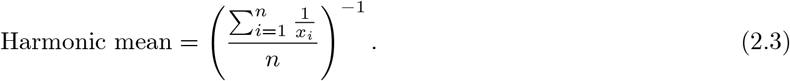

The formula of the standard deviation is

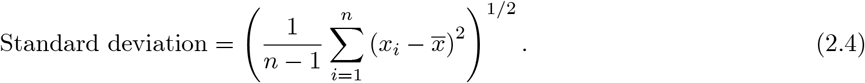

The formula of the skewness is

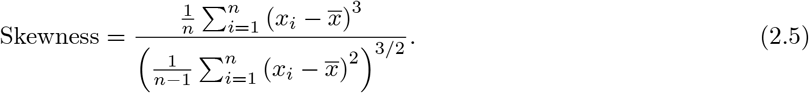

The formula of the variance is

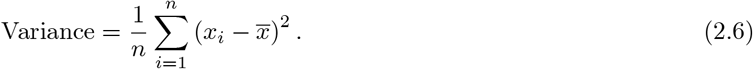

The formula of the covariance is

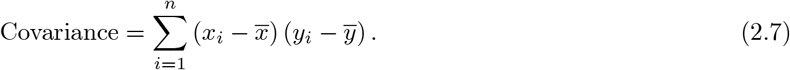

The formula of the Pearson correlation is

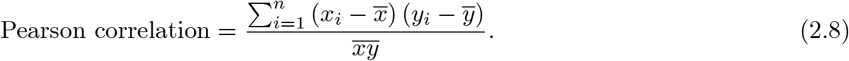

The formula of the Spearman correlation is

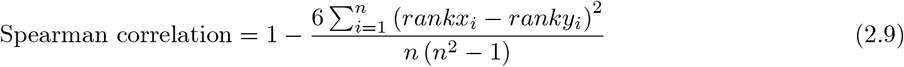

where rank enables to compared a numeric value with other values in the same list.

### 2.1 Statistical analysis for Turkey

In this section, we aim to provide a detailed statistical analysis of the collected data from Turkey. These data include, daily number of new infected, daily numbers of deaths, daily numbers of recovered and finally daily numbers of tested individuals. The collected data are from 11 March 2020 to 3 May 2020. The main aim of this section is to predict what could possibly happen in the near future using the reliability level method, additionally, to find which distribution each class follows. With the collected data, we will first present histogram, pie chart and nonlinear graphs for each class. The histograms will help identify the density of probability associated to each set of collected data. Additionally, we provide a polynomial fitting against collected. The results are presented in Figure 1 to 16. For each case, we present arithmetic, geometric, harmonic means respectively, skewness, variance, covariance, Pearson correlation and Spearman correlation and their results are presented in Table 1.

**Figure 1.**
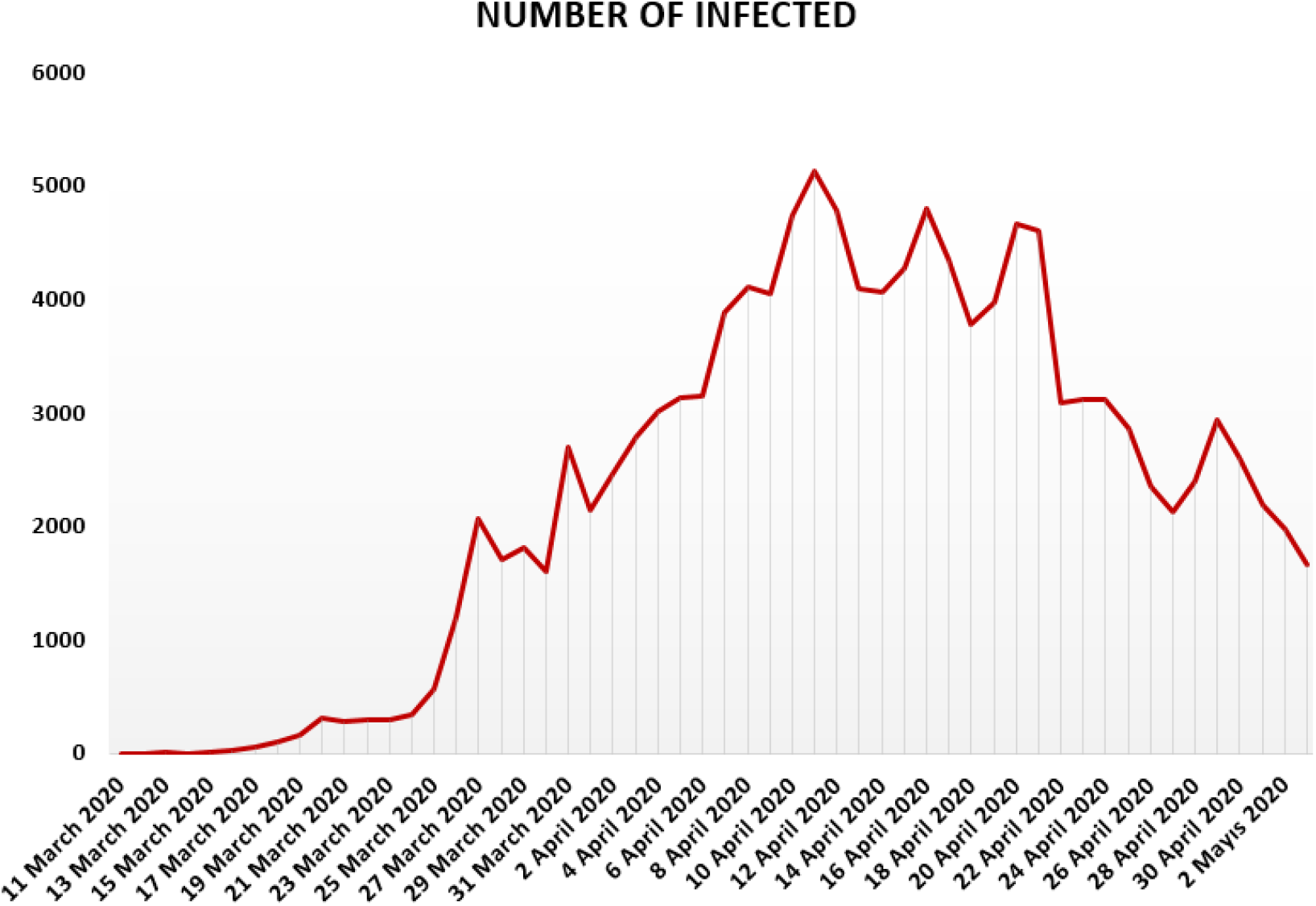
Number of infected people in Turkey from 11 March 2020 to 3 May 2020.

**Figure 2.**
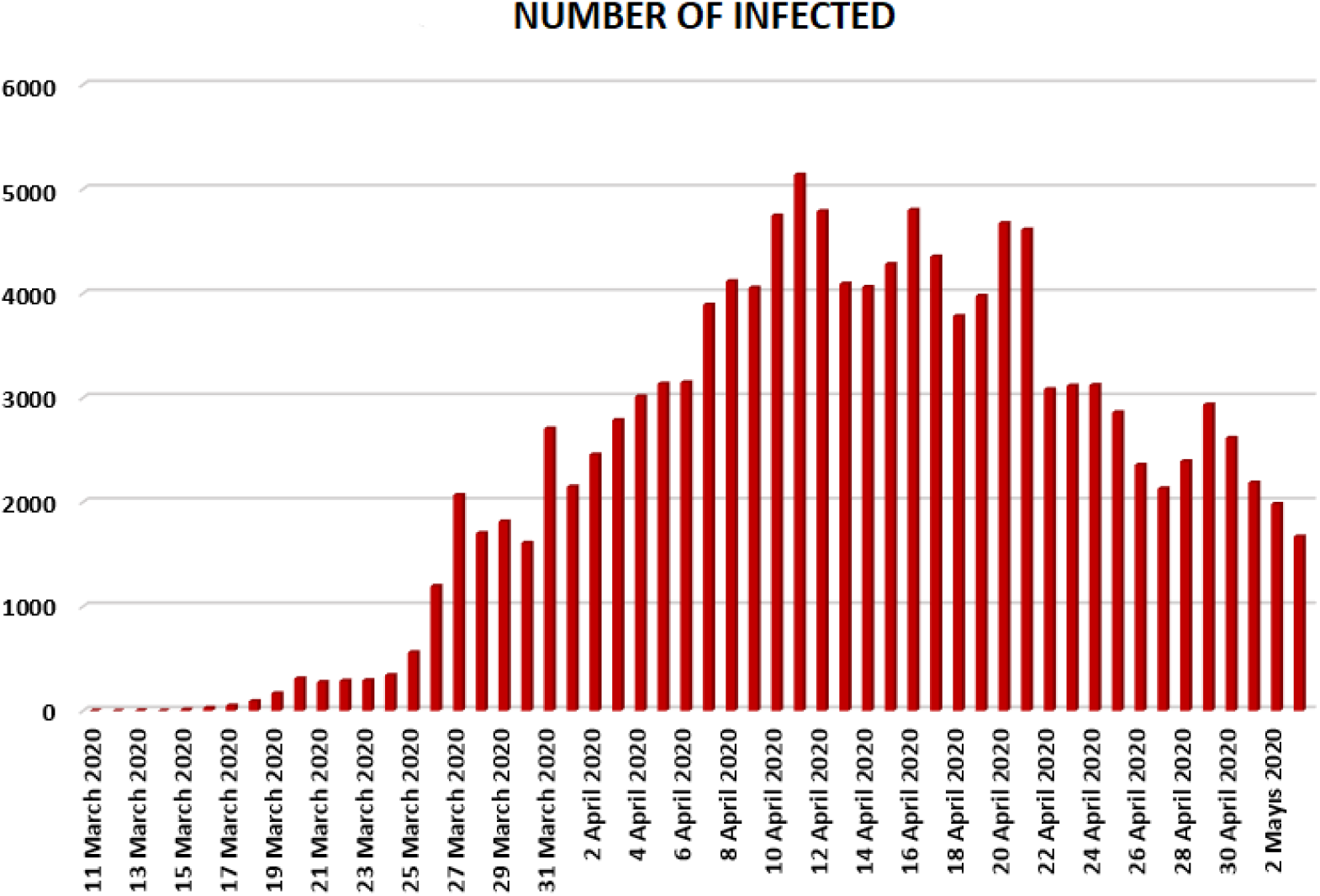
Number of infected people in Turkey from 11 March 2020 to 3 May 2020.

**Figure 3.**
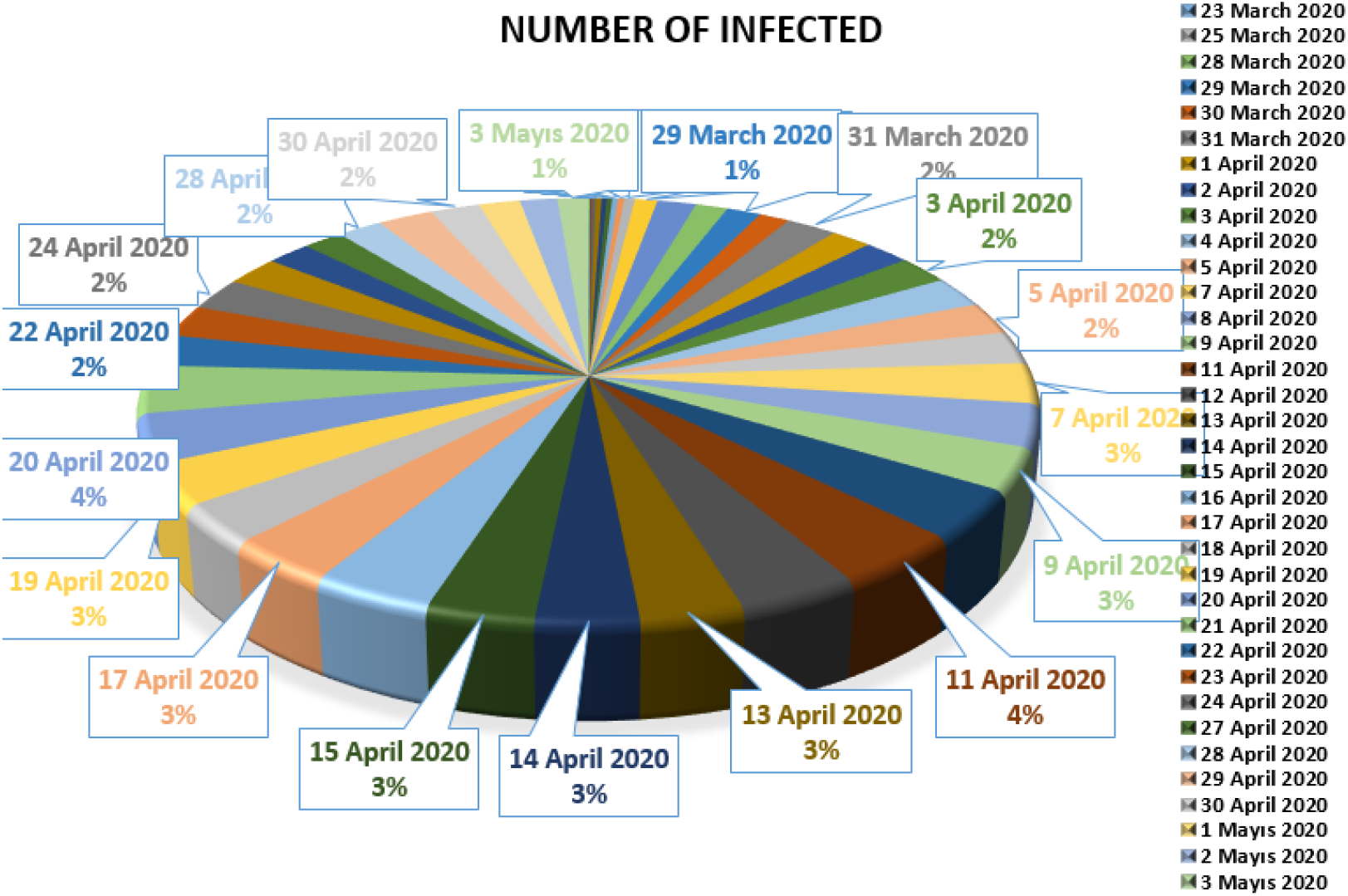
Number of infected people in Turkey from 11 March 2020 to 3 May 2020.

**Figure 4.**
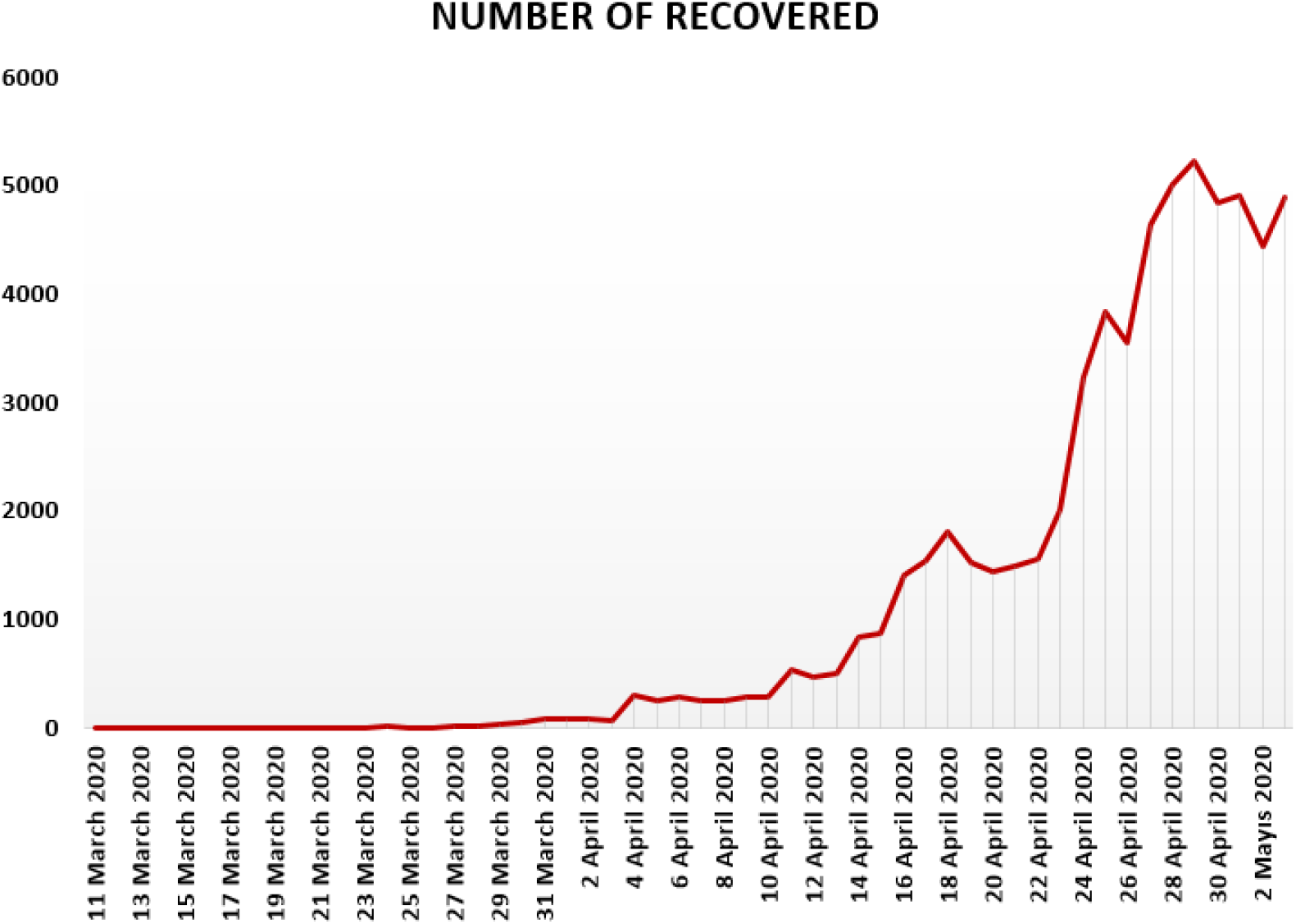
Number of recovered people in Turkey from 11 March 2020 to 3 May 2020.

**Figure 5.**
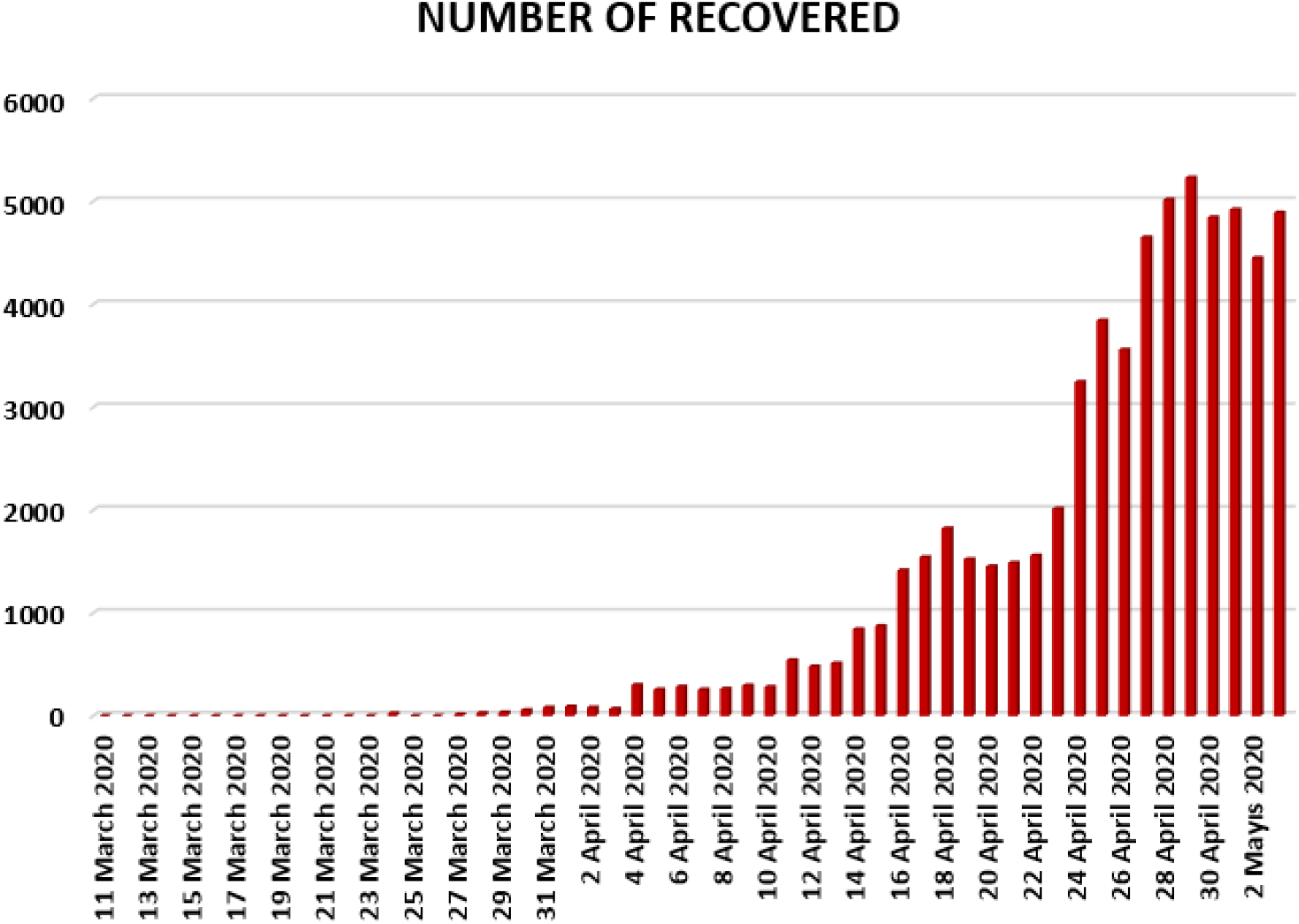
Number of recovered people in Turkey from 11 March 2020 to 3 May 2020.

**Figure 6.**
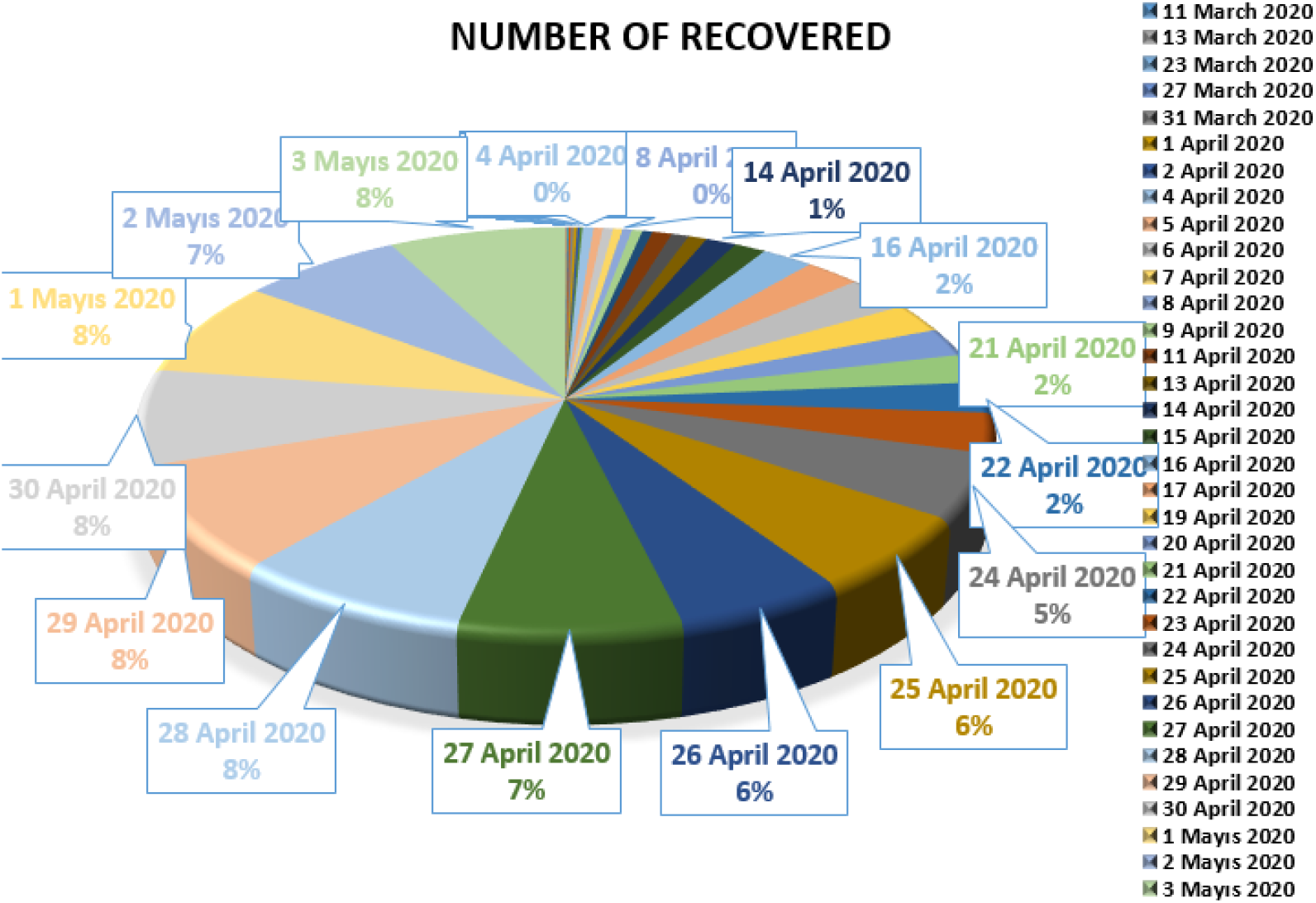
Number of recovered people in Turkey from 11 March 2020 to 3 May 2020.

In Figure 7, 8 and 9, we present some statistical simulation about number of died people due to COVID-19 in Turkey from 11 March 2020 to 3 May 2020.

**Figure 7.**
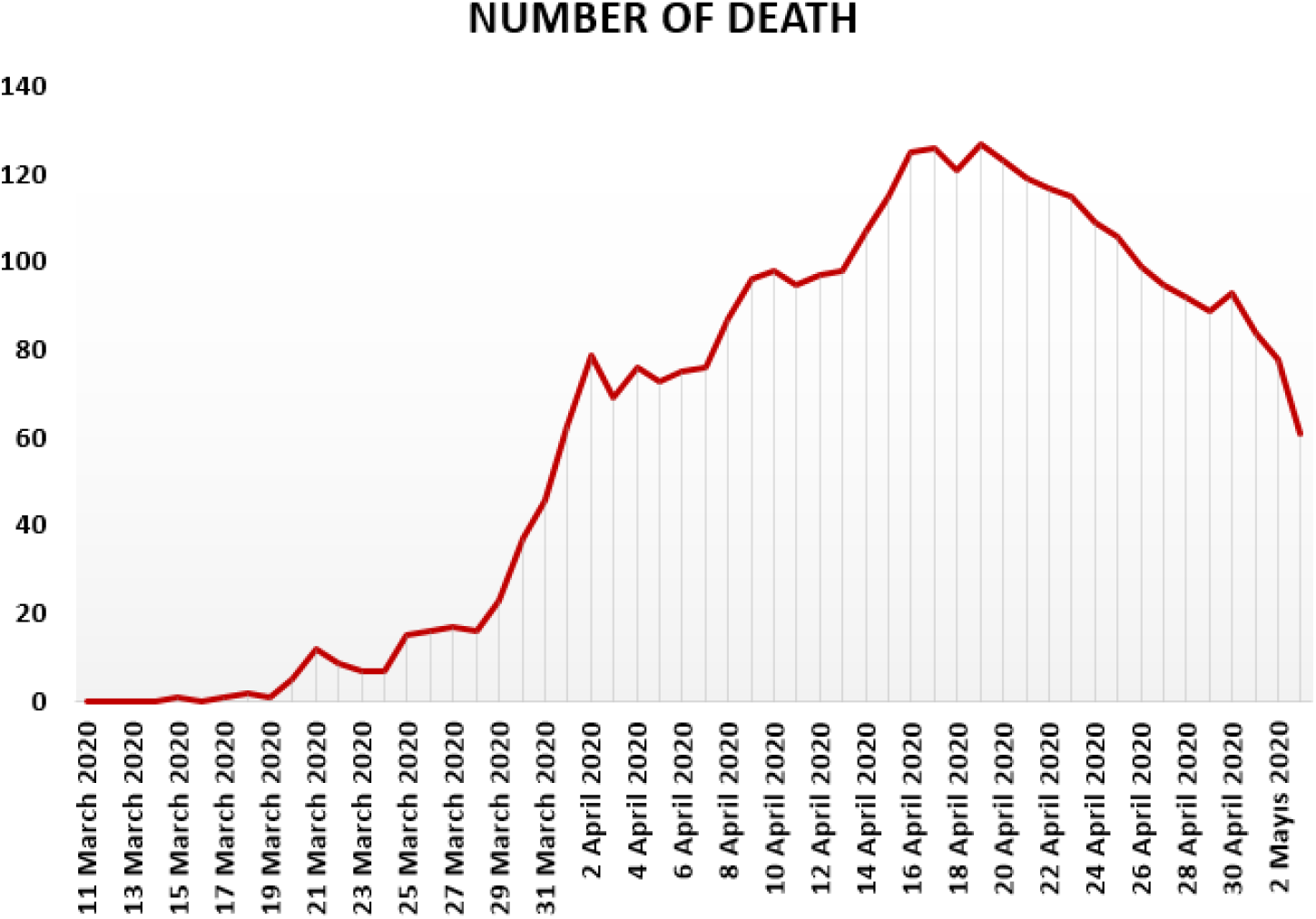
Number of death people in Turkey from 11 March 2020 to 3 May 2020.

**Figure 8.**
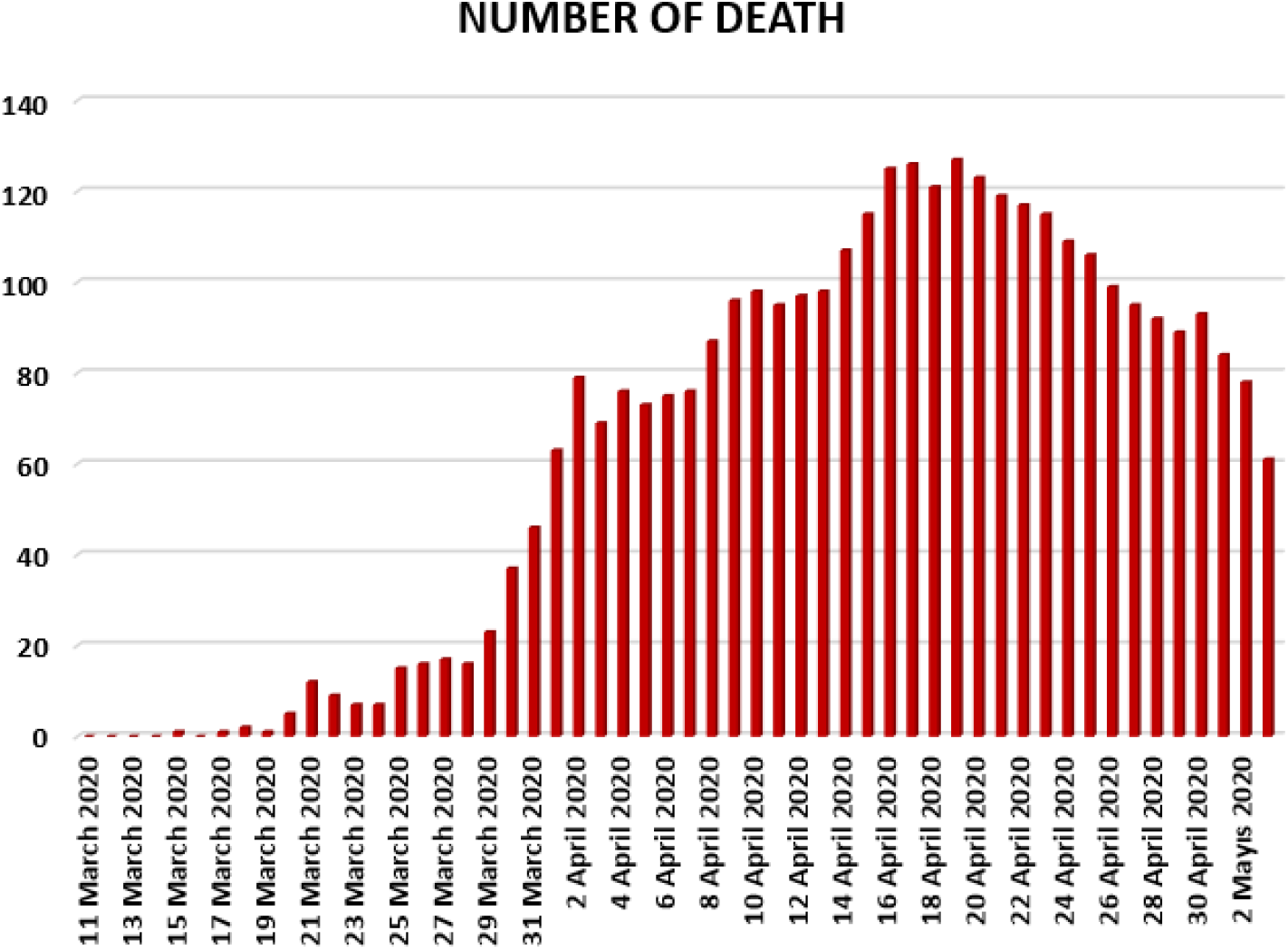
Number of death people in Turkey from 11 March 2020 to 3 May 2020.

**Figure 9.**
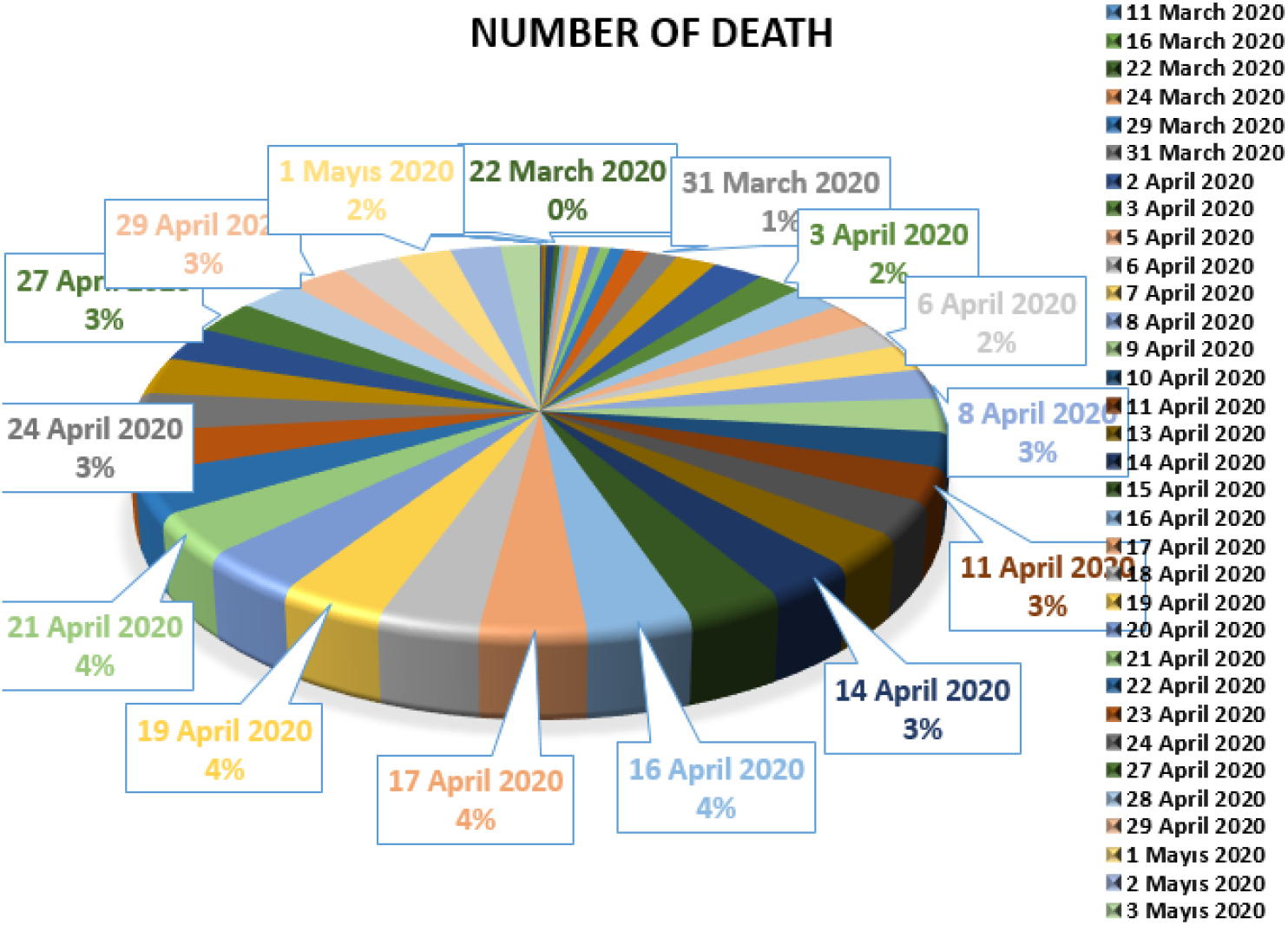
Number of death people in Turkey from 11 March 2020 to 3 May 2020.

In Figure 10, 11 and 12, we present some statistical simulation about number of tested people due to COVID-19 in Turkey from 11 March 2020 to 3 May 2020.

**Figure 10.**
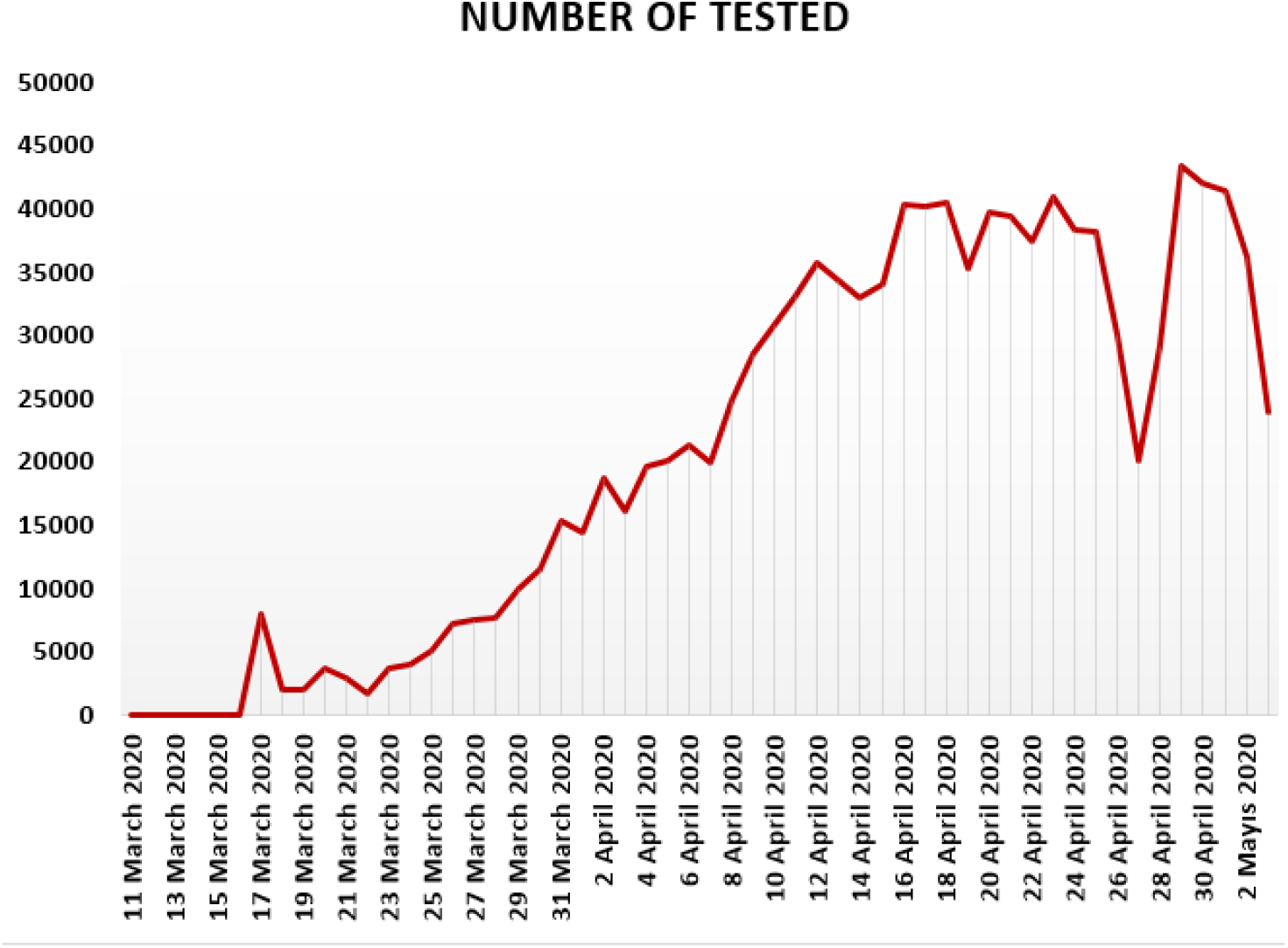
Number of tested people in Turkey from 11 March 2020 to 3 May 2020.

**Figure 11.**
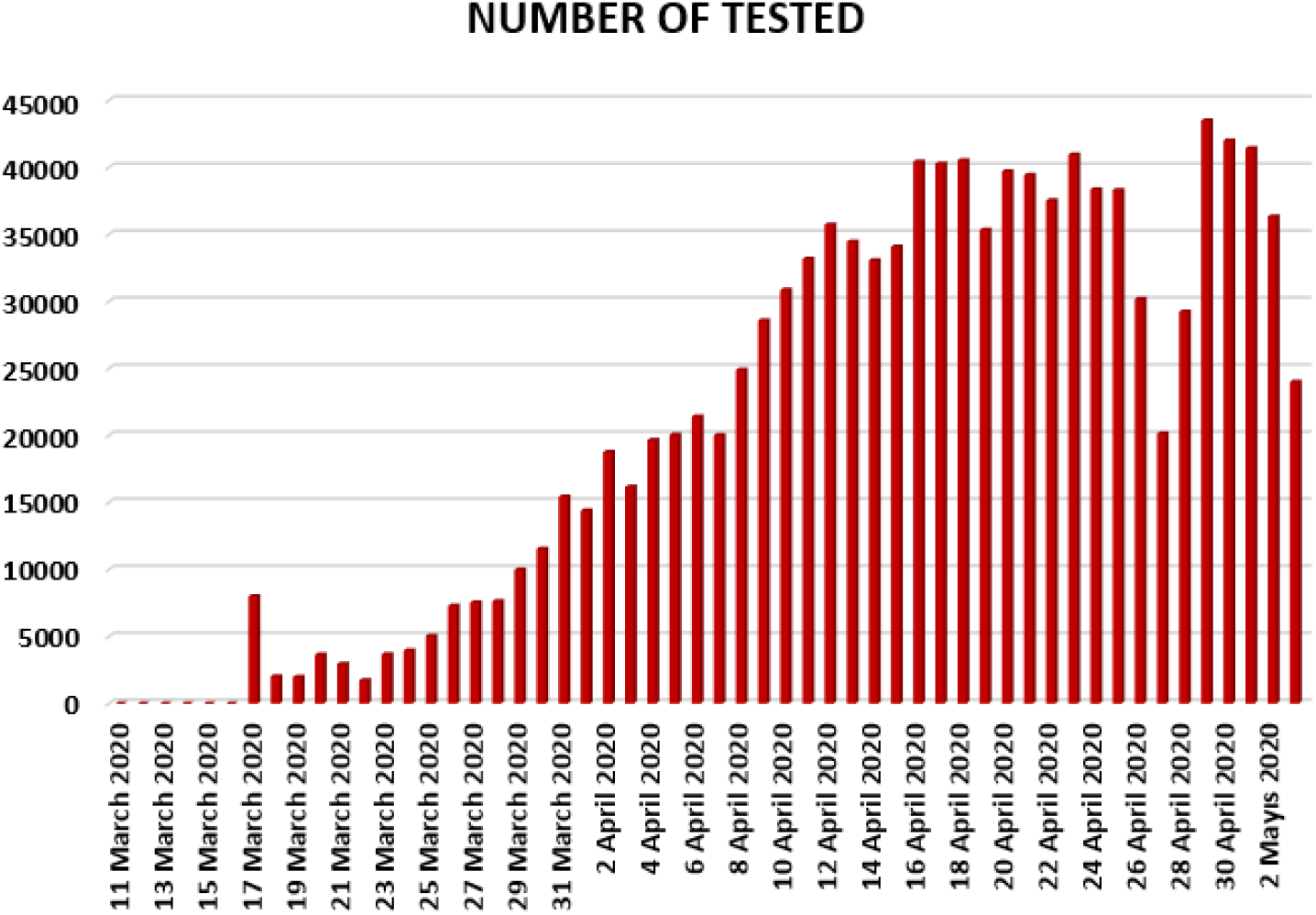
Number of tested people in Turkey from 11 March 2020 to 3 May 2020.

**Figure 12.**
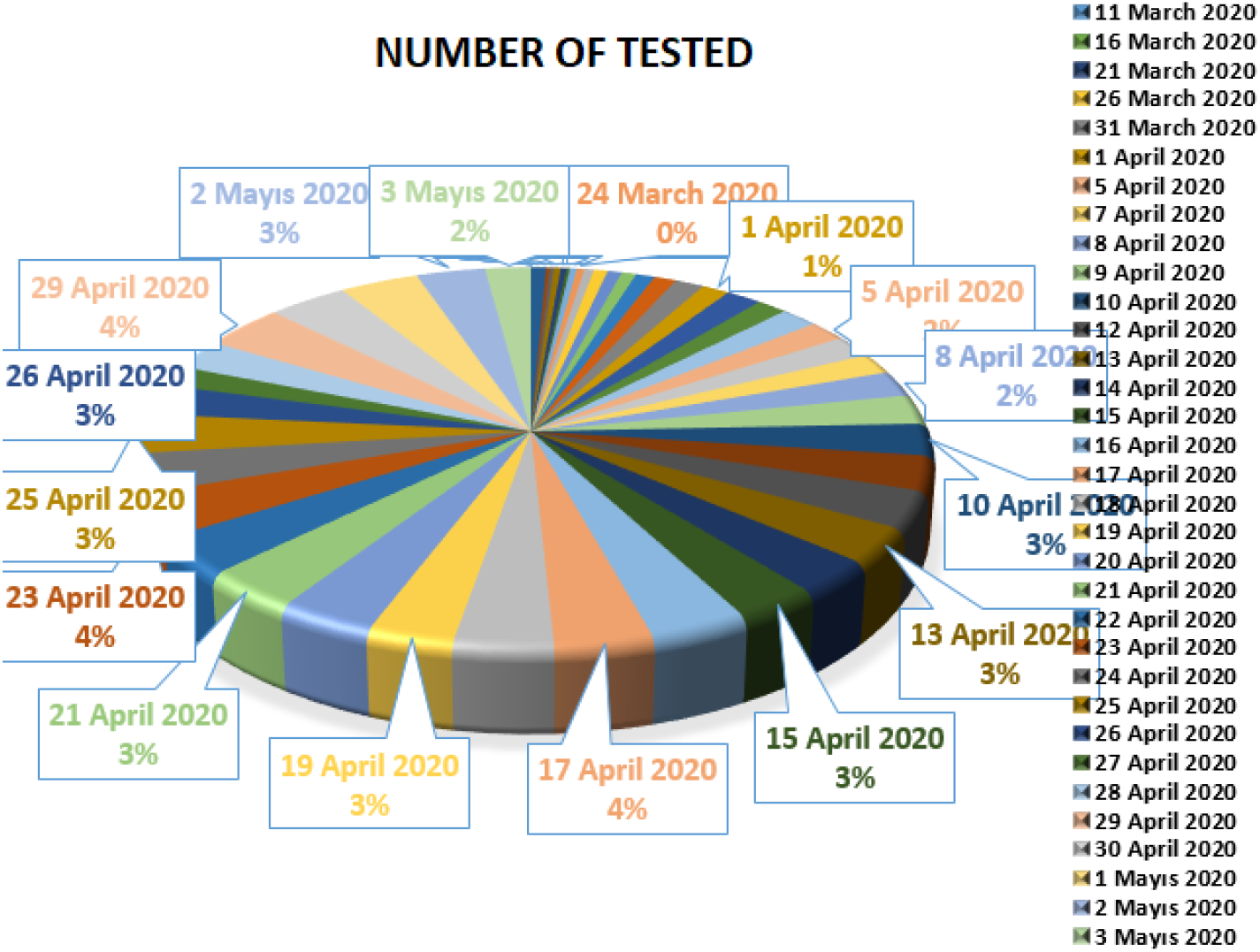
Number of tested people in Turkey from 11 March 2020 to 3 May 2020.

**Figure 13.**
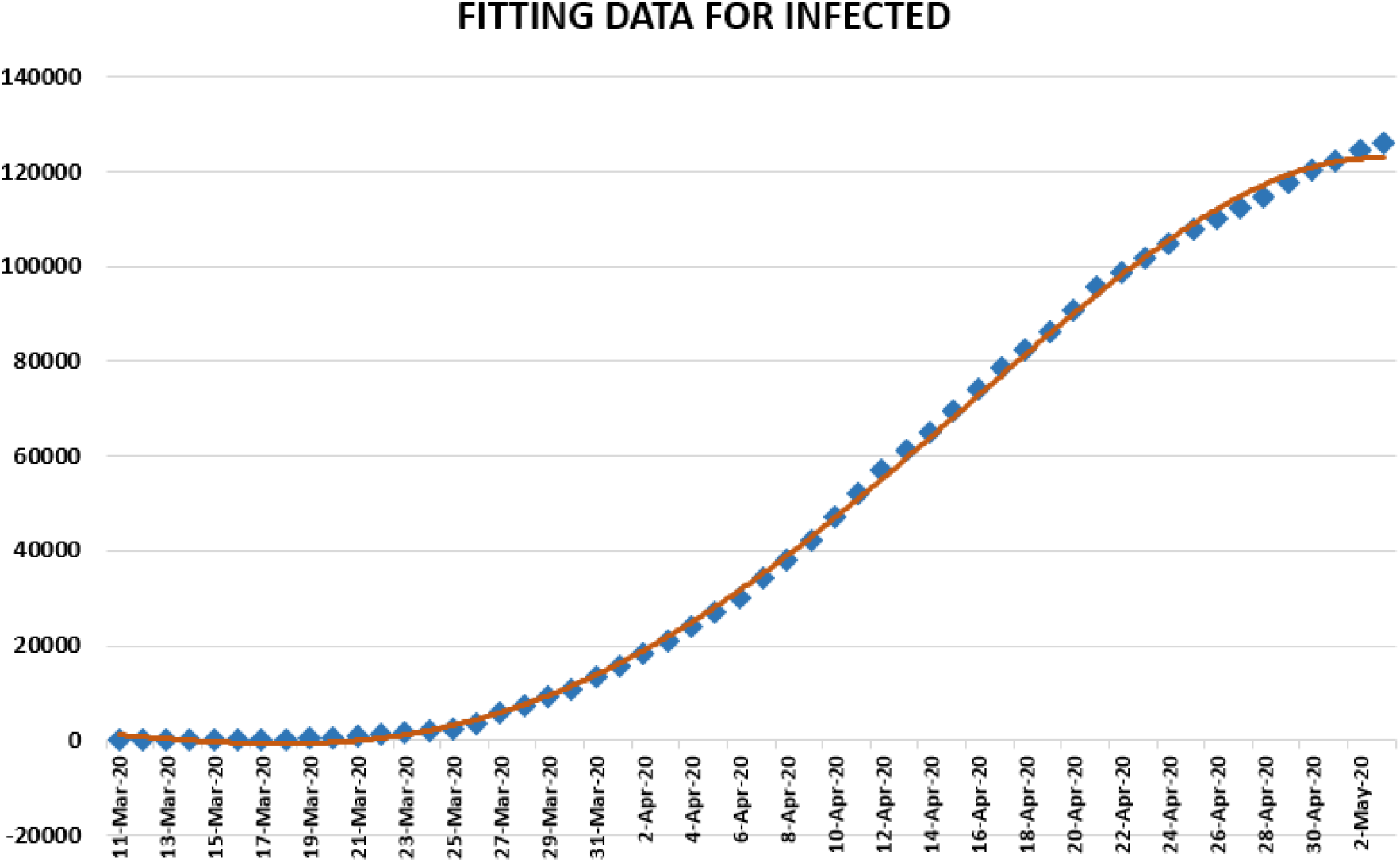
Polynomial fitting data for infected people in Turkey from 11 March 2020 to 3 May 2020.

**Figure 14.**
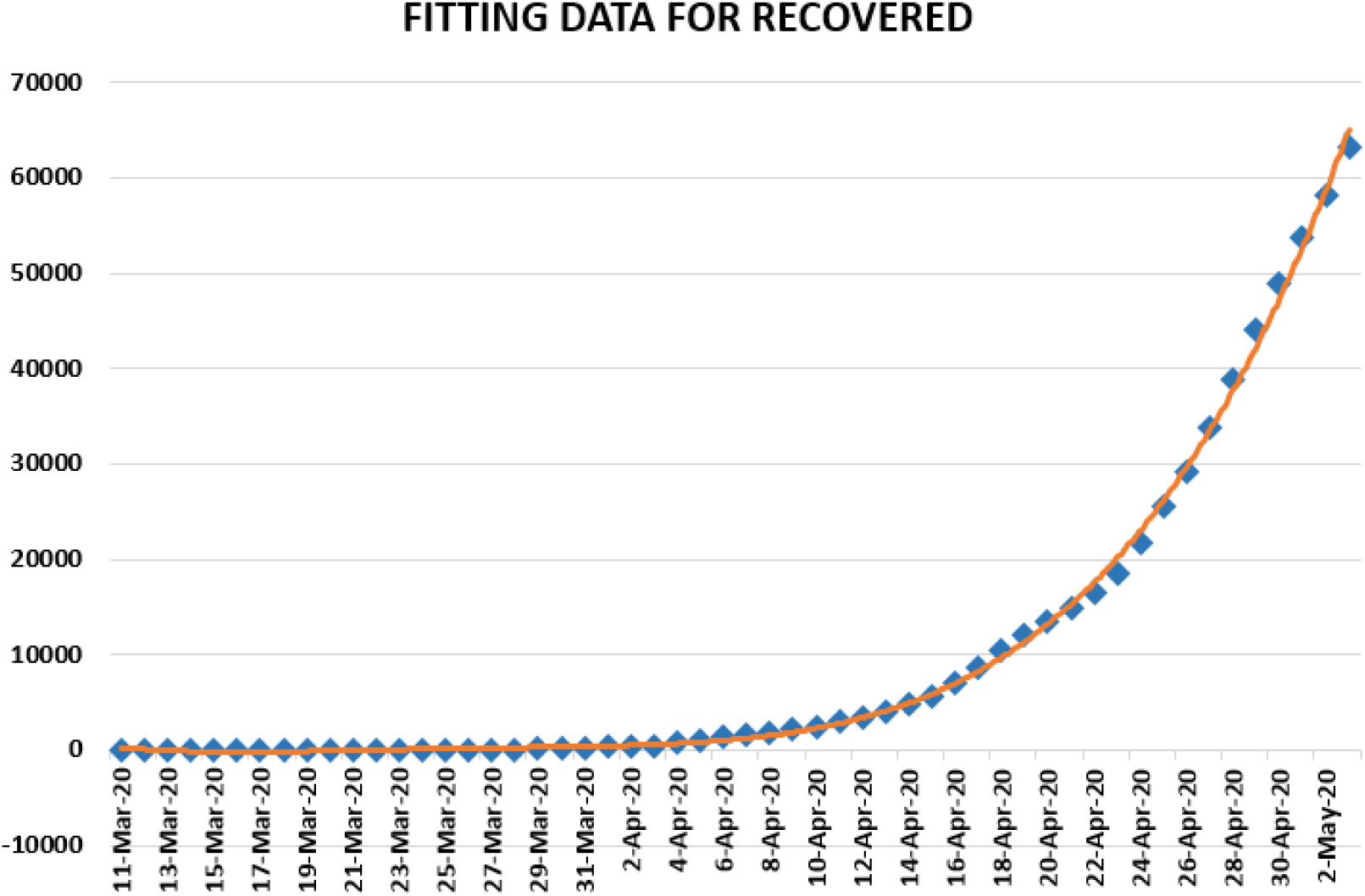
Polynomial fitting data for recovered people in Turkey from 11 March 2020 to 3 May 2020.

**Figure 15.**
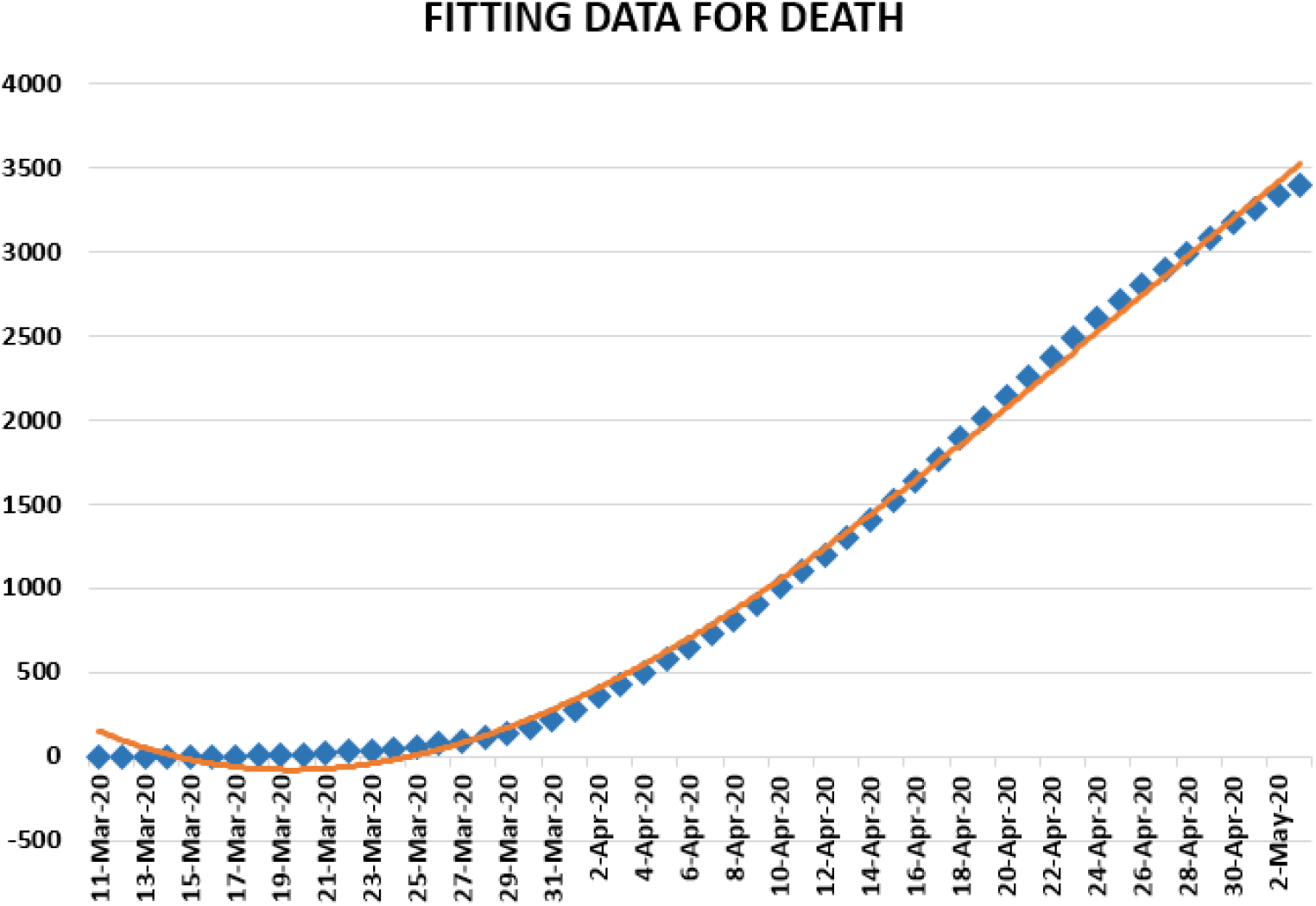
Polynomial fitting data for died people in Turkey from 11 March 2020 to 3 May 2020.

**Figure 16.**
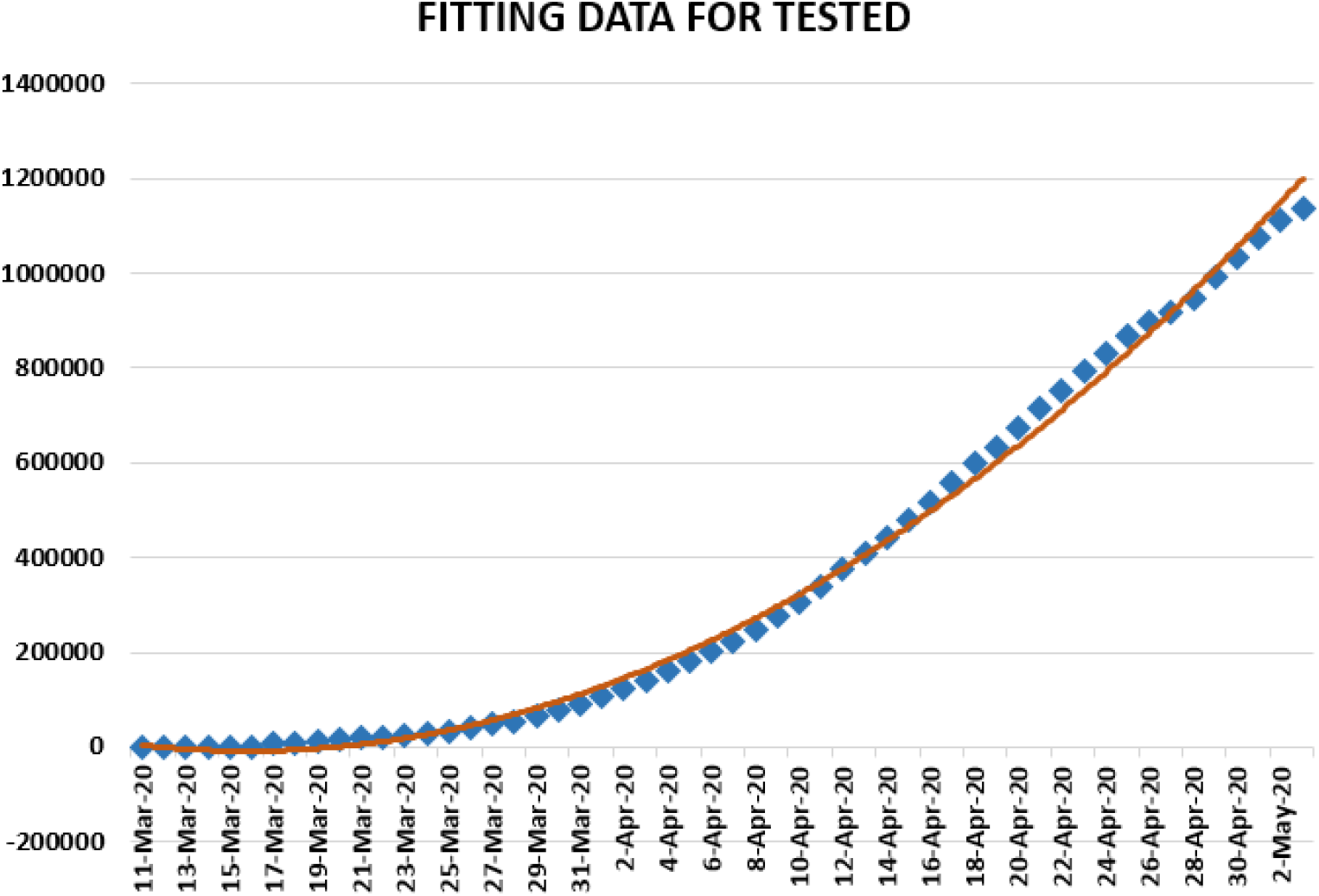
Polynomial fitting data for tested people in Turkey from 11 March 2020 to 3 May 2020. We present some statistical data about corona in Turkey in Table 1.

In Figure 1, 2 and 3, we present some statistical simulation about number of infected people due to COVID-19 in Turkey from 11 March 2020 to 3 May 2020.

#### 2.1.1 Regression analysis

Regression analysis which is also used in epidemiologic research enables us to examine amongst a set of variables. Here aim is to estimate outcomes benefitting from this set of variables. To do this, we find a prediction model in which we obtain model that fits best to the considered data and explains the response variable. We can utilize all possible independent variables, interactions, transformations with these models. To evaluate goodness of fit for the obtained model, we can utilize *R square* ability which is one of the different techniques used in regression diagnostics.

Linear regression models are given by

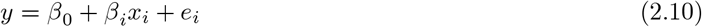

where *β*_0_, *β_i_* are the unknown constants, *x_i_* are the independent variables, *y* is the dependent variable and *e_i_* are the error terms in given data. If the value of *R square* is close to zero, this means that the significance of fit for model is unsuitable to predict outcomes. In other words, the obtained model used is not suitable for the given data and the obtained model should be left aside and another model should be found.

If the value of *R square* is close to one, this means that the significance of fit for model is suitable to predict outcomes. In this case, it can be passed to the other step of control analysis.

We firstly present a predictive analysis for infected people. According to the results obtained, we obtain a linear regression which is calculated as

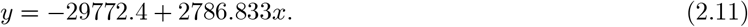

*F – test* was calculated as 1.94 × 10^−32^. *R square* was calculated as 0.93445. We can conclude from these data that the significance of fit for the obtained model is suitable for the considered data. Also we present polynomial regression which is calculated as

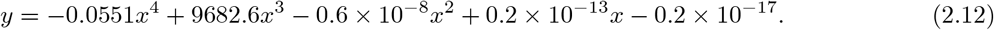

For this polynomial, *R square* was calculated as 0.9993. We present polynomial fitting data for infected people from 11 March to 3 May 2020.

We present a predictive analysis for recovered people. According to the results obtained, we get a linear regression which can be calculated as

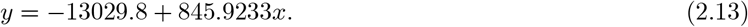

*F – test* was calculated as 1.39 × 10^−12^. *R square* was calculated as 0.622381. We can say that the significance of fit for model is not enough suitable for the considered data. To overcome this case, we can suggest another regression model

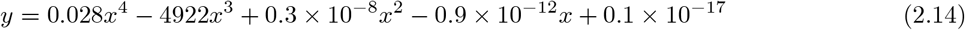

which is polynomial. For this polynomial, *R square* was calculated as 0.9987. We present a simulation about polynomial fitting data for recovered people from 11 March to 3 May 2020.

We present a predictive analysis for died people. According to the results obtained, we get a linear regression which can be calculated as

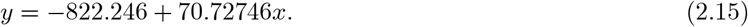

*F – test* was calculated as 1.75 × 10^−28^. *R square* was calculated as 0.907007. We can say that the significance of fit for model is suitable for the considered data. Also, we can present regression model

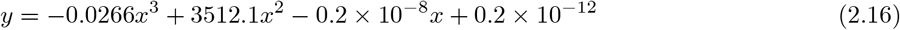

which is polynomial of third order. For this polynomial, *R square* was calculated as 0.9971. We present a simulation about polynomial fitting data for died people from 11 March to 3 May 2020.

We firstly present a predictive analysis for tested people. According to the results obtained, we get a linear regression which can be calculated as

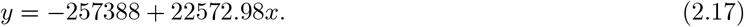

*F – test* was calculated as 5.17 × 10^−28^. *R square* was calculated as 0.903051. We can say that the significance of fit for model is suitable for the considered data. Also, we give polynomial regression model

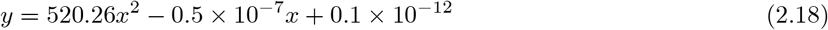

which is polynomial of second order. For this polynomial, *R square* was calculated as 0.9962. We present a simulation about polynomial fitting data for tested people from 11 March to 3 May 2020.

**Table 1.** Some data about corona in Turkey.

|  | **Infected** | **Recovered** | **Death** |
| --- | --- | --- | --- |
| **Arithmetic mean** | 2334.166667 | 1169.462963 | 56.63636364 |
| **Geometric mean** | 1029.876948 | 592.2822821 | 41.57620243 |
| **Harmonic mean** | 21.75203938 | 142.4039454 | 8.182426471 |
| **Standard deviation** | 1641.461139 | 1701.397957 | 48.05766445 |
| **Skewness** | -0.07519891 | 1.416616957 | 0.100241163 |
| **Variance** | 2644498.472 | 2841148.434 | 2045.624143 |
| **Covariance** | 17324.10185 | 21927.12037 | 607.7037037 |
| **Pearson Correlation** | 0.683518717 | 0.834653045 | 0.862085432 |
| **Spearman Correlation** | -17.9276729 | 0.188679245 | -8.39622641 |

Table 2 presents the covariance, the Pearson and Spearman correlation coefficients between daily cases-recovered, recovered-death and infected-death about COVID-19 in Turkey.

**Table 2.** Some data corona in Turkey.

|  | **Infected-Recovered** | **Recovered-Death** | **Infected-Death** |
| --- | --- | --- | --- |
| **Covariance** | 622993,071 | 38856,60837 | 66660,3642 |
| **Pearson Correlation** | 0,22728176 | 0,509688703 | 0,90632320 |
| **Spearman Correlation** | 0,64139508 | 0,817762531 | 0,88601105 |

We now present lognormal distribution for all cases in Turkey from 11 March to 03 May 2020.

**Figure 17.**
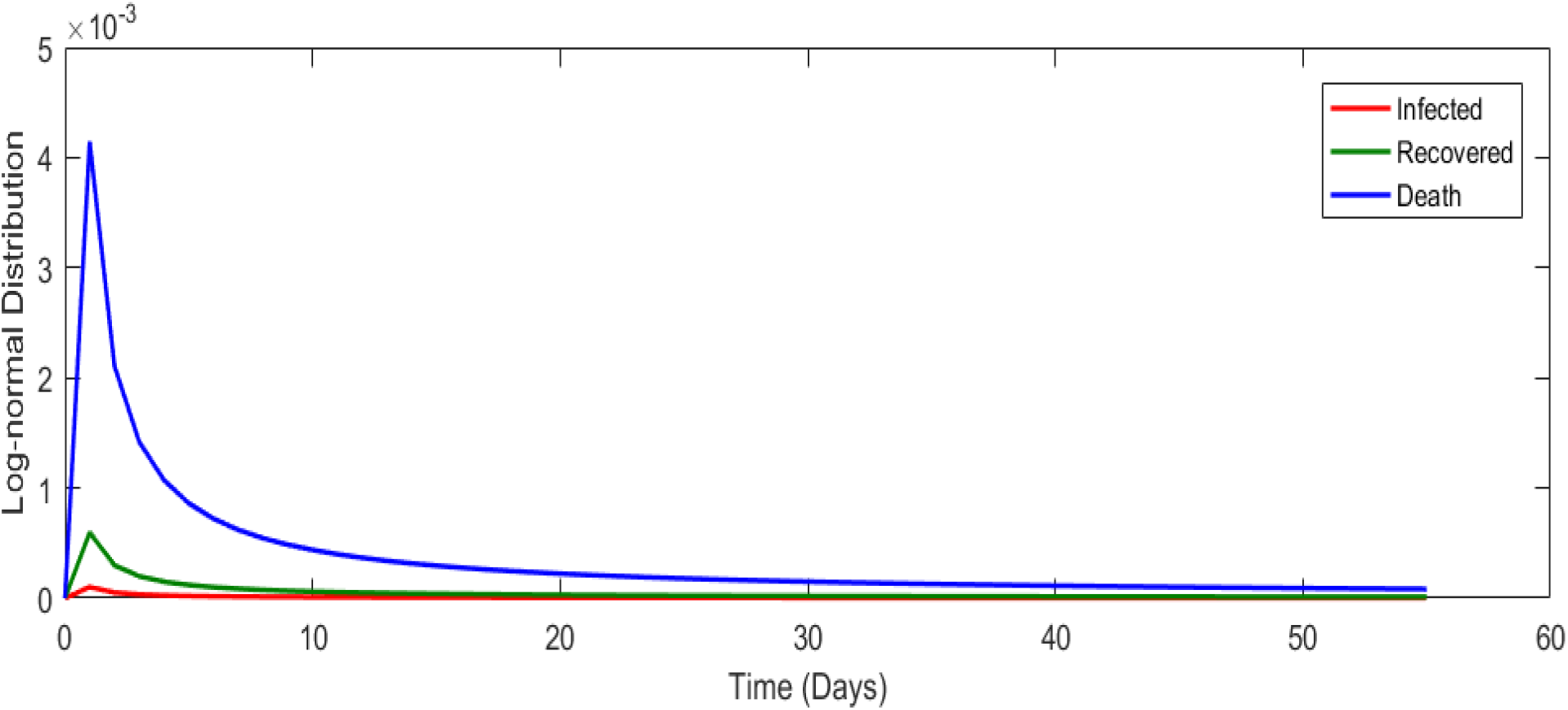
Lognormal distribution for all cases in Turkey from 11 March 2020 to 3 May 2020.

#### 2.1.2 Prediction about corona data in Turkey

In this section, we aim at performing prediction using existing data and reliability level method. The collected data will be considered from 11 March 2020 to 3 May 2020. The future prediction will start from 3 May 2020 to 15 of June 2020. This will help us give a prediction on numbers of new daily infected class, recovered, daily numbers of deaths in Turkey within this period. The prediction will consist of three different graphs comprising upper boundaries, middle lines and low boundaries. The upper boundaries represent the worse cases scenario, of course a scenario that is not needed for deaths class and infected but an ideal one for recovered class, and the lower boundaries representing perfect scenarios (A scenario that is needed) for Turkey to get rid of the infection. These results of prediction for future daily new infected, recovered and deaths are represented graphically in Figure 18, 19 and 20 respectively.

**Figure 18.**
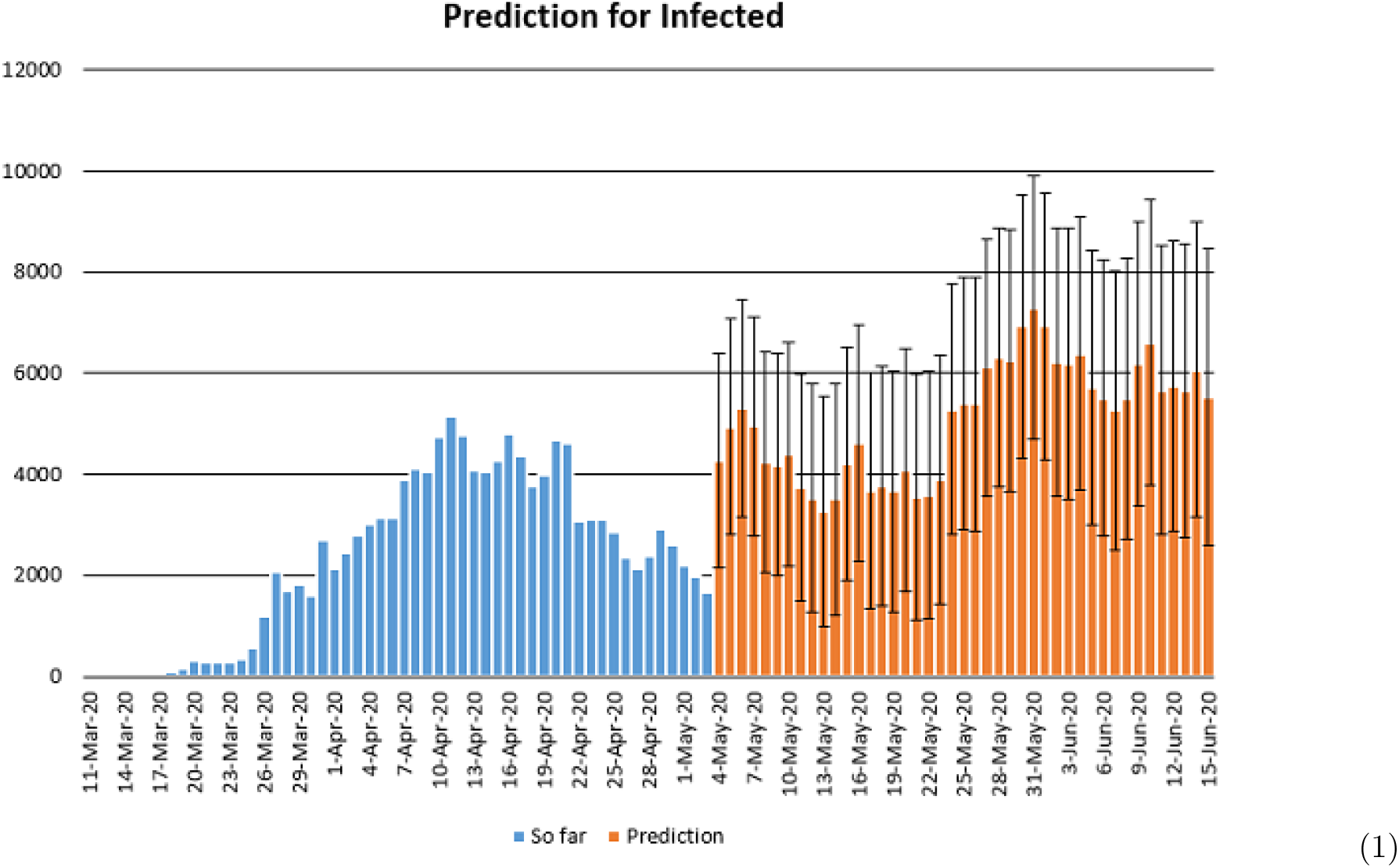
Prediction of daily number of infected in Turkey using Forecast Sheet with reliabilty level %97.

**Figure 19.**
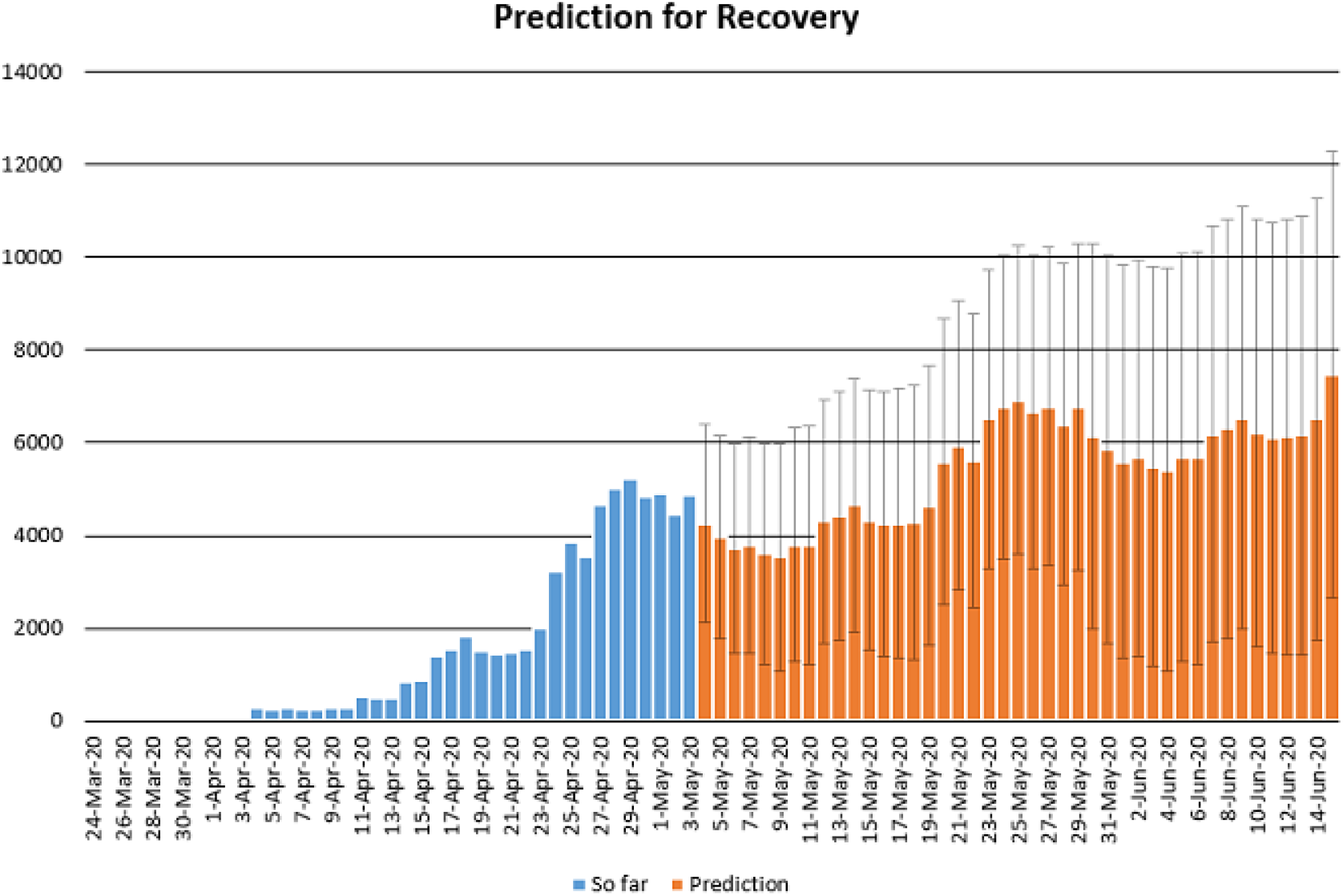
Prediction of daily number of recovered in Turkey using Forecast Sheet with reliabilty level %97.

**Figure 20.**
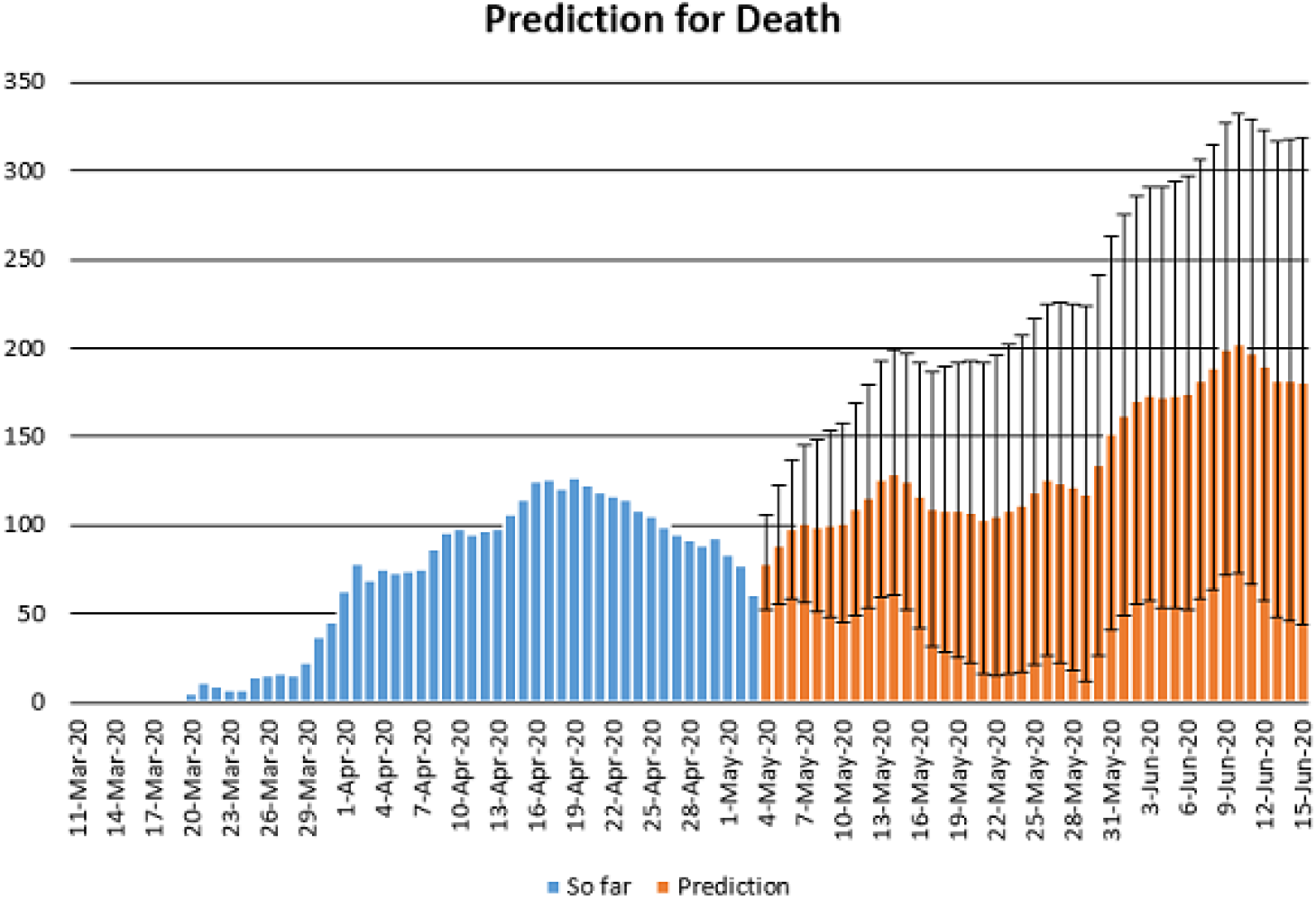
Prediction of daily number of deaths in Turkey using Forecast Sheet with reliabilty level %97.

### 2.2 Detailed Statistical Analysis for South Africa

In this section, we aim to provide a detailed statistical analysis of the collected data representing the evolution COVID-19 spread within the republic of South Africa. These data include, daily number of new infected and daily numbers of deaths. The collected data are from 5 March 2020 to 3 May 2020 [7]. The main aim of this section is to predict what could possibly happen in the near future using the reliability level method, additionally, to find which distribution each class follows. With the collected data, we will first present histogram, pie chart and nonlinear graphs for each class. The histograms will help identify the density of probability associated to each set of collected data. Additionally, we provide a polynomial fitting against collected. The results are presented in Figure 21 to 30. For each case, we present arithmetic, geometric, harmonic means respectively, skewness, variance, covariance, Pearson correlation and Spearman correlation and their results are presented in Table 2.

In Figure 21, 22 and 23, we present some statistical simulation about number of infected people due to COVID-19 in South Africa from 5 March 2020 to 3 May 2020.

**Figure 21.**
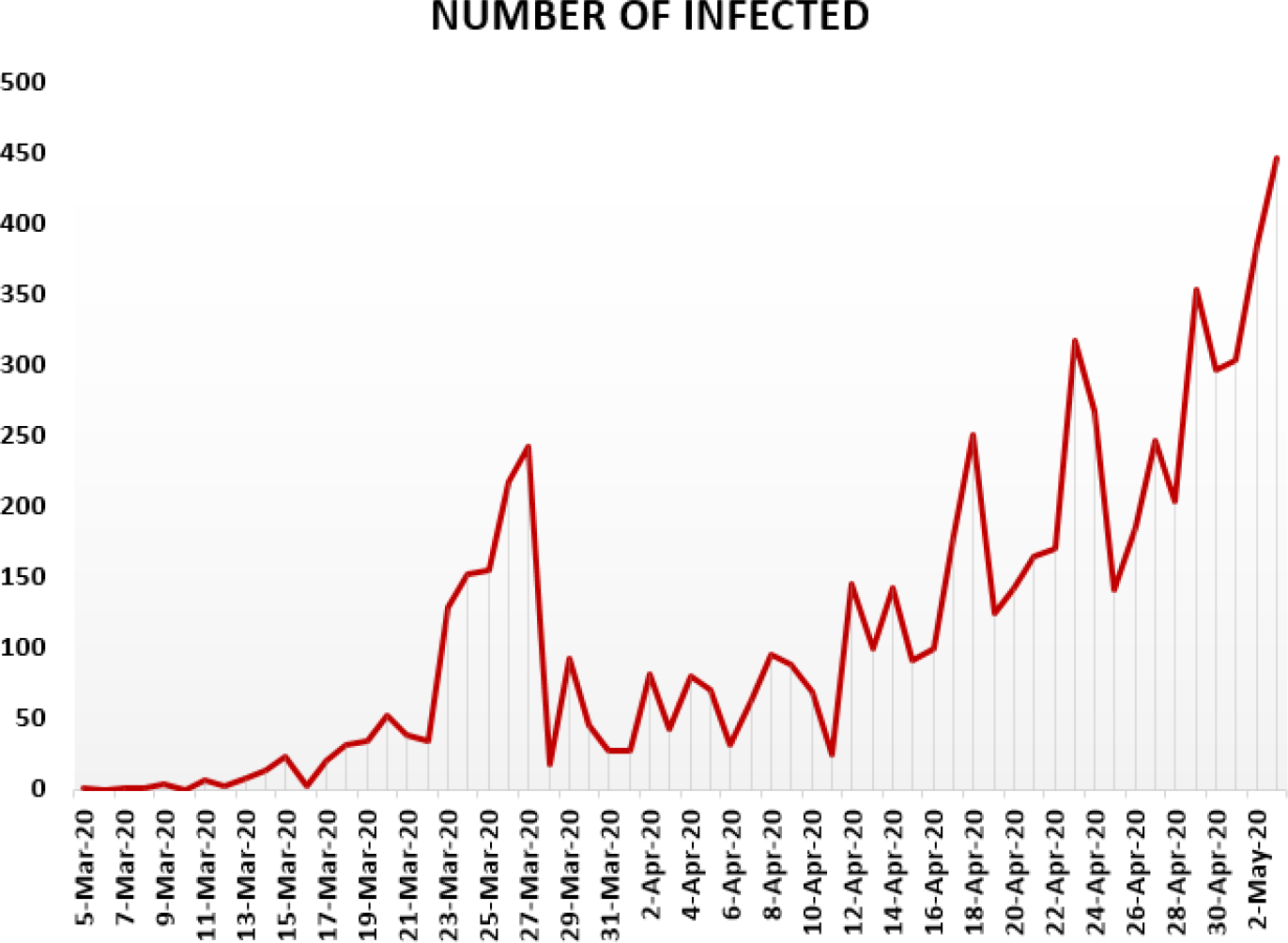
Number of infected people in South Africa from 5 March 2020 to 3 May 2020.

**Figure 22.**
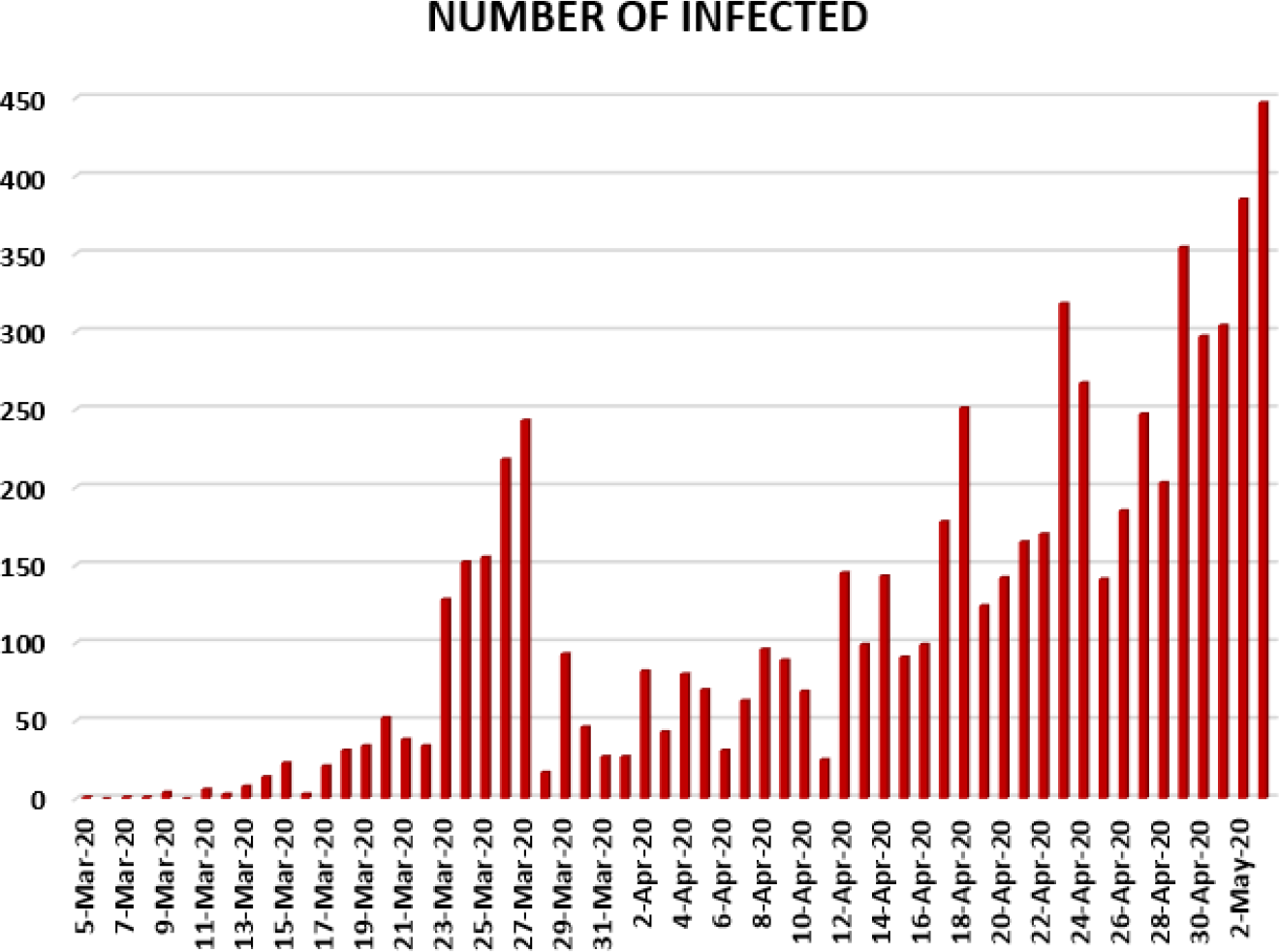
Number of infected people in South Africa from 5 March 2020 to 3 May 2020.

**Figure 23.**
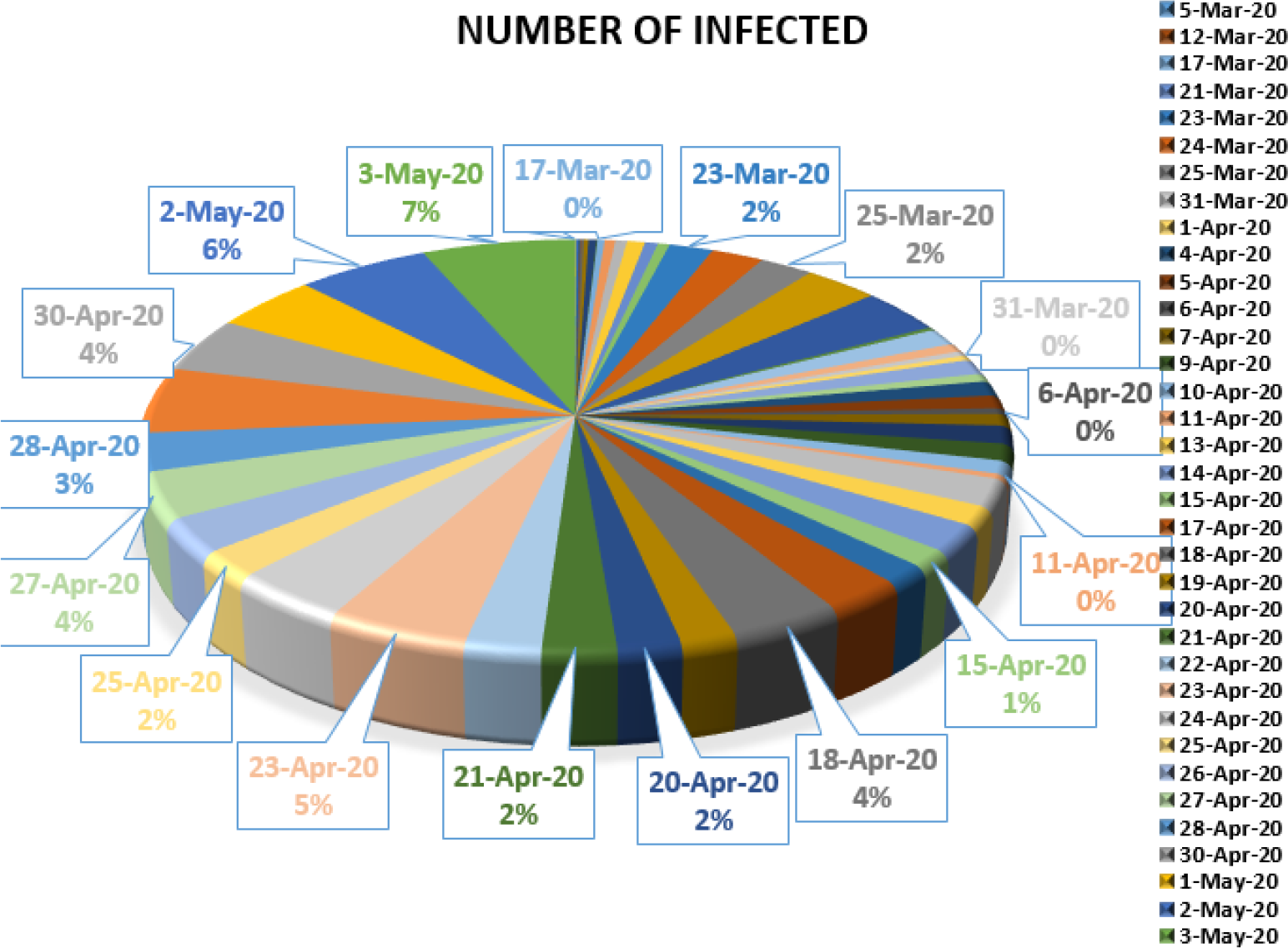
Number of infected people in South Africa from 5 March 2020 to 3 May 2020.

In Figure 24, 25 and 26, we present some statistical simulation about number of died people due to COVID-19 in South Africa from 15 March 2020 to 3 May 2020.

**Figure 24.**
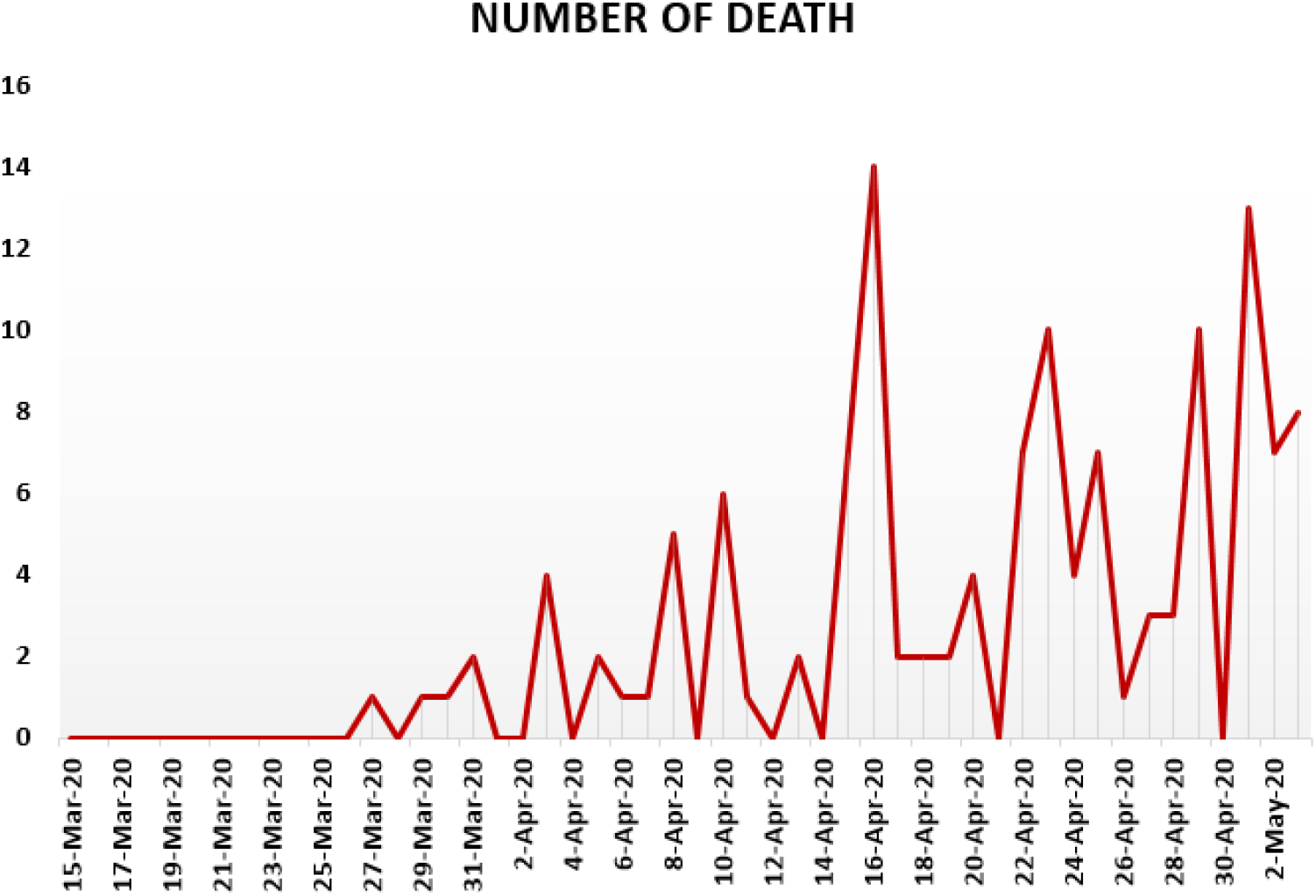
Number of death in South Africa from 15 March 2020 to 3 May 2020.

**Figure 25.**
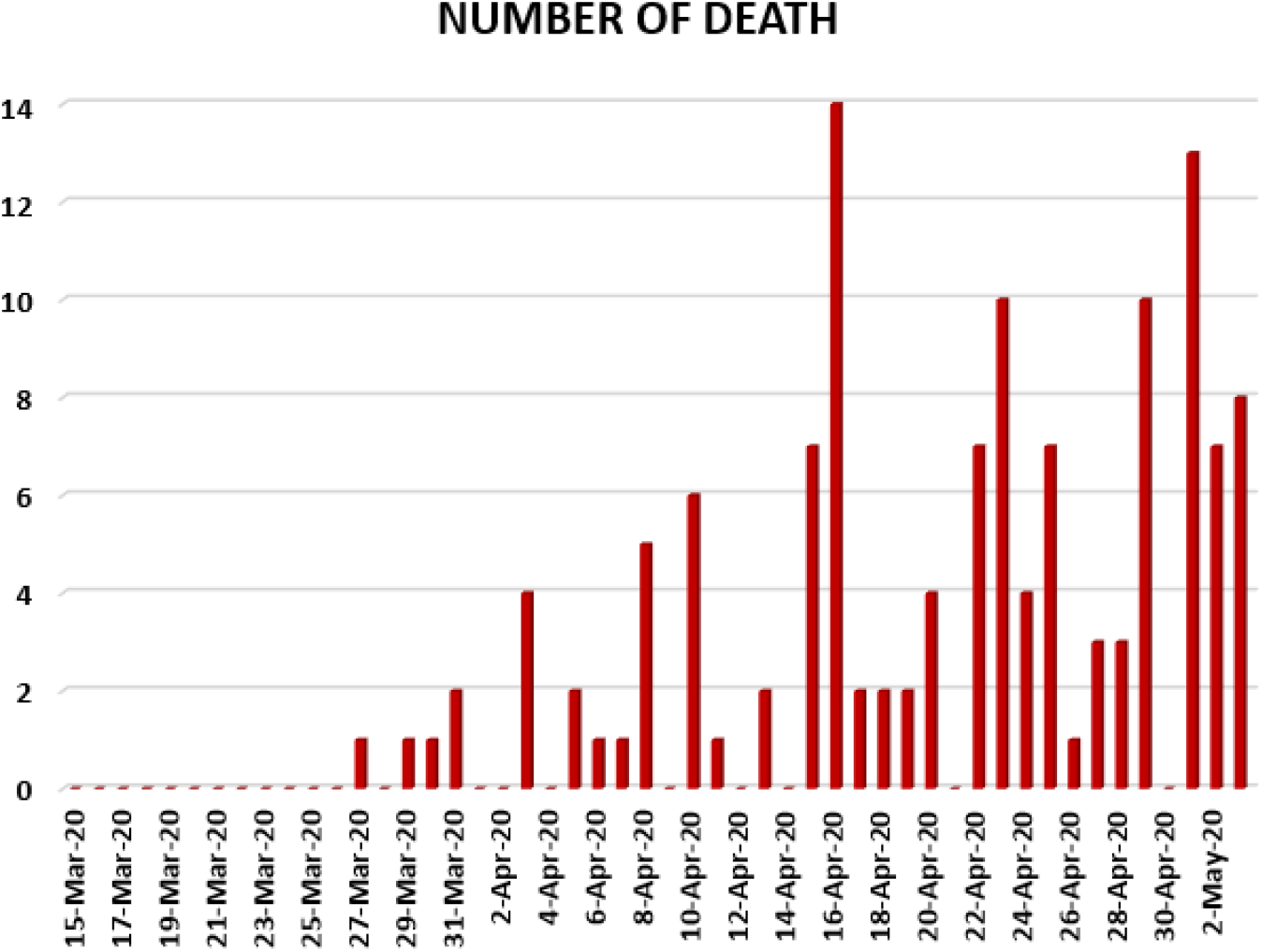
Number of death in South Africa from 15 March 2020 to 3 May 2020.

**Figure 26.**
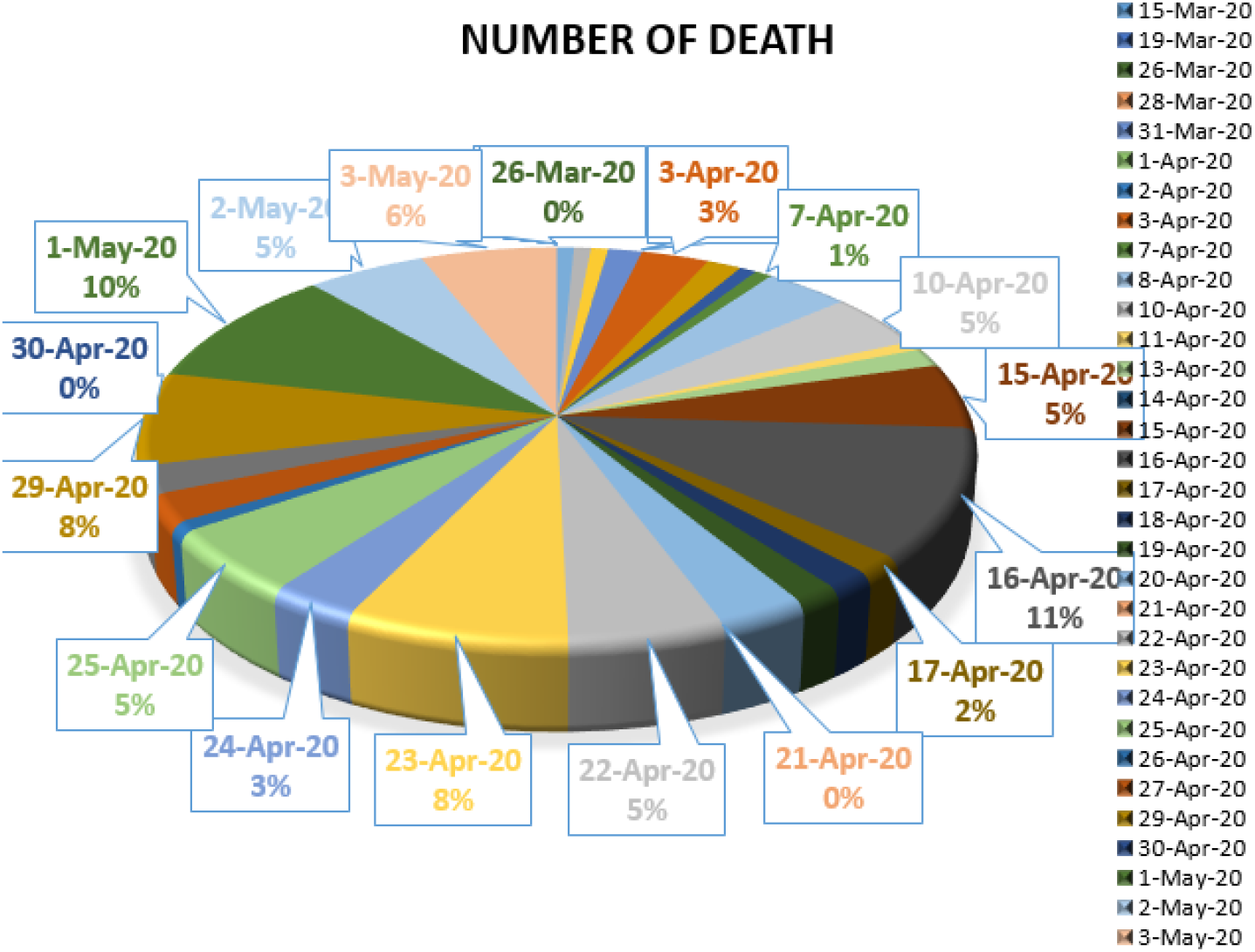
Number of death in South Africa from 15 March 2020 to 3 May 2020.

**Figure 27.**
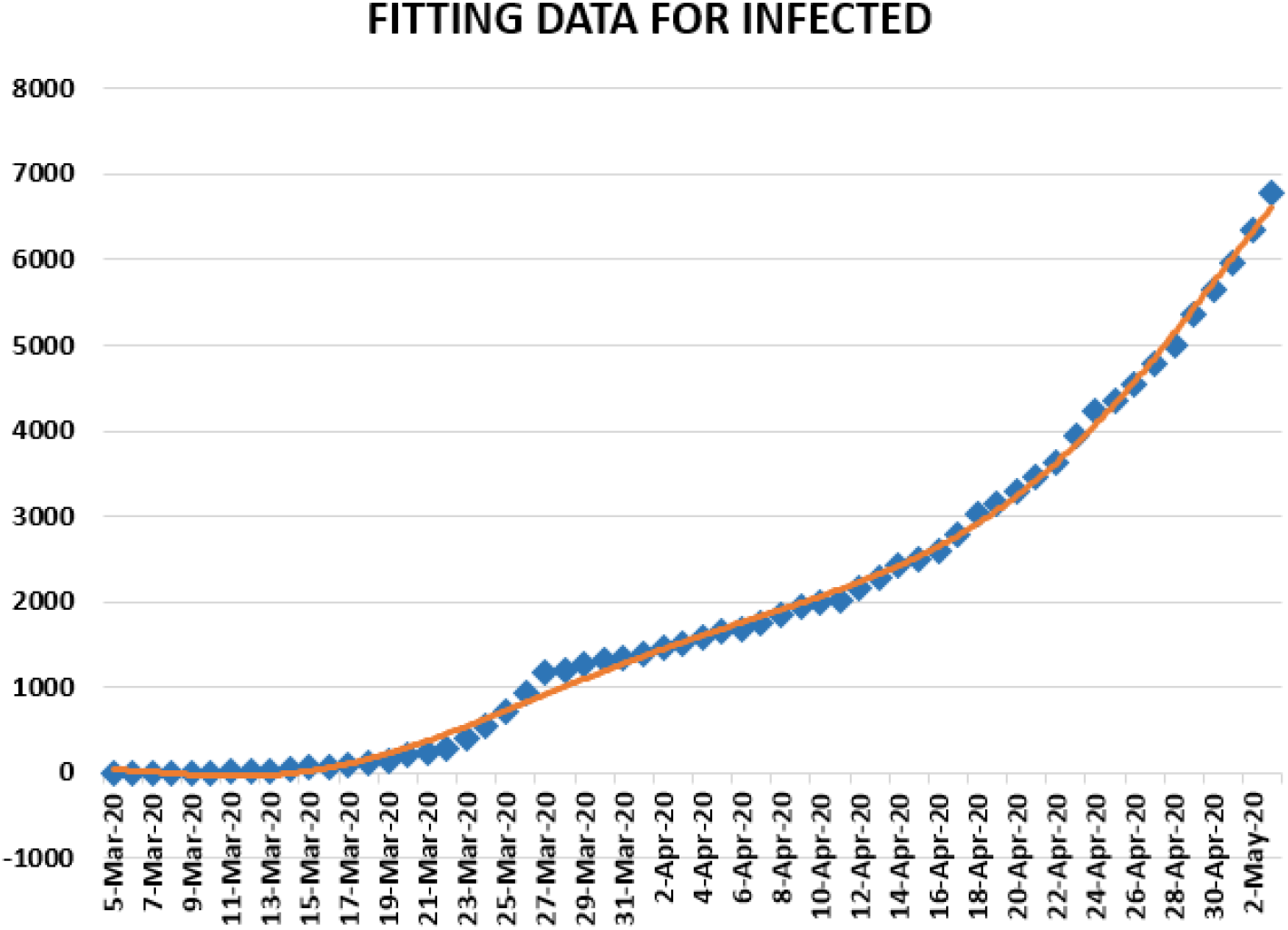
Polynomial fitting data for infected in South Africa from 5 March 2020 to 3 May 2020.

**Figure 28.**
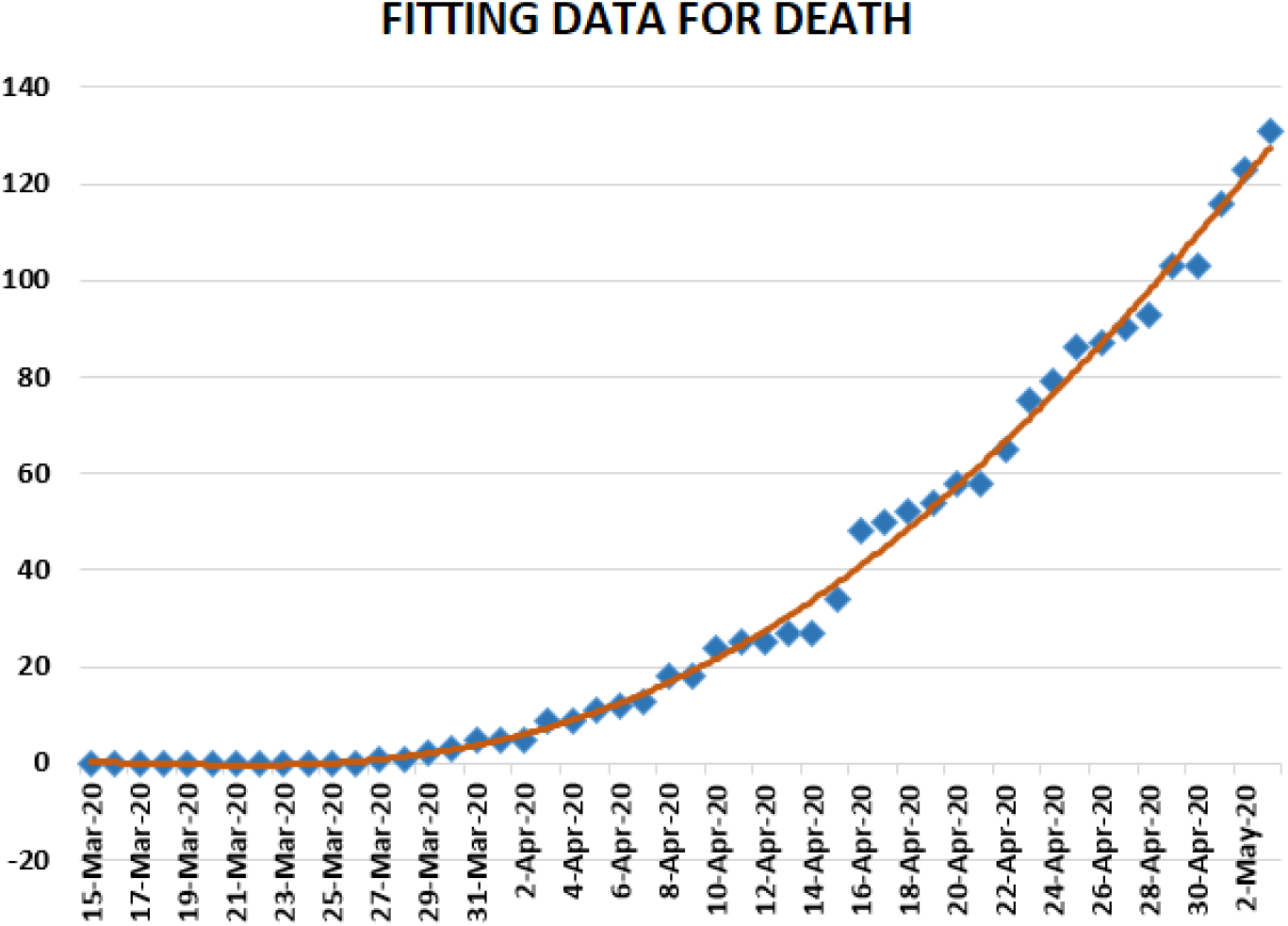
Polynomial fitting data for death in South Africa from 15 March 2020 to 3 May 2020.

**Figure 29.**
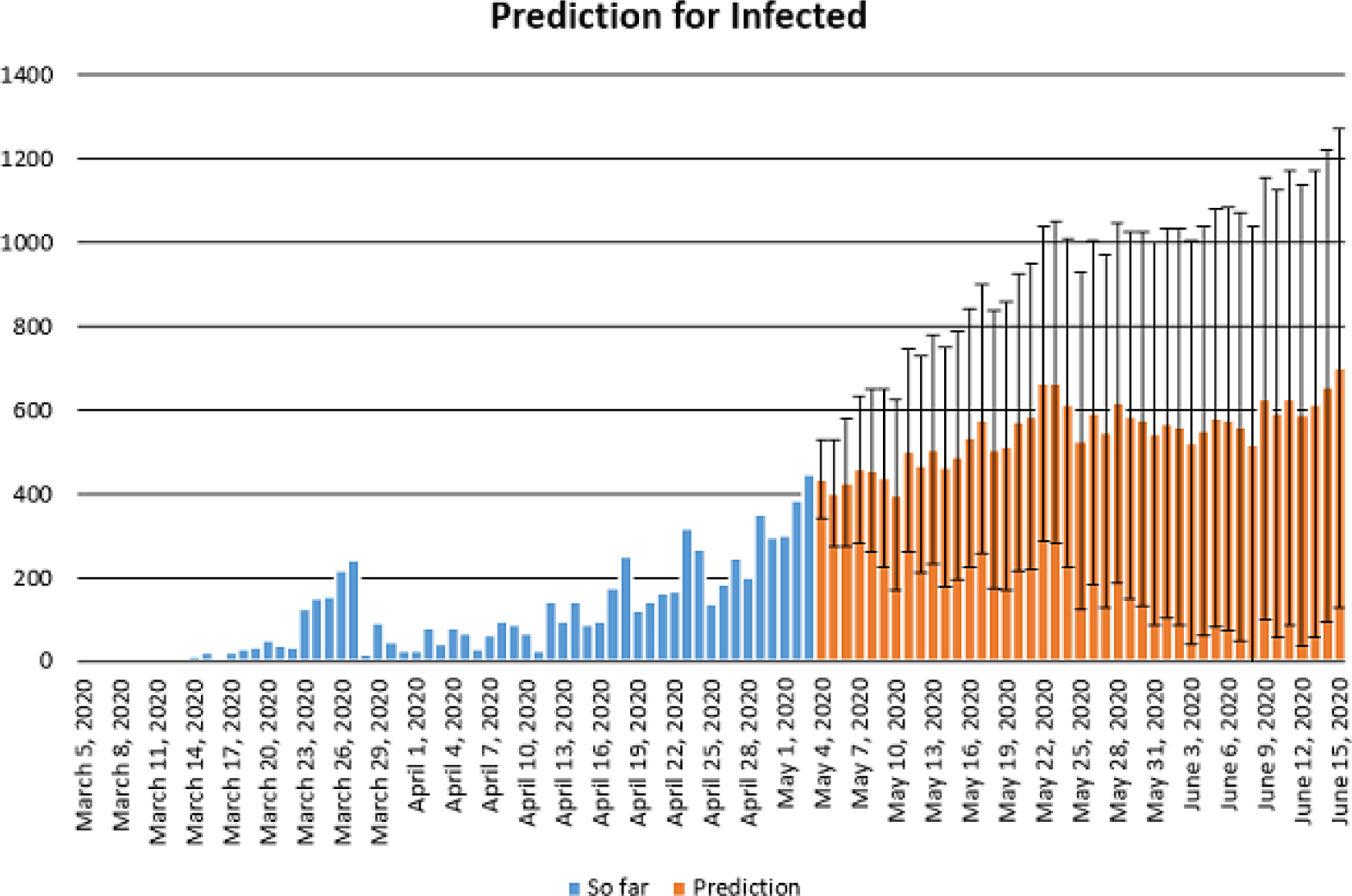
Prediction of daily number of infected in South Africa using Forecast Sheet with reliabilty level %90.

**Figure 30.**
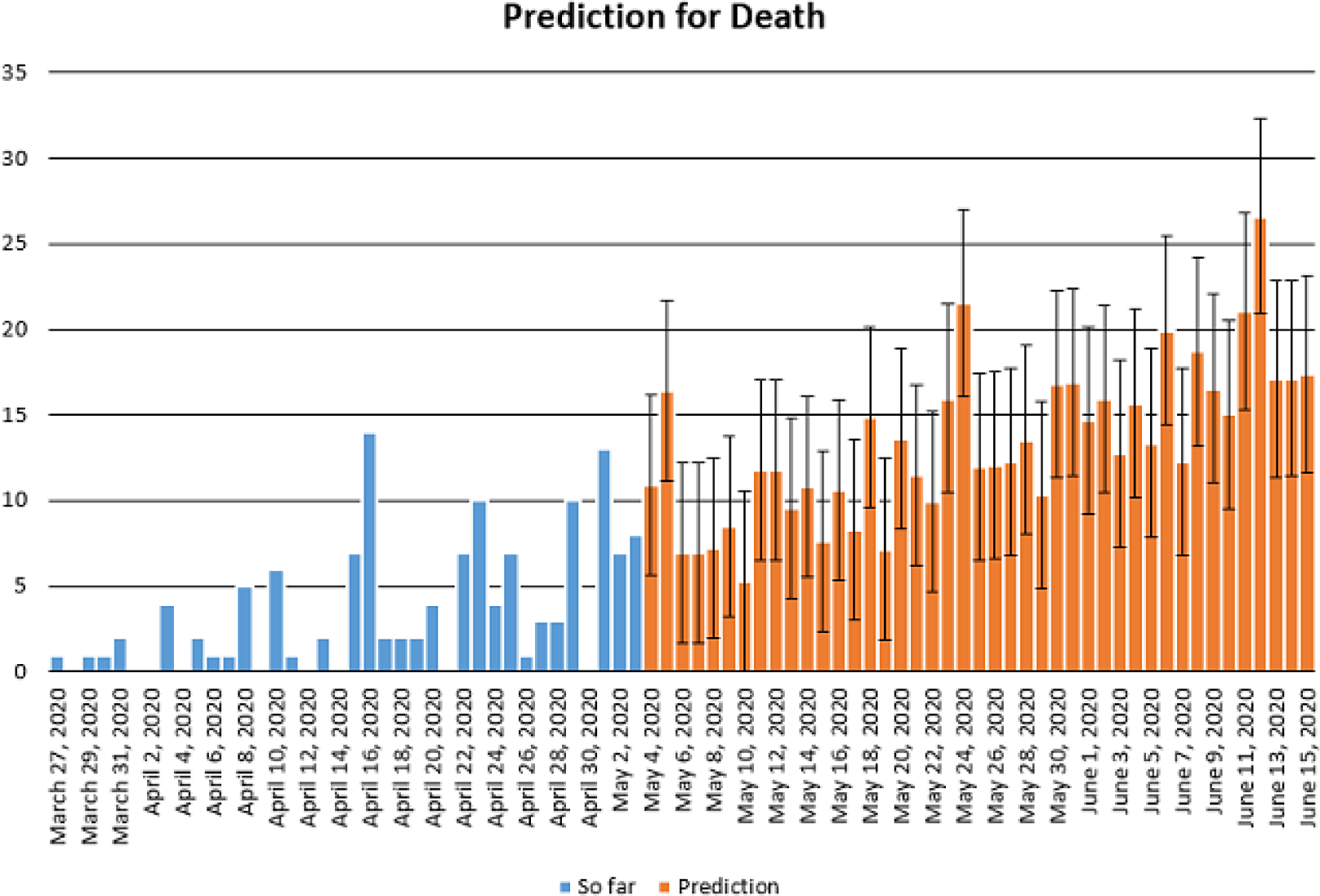
Prediction of daily number of deaths in South Africa using Forecast Sheet with reliabilty level %90.

Now we present regression analysis about COVID-19 in South Africa from 5 March 2020 to 3 May 2020. We firstly present a predictive analysis for infected people. According to the results obtained, we get a linear regression which can be calculated as

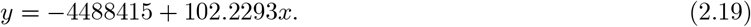

*F* – *test* was calculated as 4.84 × 10^−31^. *R square* was calculated as 0.902781. We can say that the significance of fit for model is suitable for the considered data. We can give another regression model

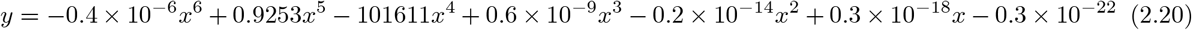

which is polynomial of sixth order. For this polynomial, *R square* was calculated as 0.9978. We present a simulation about polynomial fitting data for infected people from 5 March to 3 May 2020.

Now we present a predictive analysis for died people. According to the results obtained, we get a linear regression which can be calculated as

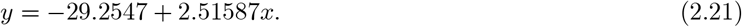

*F* – *test* was calculated as 5.36 × 10^−22^. *R square* was calculated as 0.858225. We can say that the significance of fit for model is for the considered data. We can suggest another regression model

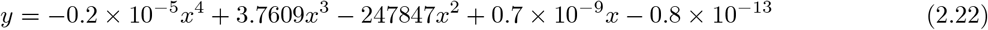

which is polynomial of fourth order. For this polynomial, *R square* was calculated as 0.9958. We present a simulation about polynomial fitting data for died people from 15 March to 3 May 2020.

#### 2.2.1 Prediction about corona data in South Africa

In this section, we aim at performing prediction using existing data collected representing daily numbers of new infected, deaths and reliability level method. The collected data will be considered from 5 March 2020 corresponding to the first day of confirmed case of COVID-19 in South Africa to 3 May 2020. The future prediction will start from 3 May 2020 to 15 of June 2020. This will help us give a prediction on numbers of new daily infected class, recovered, daily numbers of deaths in South Africa within this period. The prediction will consist of three different graphs comprising upper boundaries, middle lines and low boundaries. The upper boundaries represent the worse cases scenario, of course a scenario that is not needed for deaths class and infected but an ideal one for recovered class, and the lower boundaries representing perfect scenarios (A scenario that is needed) for South Africa to get rid of the infection. These results of prediction for future daily new infected, recovered and deaths are represented graphically in Figure 29 and 30 respectively. The prediction of daily new infected in the case of South Africa seems to follow the upper boundaries and low boundary for daily number of deaths.

We now give some data about corona in South Africa in Table 3.

**Table 3.** Some data about corona in South Africa.

|  | **Infected** | **Death** |
| --- | --- | --- |
| **Arithmetic mean** | 113,05000 | 2,62000 |
| **Geometric mean** | 57,35444 | 3,184267 |
| **Harmonic mean** | 11,569120 | 2,260659 |
| **Standard deviation** | 109,71834 | 3,613410 |
| **Skewness** | 1,1222456 | 1,594677 |
| **Variance** | 11837,4808 | 12,79560 |

Table 4 presents the covariance, the Pearson and Spearman correlation coefficients between daily cases-recovered, recovered-death and infected-death about COVID-19 in South Africa.

**Table 4.** Some data corona in South Africa.

|  | **Infected-Death** |
| --- | --- |
| **Covariance** | 209,4908333 |
| **Pearson Correlation** | 0,564936726 |
| **Spearman Correlation** | 0,607349264 |

We present lognormal distribution for infected and death cases about COVID-19 in Figure 31.

**Figure 31.**
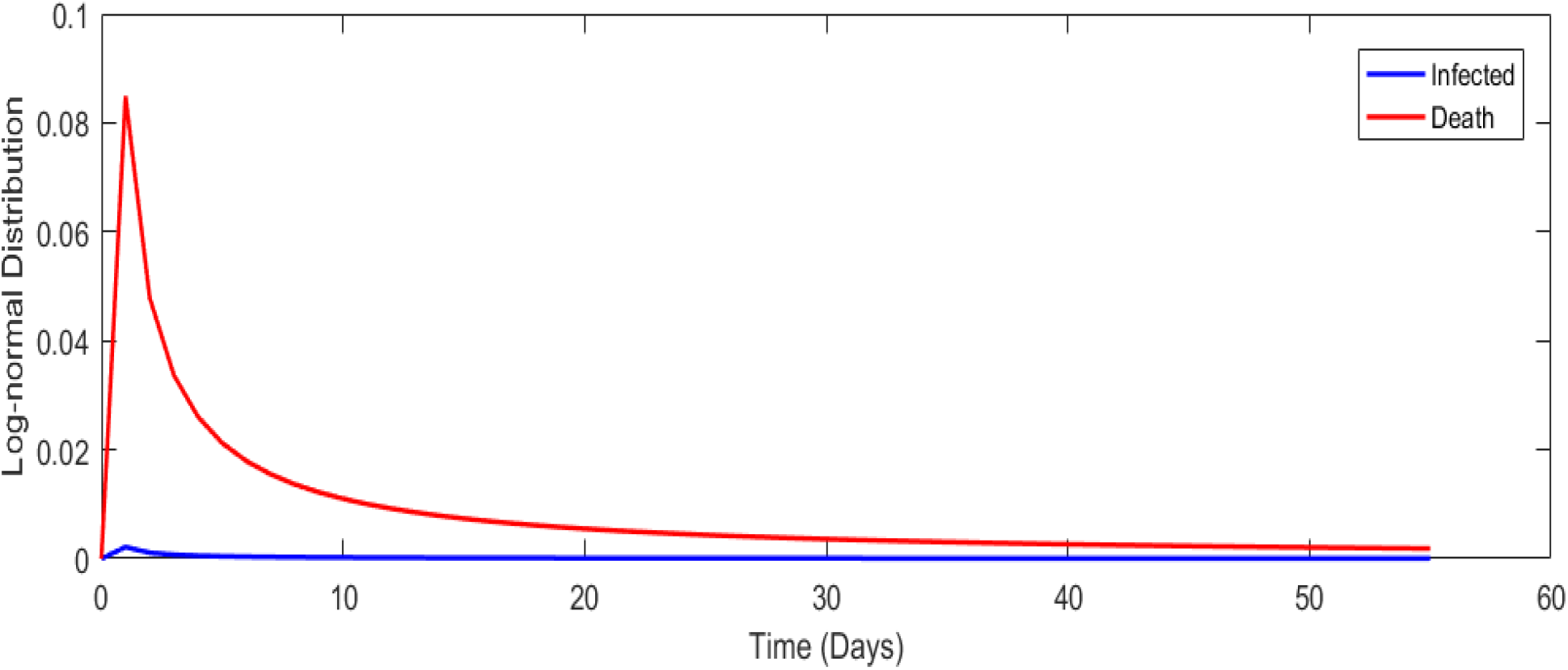
Lognormal distribution for all cases in South Africa.

**Figure 32.**
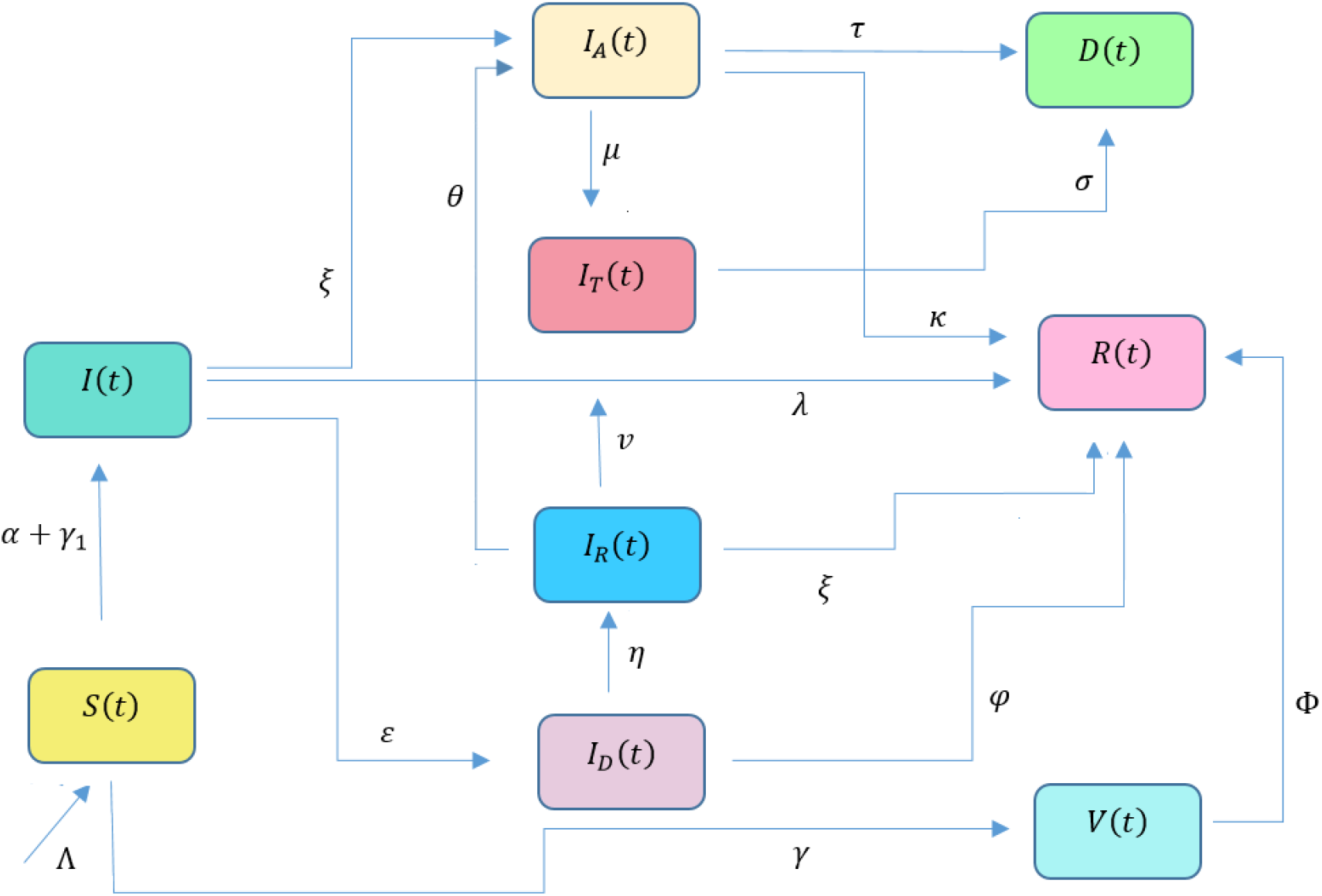
Diagram summarizing COVID-19 model given by the system (3.1).

### 2.3 Parameters estimation using the Bell curving approach

In the previous section,we presented the graph of a day-to-day evolution of COVID-19 spread including infected, recovery and death for South Africa and Turkey. To be honest, one cannot for sure tell if those curves follow the normal distribution or lognormal distribution. Therefore in this section, two cases are considered. In the first case, we assume a lognormal curve and second we assume normal distribution curve.

**Case I:** We consider the lognormal density of probability1 1

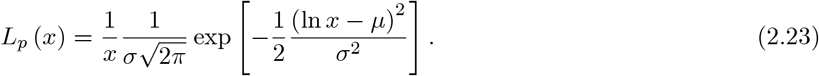

We now define a function *β* that captures daily occurrences

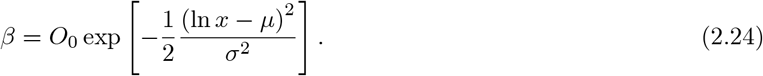

We aim to estimate *O*_0_, *σ* and *μ* achieve this, we consider first four different days

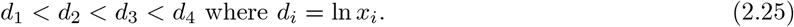

We first start by estimating *μ*, by assuming a proportion

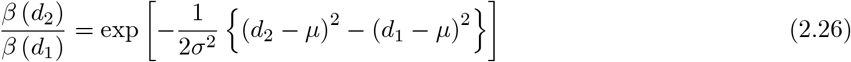

and

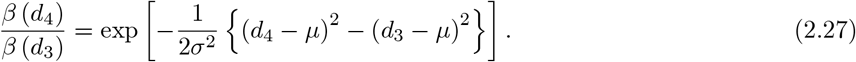

To proceed, we apply on both sides the ln function

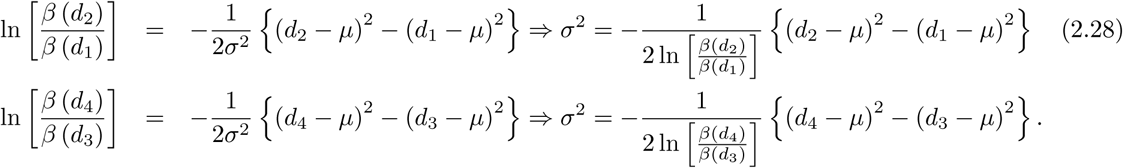

Due to the equality, we can now have

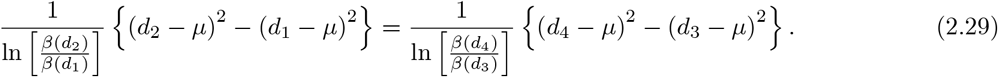

Thus

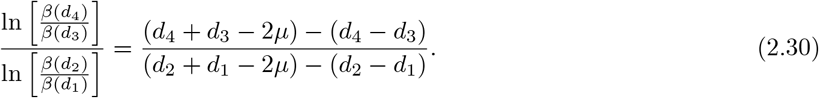

The solution is the above is

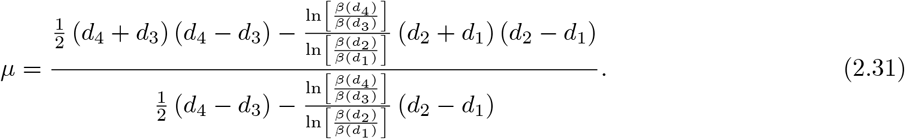

Having *μ*, we can determine

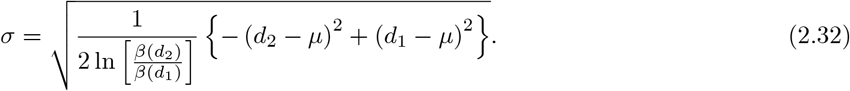

Alternatively, we consider 8 days to capture more facts *x_i_*, *i* = 1, 2, 3, 4, 5, 6, 7, 8, we put *d_i_* = ln *x_i_*. We assume a proportionality λ of {*d*_1_, *d*_2_, *d*_3_, *d*_4_} and {*d*_5_, *d*_6_, *d*_7_, *d*_8_}. Therefore

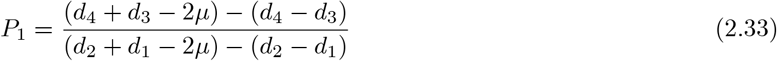

and

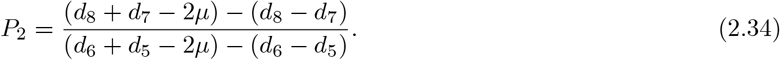

We now assume that *P*_1_ is proportional to *P*_2_ then

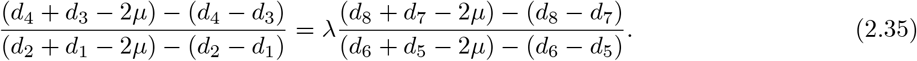

For simplicity, we put *d_i_* + *d_j_* = *A_ij_* = *A_ji_*

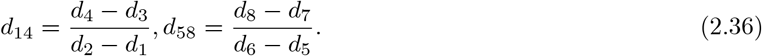

Therefore, the above can be reformulated as

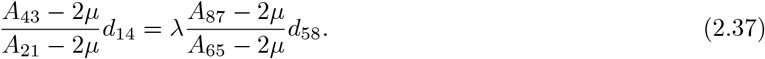

Also we write

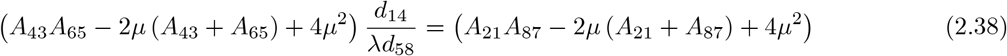

and

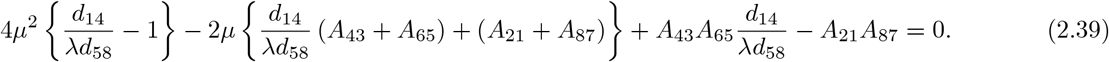

Thus we have

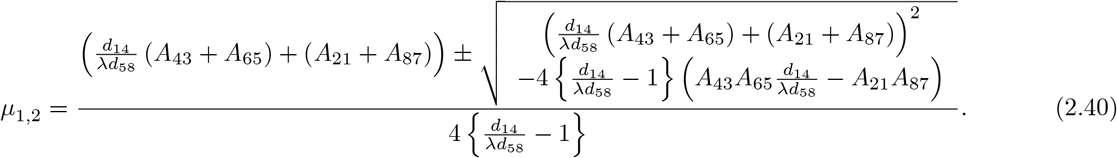

Thus for Case I, we get

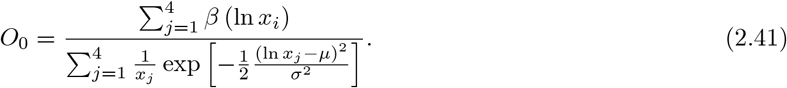

Case II, we get

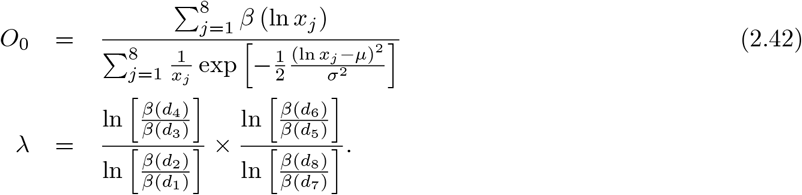

For each case, the cumulative distribution function can be calculated by

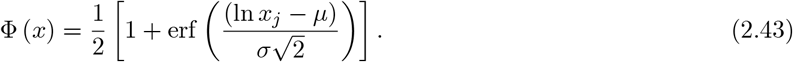

**Case II:** We assume that the curve follows the normal distribution, thus

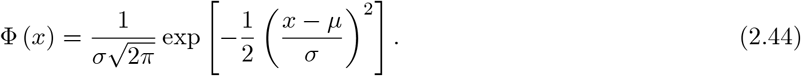

However, we consider the following function

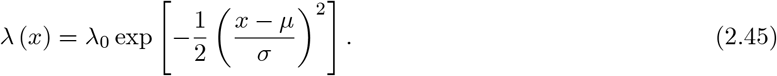

We aim to determine λ_0_, *σ* and *μ*. Here we choose three points *d*_1_, *d*_2_, *d*_3_ such that λ (*d*_2_) corresponds to the maximum point. Following the procedure presented earlier, we have

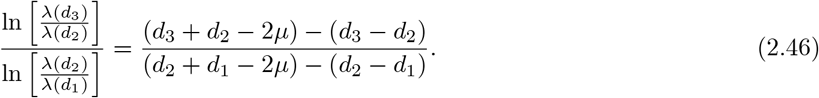

Thus

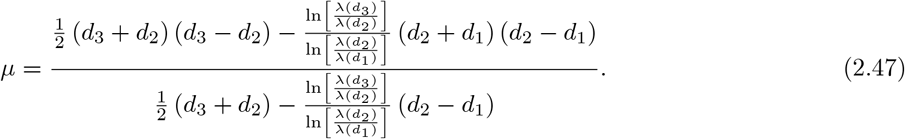

With *μ* in hand, we determine

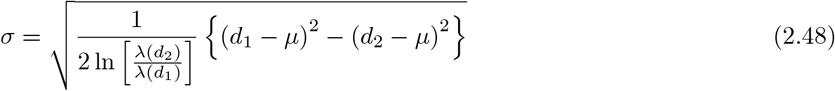

and

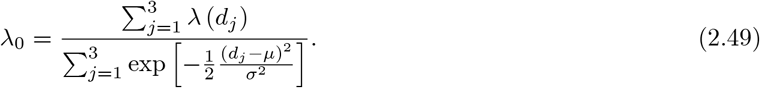

Particular case if we consider the case where *d*_2_ − *d*_1_ = *d*_3_ − *d*_3_ that is λ (*d*_1_) = λ (*d*_3_) due to symmetry of normal distribution, then we get

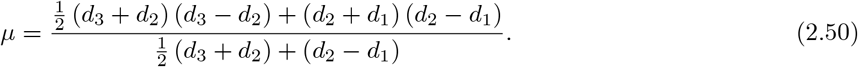

### 2.4 Comparison: Turkey vs South Africa

In this subsection, we present a comparison between Turkey and South Africa about COVID-19.

**Table 5:** Comparison between Turkey and South Africa updated on COVID-19

| Country | South Africa:5 March to 02 May 2020 | Turkey:11 March to 02 May 2020 |
| --- | --- | --- |
| Number of infected | 6336 | 124375 |
| Number of death | 123 | 3336 |
| Number of recovered | 2549 | 58259 |
| Total tested | 230686 | 1,033,617 |
| Lockdown date | 26 March 2020<br>(Total lockdown) | 11April 2020, (Partial lockdown:<br>Less than 20 and<br>older than 65) |
| Donations | NA | ₺ 1,865,799,782 up to<br>28 April 2020 |

The analysis presented in this section does not aim at praising nor criticizing any country; but just to assess the effect of lockdown and its regulations, and to perceive if this concept can help save humans before the vaccine. The fundamental question to answer here is to know why South Africa has less number of deaths and infected people than Turkey; if it recorded its first confirmed case six days earlier before Turkey recorded its first. The answer may rely on two fundamental facts which include the period lockdown was implemented and the type of lockdown put in place. The South African government publicized on 23 March 2020 a 21-day of national lockdown which started effectively from midnight 27 March. This was announced 22 days after the first confirmed case was recorded in the country. The lockdown came with strict measures encompassing, immediate deployment of South African National Defence force to ensure that all people living within the territory of South Africa obey the lockdown rules. Only workers considered necessary to operative response to the pandemic were exempted, namely: health caregivers, security service providers, essential service providers that are fundamental to the rudimentary functioning of economy as well as other workers in industries that cannot be economically shut down. This implies that the mentioned categories were permitted to go to their places of work during the lockdown. On the other hand, the numbers of people at gatherings apart from funerals were limited to 50 people; while restaurants, taverns, bottle stores and shops that are not selling indispensable goods were forced to close. Thus, a large population was not allowed to leave their houses except for essential needs. Consequently, the movement between provinces, metropolitan and districts were also restricted unless for essential reasons that cannot be catered for within provincial boundaries. The South African government further closed all of its national borders and only allowed the transportation for indispensable reasons. Likewise, all international and domestics passenger flights were prohibited, except those assigned to evacuate citizens from foreign countries and certain repatriations due to COVID-19. However, the measures taken by Turkey were not implemented swiftly upon the confirmation of its first COVID-19 case. It is recorded that the Turkish government announced a partial lockdown on 11 April 2020, a month after the country registered it first confirmed case of COVID-19. Prior to the announcement of the partial lockdown, mosques cafes, night clubs, and all universities within the country were already closed on 11 March 2020. The restriction measures applied only on people younger than 20 years old and older than 65 years old, who were not allowed to leave their homes except for indispensable reasons. In addition, the government ordered a ban on movements between 30 major cities with metropolitan status as well as Zonguldak; whereby the lockdown is applied every weekend since 11 April 2020 and also 21-23 April 2020. Punishment (money) is applied for people who go out. Very importantly the government have totally banned the sale of masks, but provided free masks to its people to be compulsory utilized in public places. However, the newly placed order exempted health care assistance, funerals, military and passenger transports from the ban; provided that certain conditions are met. Although both countries have put severe measures to protect their citizens from the deadly disease; there are still records of rising number of infected and dead people in both Turkey and South Africa. Does that mean that the lockdown regulations are worthless? Absolutely not! It is only that several citizens in respective countries are not adhering to the rules and regulations put in place by their respective government authorities. This results from the concept of social distancing being largely misunderstood, as it is not clearly defined to mean whether persons should stay one meter away from one another or only from any infected humans, contaminated air and other objects because of the nature in which COVID-19 can be spread. Nevertheless, due to a long incubation period of COVID-19, approximately 14 days maximum; which renders ordinary citizens not to differentiate an infected person from others; then it is crucial that stringent measures be implemented, which will prohibit people from leaving their homes; and in case they had gone out, they should maintain the one metre distance away from each other and frequently wash their hands upon touching any object.

## 3 Mathematical model of COVID-19 in South Africa and Turkey

Mathematical models of infected diseases are deemed not that useful by some people who feel that they cannot be utilized to develop a vaccine or cure of any given disease. However, it is important to note that the principal aim of these mathematical models is to describe a system using mathematical tools, concepts and language. Hence, throughout the history of human beings, researchers working within the field of mathematics have developed more accurate and efficient mathematical models. For instance, history has made reference to one of the well-known Newtonian laws which described very accurately many problems in our daily lives; although they are coupled with some limits. In instances where these laws failed, two other well-known concepts namely; theory of relativity and quantum mechanics using mathematical formulas can be utilized instead. Generally, these concepts are of great importance in all fields of science such as in natural sciences including chemistry, biology, physics, and earth science, in engineering such as computer science, and electrical engineering, as well as in social science where their applicability to economics, sociology, psychology and political science can be relevant. In other words, mathematical models can help to provide a clear explanation of a system and investigate the effect of several components, and later make an accurate predictions based on the observed facts. In the current situation under study, due to the magnitude of fear imposed by COVID-19 on humans, it is therefore paramount for mathematicians to provide conceptual models, using mathematical tools called differential and integral operators, to suggest well-constructed mathematical models that will be used to understand and predict the spread of COVID-19.

In this section, a mathematical model that takes into account nine classes (susceptible, infected which has 5 sub-classes, recovered, death and vaccinated classes), the dynamic is presented and explained with the subsequent diagrams; but the death class is omitted because it can produce a complex model. The created model incorporates the lockdown effect, represented by a coeff cient that takes into account the social distancing and a contact coefficient.

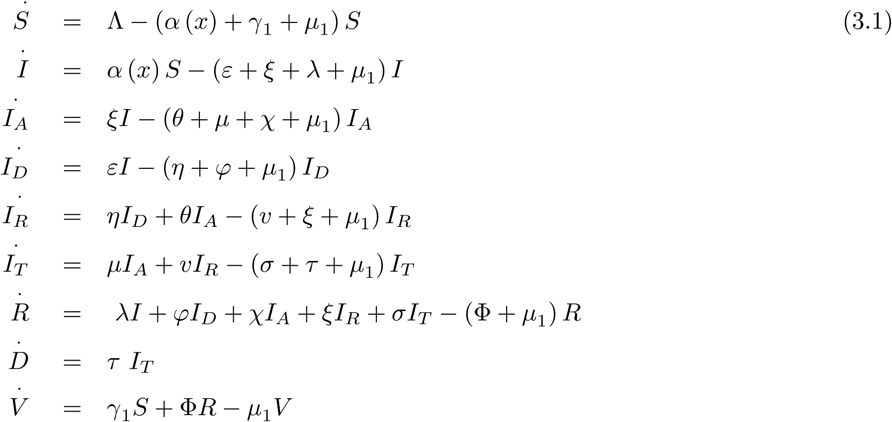

Where

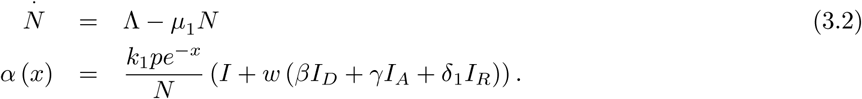

*S* (*t*) is the class of individuals that are susceptible to contact COVID-19 at time *t*. *I* (*t*) is the class of individuals that are susceptible to contacted COVID-19, but have no symptoms and have not been tested. *I_A_* (*t*) is the class of individuals that have some symptoms but not tested yet. *I_D_* (*t*) is the class of individuals that have contacted COVID-19, have been tested positive, but no symptoms. *I_R_* (*t*) is the class of individuals that have contacted COVID-19, have been tested positive and have symptoms. *I_T_* (*t*) is the class of individuals that have contacted COVID-19 and one is critical condition. *R* (*t*) is the class of recovered individuals at time *t*. *D* (*t*) is the number of death at time *t*. *V* (*t*) is the class of individuals that have been vaccinated.

**Table 6.**
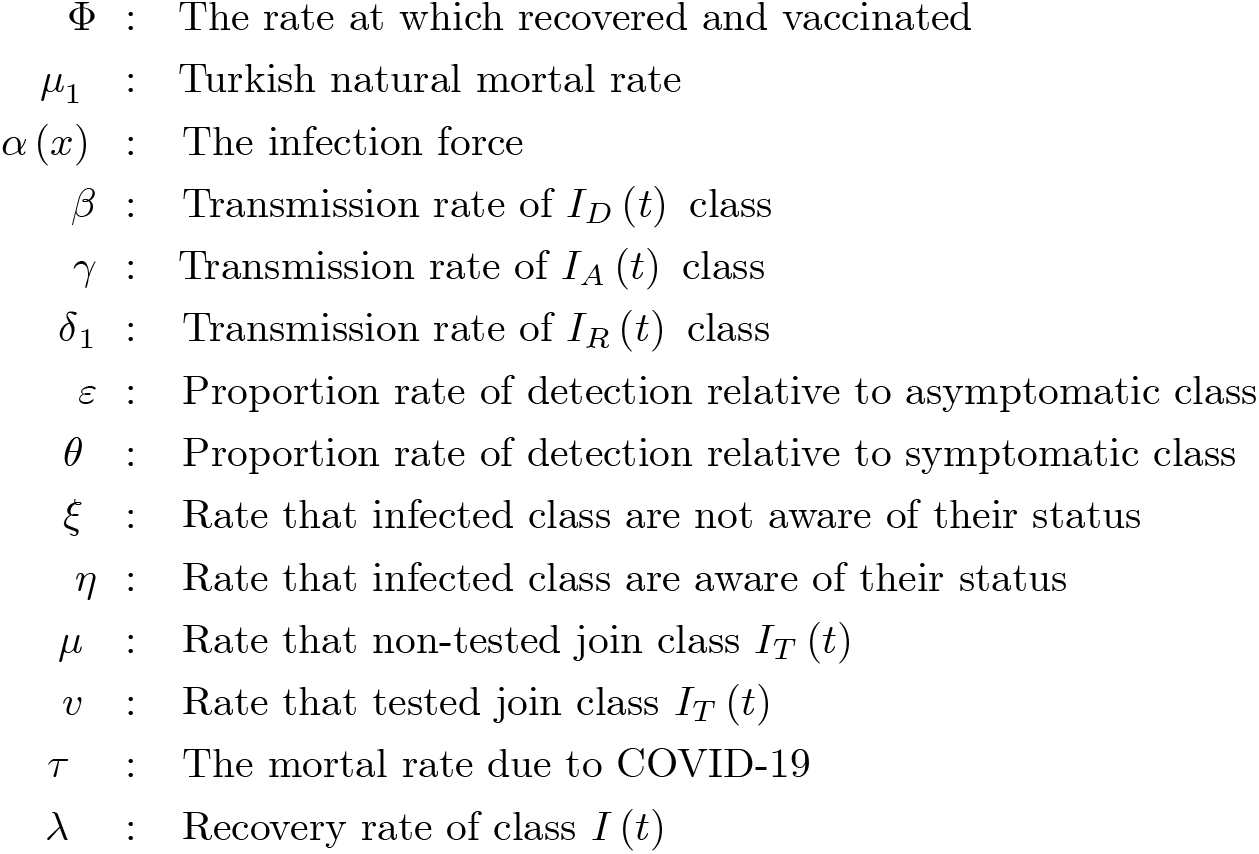
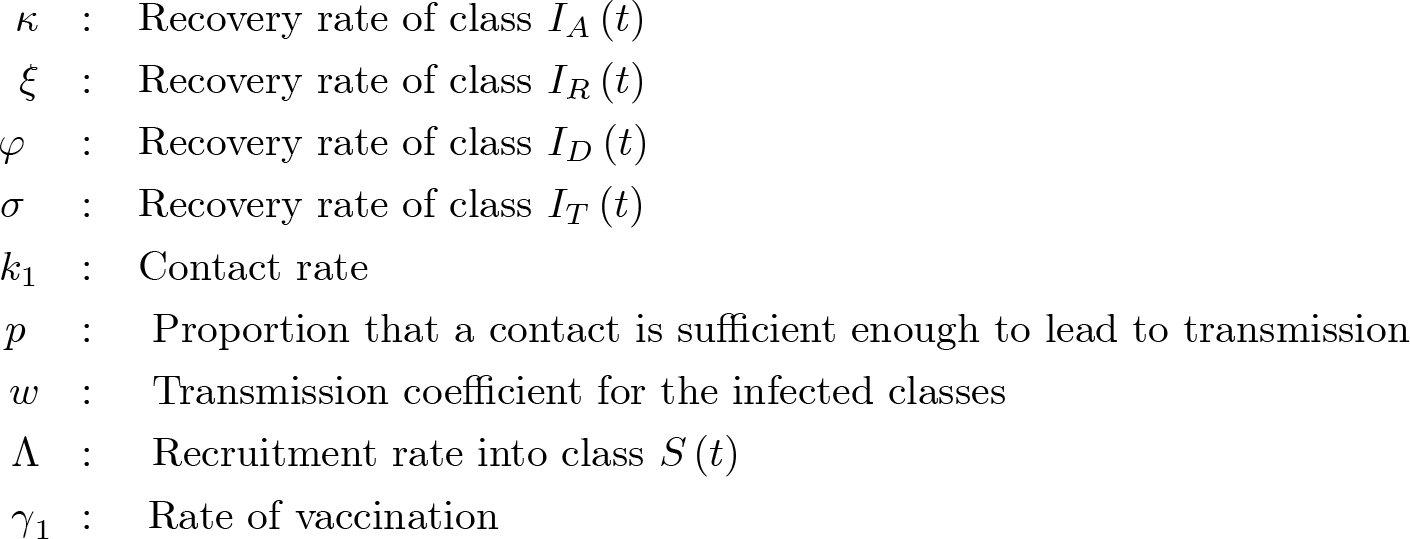
Parameters of the suggested COVID-19 model

The initial conditions are given as

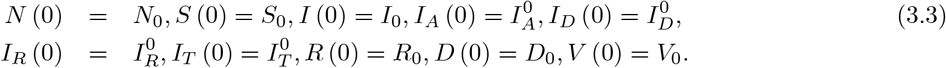

We present a diagram which summarizes COVID-19 model which is described by the system (3.1) in Figure 21.

### 3.1 Boundness and positivity of the solutions

In this section, we show that ∀*t* ≥ 0, the system solution is positive, that the model is well-posed and biologically feasible. We define the norm

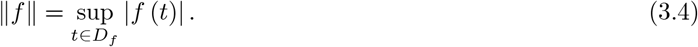

We assume that all the class

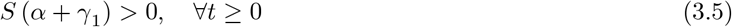

due to the model under this assumption. We write

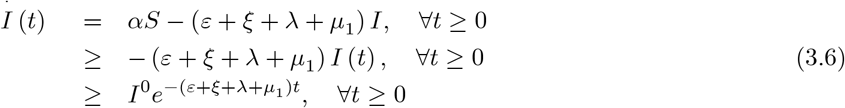

Since *I* (*t*) ≥ 0, ∀*t* ≥ 0, then

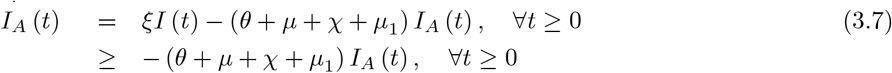

then

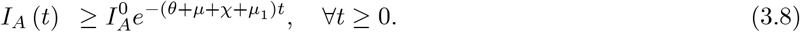

The same with *I_D_* (*t*) class

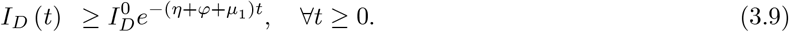

*I_A_* (*t*) and *I_D_* (*t*) are positive ∀*t* ≥ 0 and *η*, *θ* ≥ 0 then

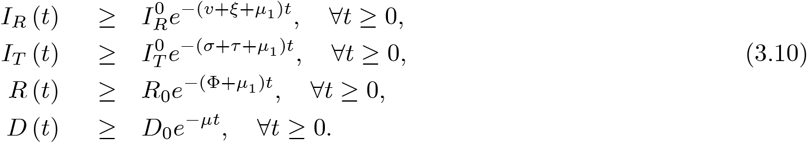

Also

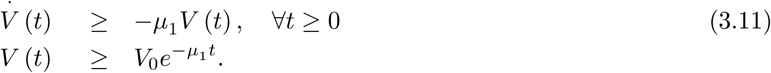

With *S* (*t*), we have to assume that

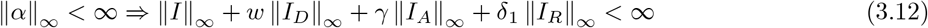

such that

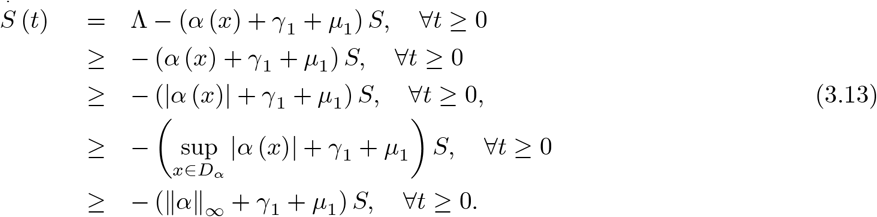

This implies that

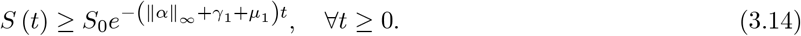

Now in the absence of the COVID-19, we have

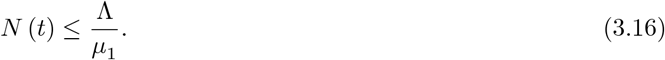

The above inequality is called threshold population level. This obtains because we assume that the total number of population must be increased or be constant

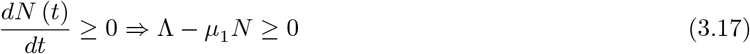

therefore 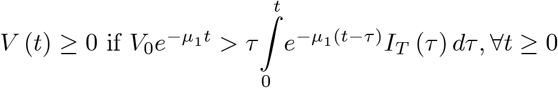. It is therefore biologically feasible that

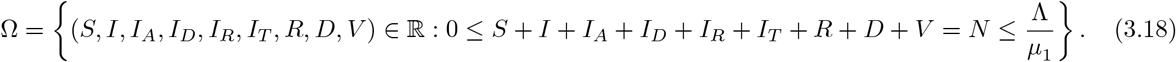

The disease free equilibrium point is

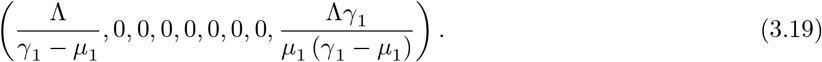

We now derive the reproduction number using the next generation operator technique [9]. We have 3 infected classes *I* (*t*), *I_A_* (*t*), *I_D_* (*t*), *I_R_* (*t*) and *I_T_* (*t*). The matrix *F* and *V* will be obtained from

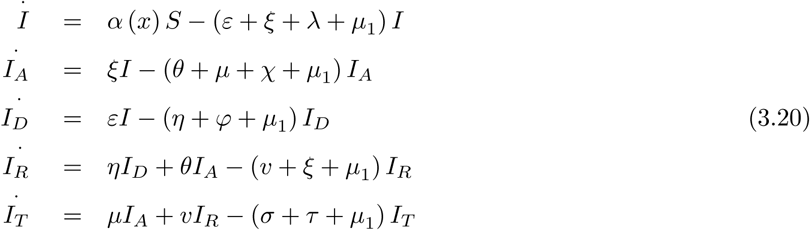

We obtain the following matrices

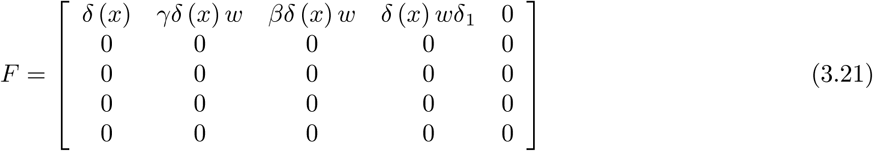

and

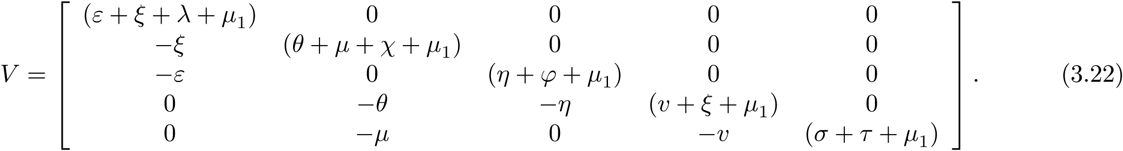

For simplicity

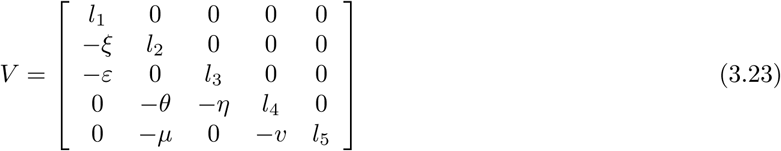

where

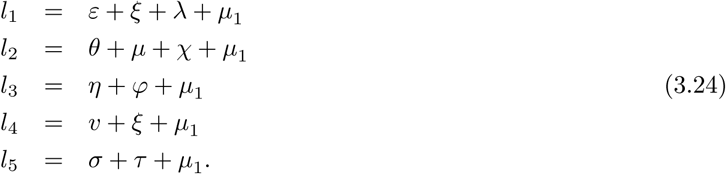

Then we have

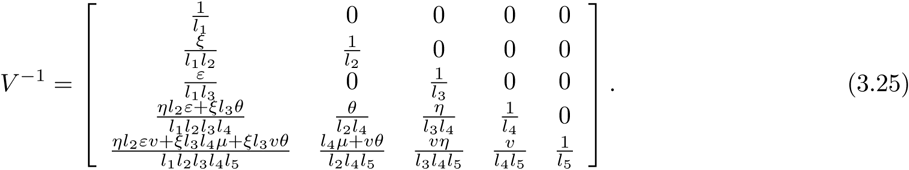

So we write the following

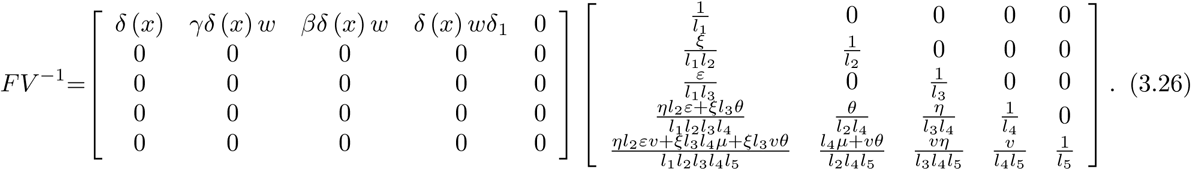

Therefore using *R*_0_ = *ρ* (*FV* ^−1^) the basic reproductive number is given as

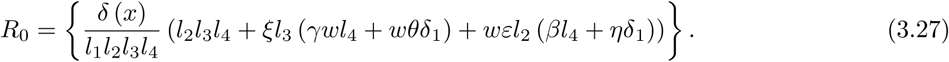

We now present disease equilibrium points. We achieve this by solving

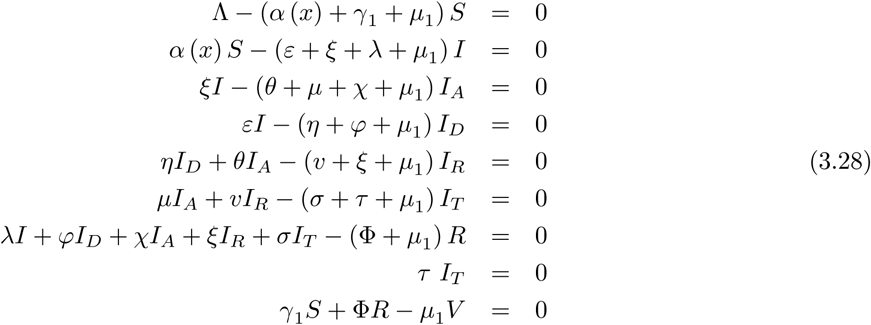

This implies that

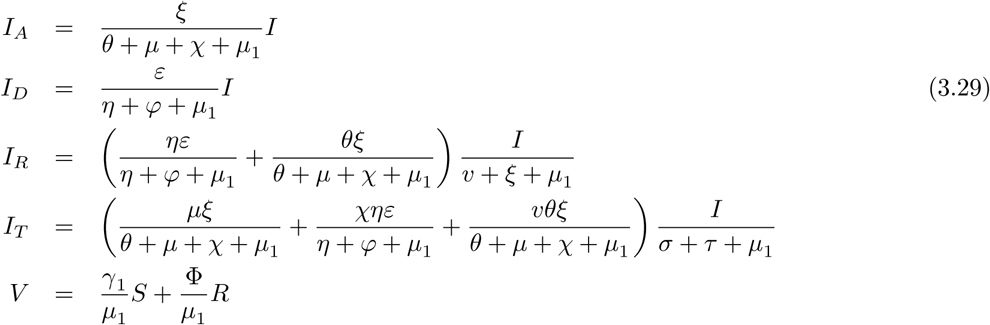

Thus

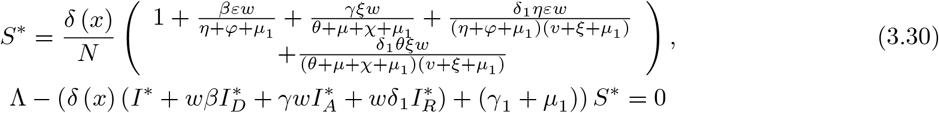

and

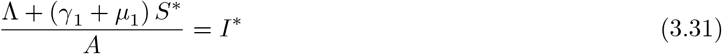

where

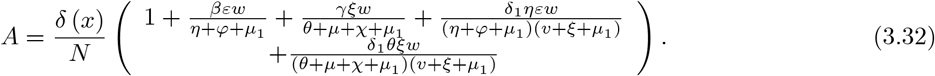

That is

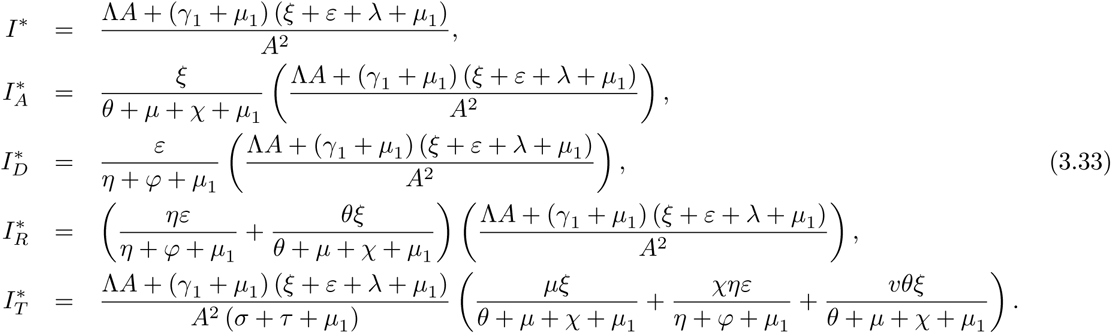

Also we get

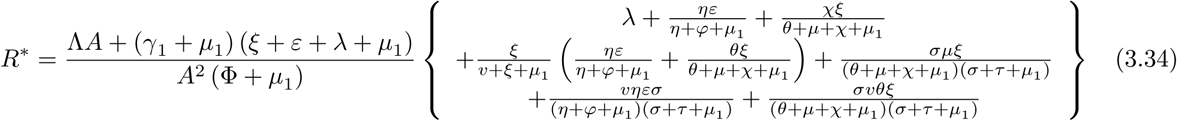

andA

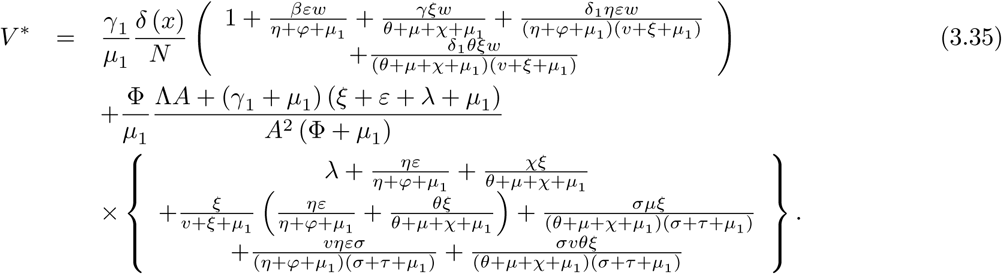

For the COVID-19 endemic with this model, we need to have

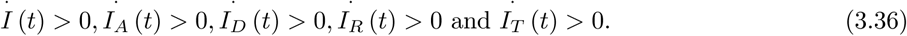

This implies

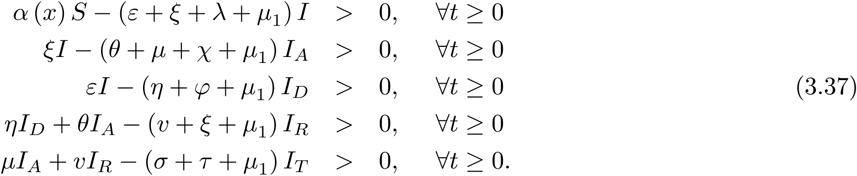

Thus

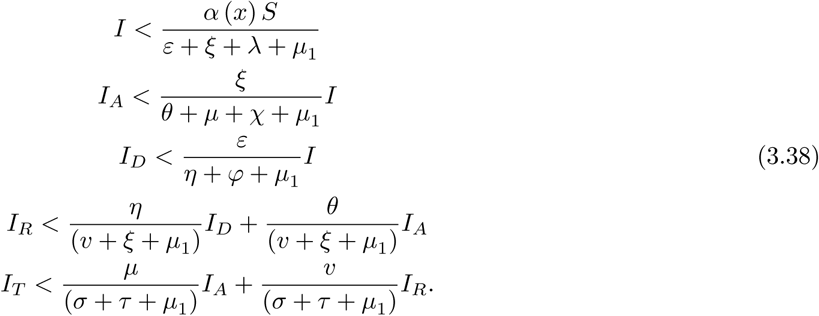

We use the fact that 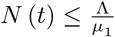

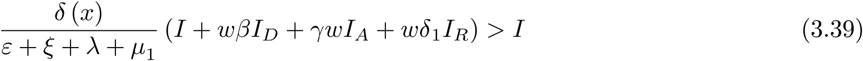

noting that

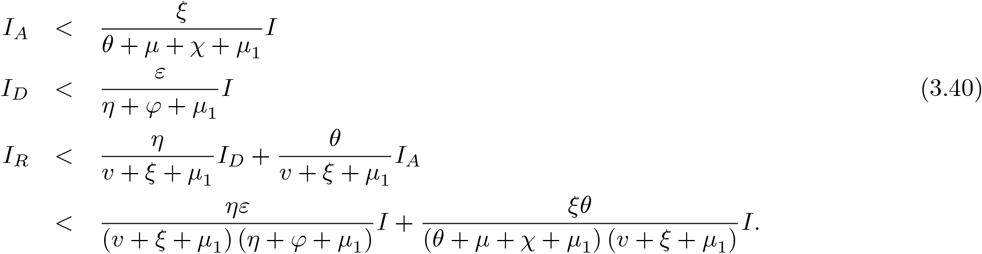

Also

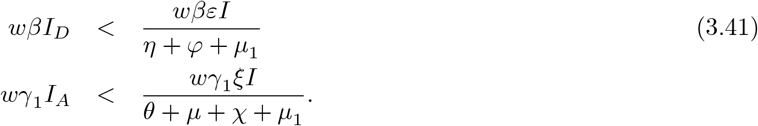

Therefore we have the following inequality in terms of *I*

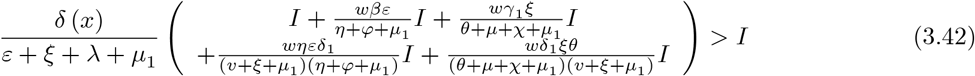

and

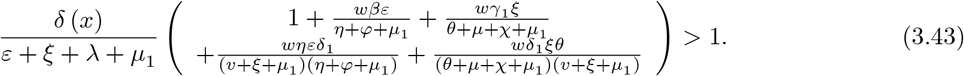

Therefore

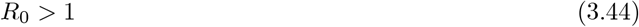

Where

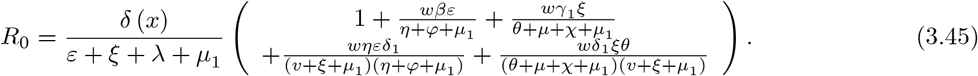

This shows that we have a unique endemic equilibrium when *R*_0_ > 1.

### 3.2 Local and global stability of the Disease-free equilibrium

#### Lemma 1

The disease-free equilibrium *E*_0_ of the COVID-19 system is locally asymptotically stable when *R*_0_ < 1 and unstable when *R*_0_ > 1. The Jacobian matrix for COVID-19 system is given by

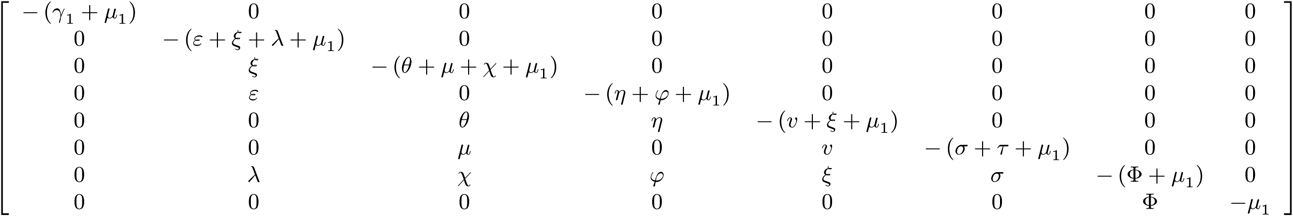

It is known that the disease-free equilibrium *E*_0_ asymptotically stable if and only if the *tr* (*J* (*E*_0_)) < 0 and the det (*J* (*E*_0_)) > 0. For the suggested COVID-19 the trace of *J* (*E*_0_) is

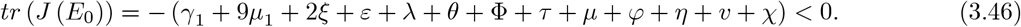

The determinant of *J* (*E*_0_) is

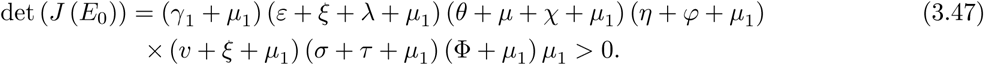

In this case, we can conclude that the disease-free equilibrium of the suggested model for COVID-19 under vaccination and treatment is locally asymptotically stable.

#### Theorem 2

The COVID-19 model disease-free equilibrium is globally asymptotically stable within the feasible interval if *R*_0_ < 1 and unstable if *R*_0_ > 1.

##### Proof

We use the Lyapunov function defined by

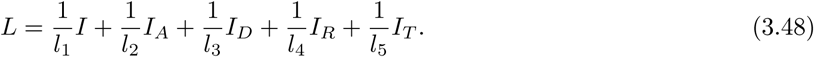

Therefore its derivative along the solutions of the COVID-19 model

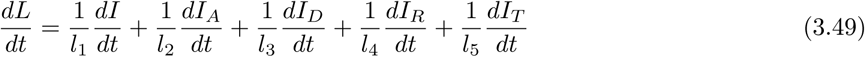

where

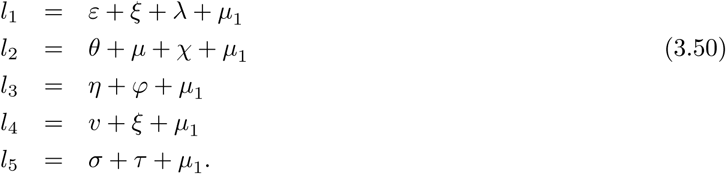

Then we write

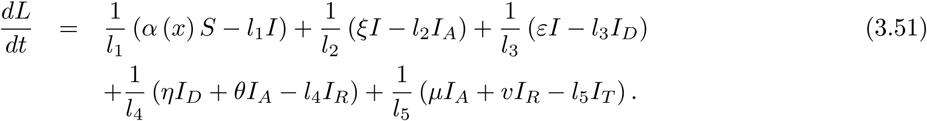

We have on the other hand that

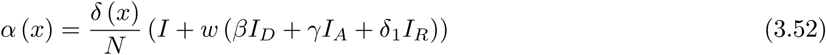

and we get

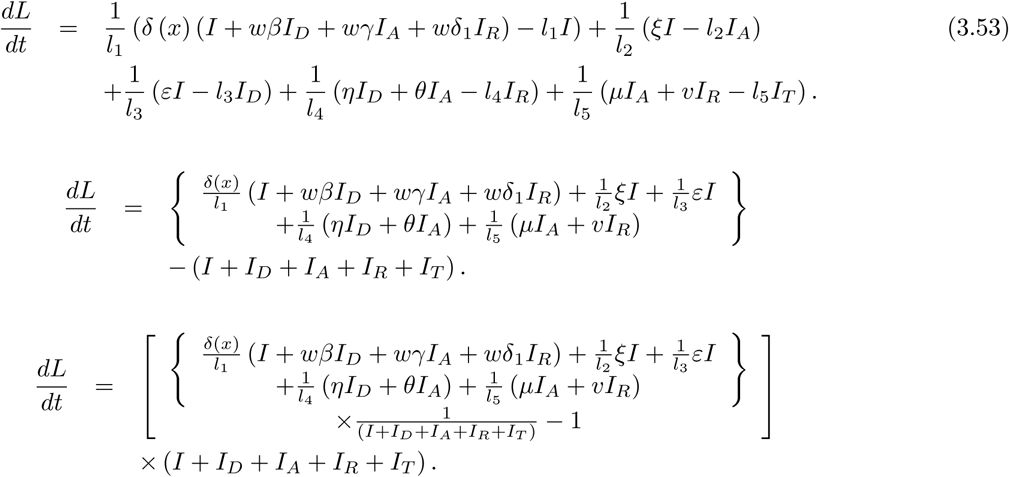

We now divide by *I* to obtain

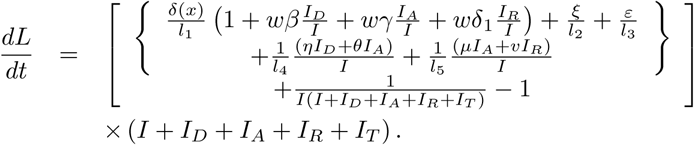

However, since *I* is greater than *I_D_*, *I_A_*, *I_R_*, *I_T_* classes, thus

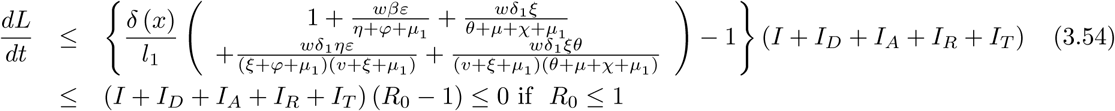

since *I* + *I_D_* + *I_A_* + *I_R_* + *I_T_* > 0, ∀*t*- Therefore the COVID-19 will be eliminated according to the suggested model if and only if *R*_0_ < 1- In particular, since all parameters in the COVID-19 model are positive thus then Lyapunov decreases 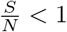 if *R*_0_ < 1 and increases if *R*_0_ > 1, finally *L* = 0 if

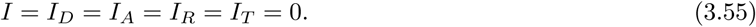

Therefore *L* is Lyapunov function within the feasible biological interval and the bigger compact invariant set in 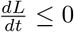 is the point *E*_0_. By the well-known Lasalle’s invariance concept [8] each solution of the COVID-19 model suggested in this work with initial condition in Ω leads to *E*_0_ when *t* → ∞ only if *R*_0_ < 1. Conclusion, the disease-free equilibrium *E*_0_ of the COVID-19 model suggested here which includes treatment and vaccination is globally asymptotically stable.

### 3.3 Local and global stability of the endemic equilibrium

We compute first the Jacobian matrix of the COVID-19 model for endemic equilibrium case

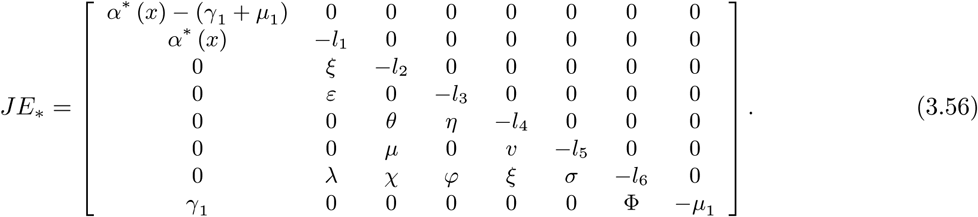

We now construct a characteristic equation associate to this model

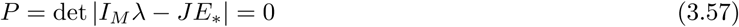

where *I_M_* is the 8 × 8 unit matrix. Then we have

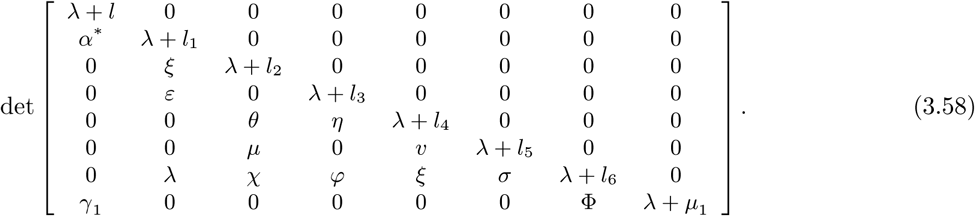

From the above, we obtain the following characteristic

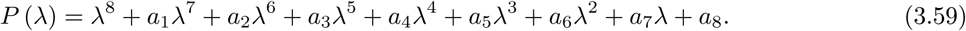

The square Hurwitz matrix associate to the above polynomial *P* (λ) is given as

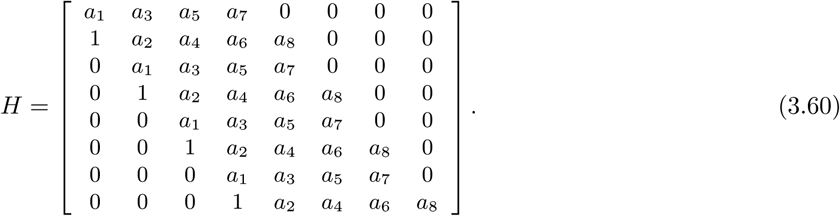

Then we have

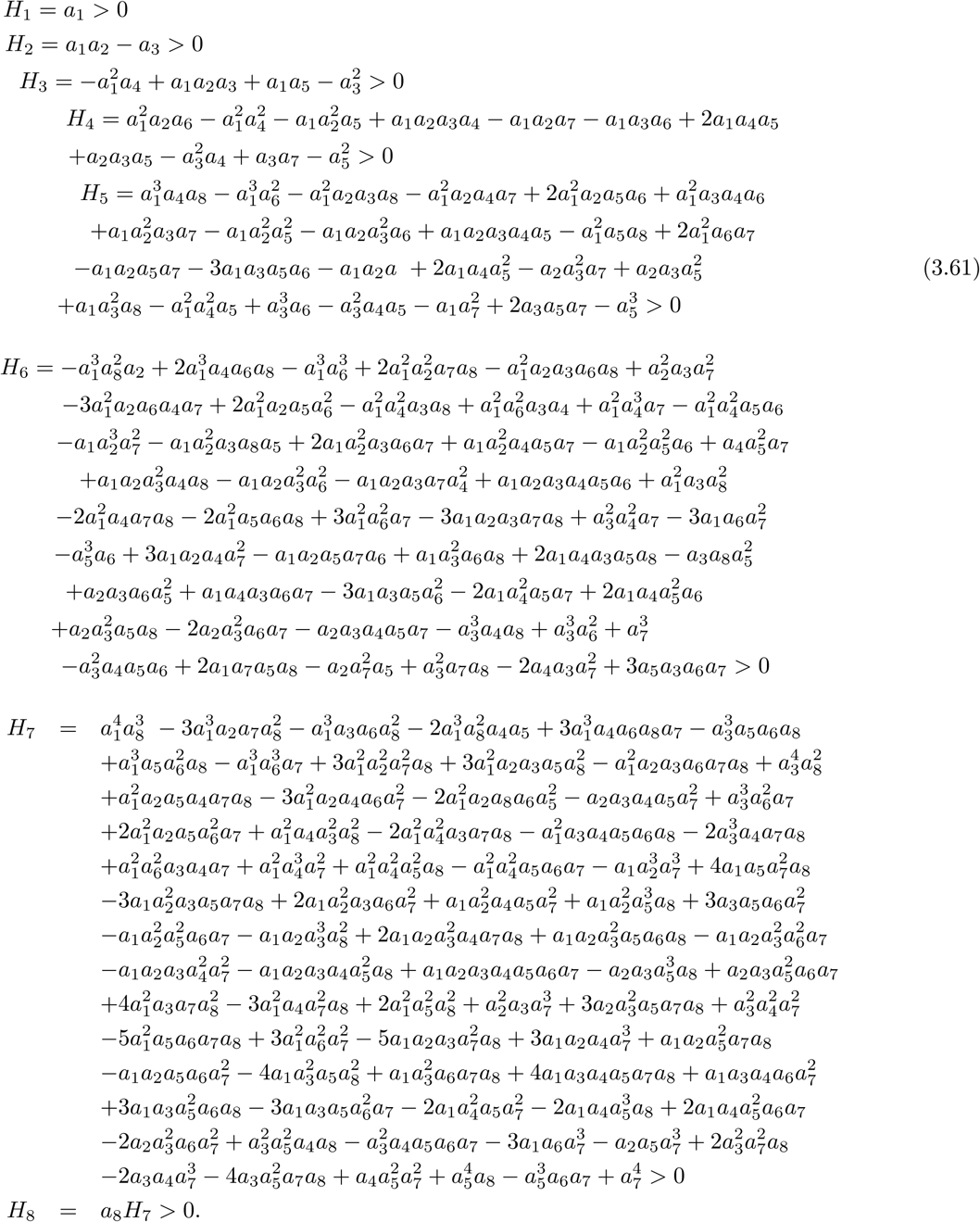

#### Theorem 3

If *R*_0_ > 1, the endemic equilibrium point *E*_*_ of the COVID-19 system is globally asymptotically stable.

##### Proof

We prove this using the Lyapunov function

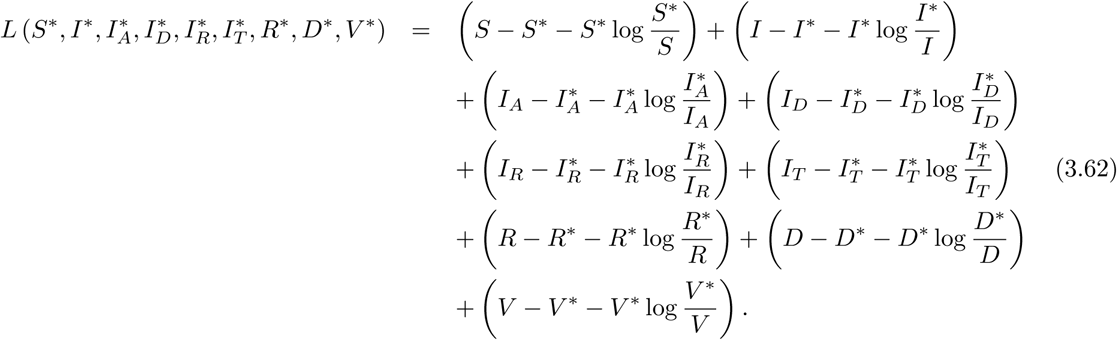

Therefore taking the derivative respect to *t* on both sides gives

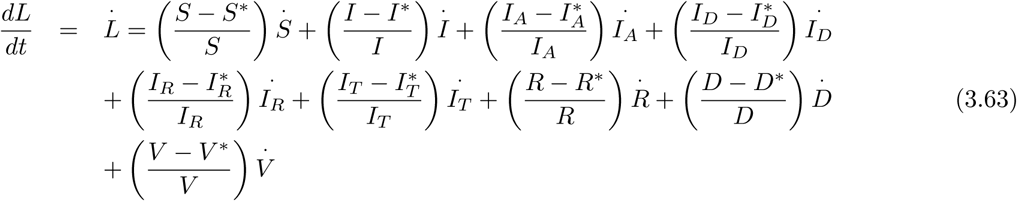

replacing 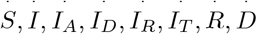 and 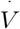 by their values, we obtain

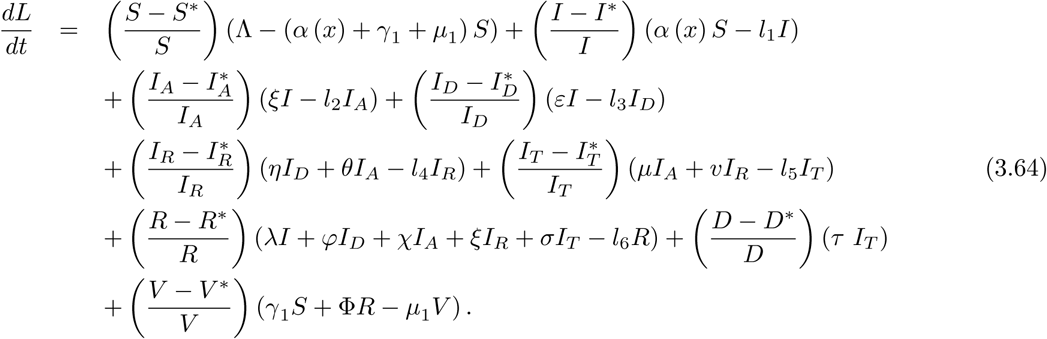

Then we have

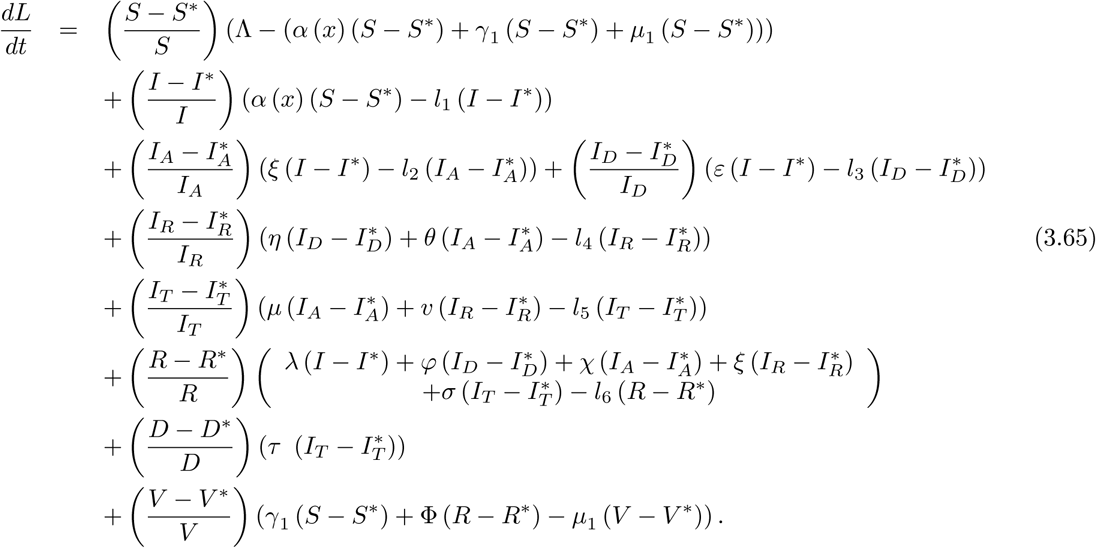

They can be separated in two part as follows

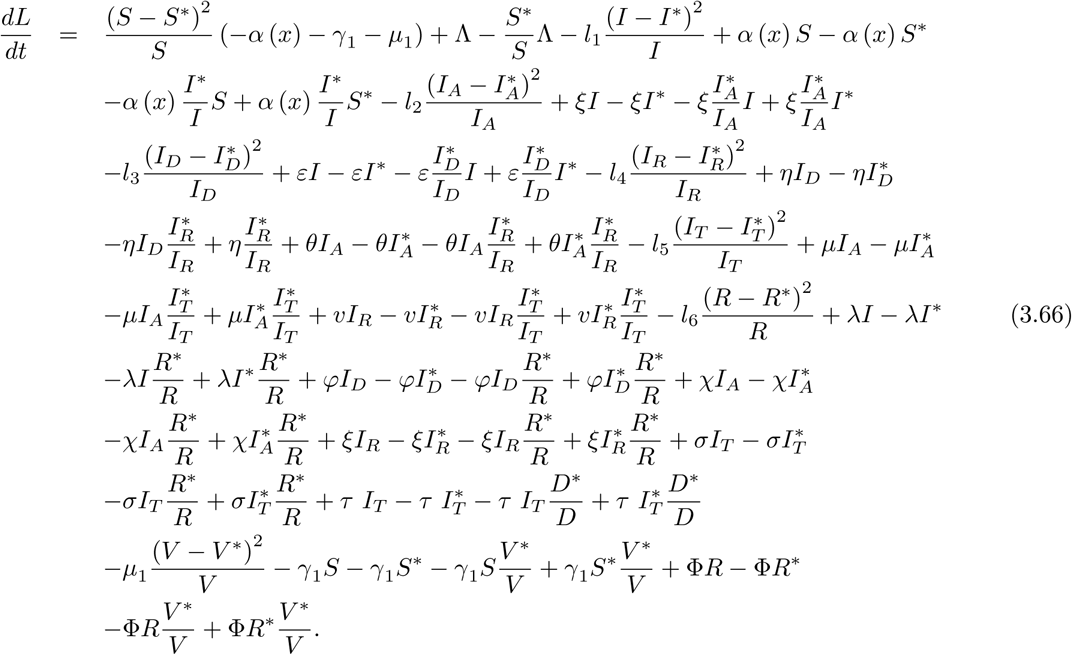

This can be simplified as

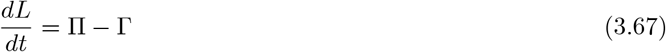

where

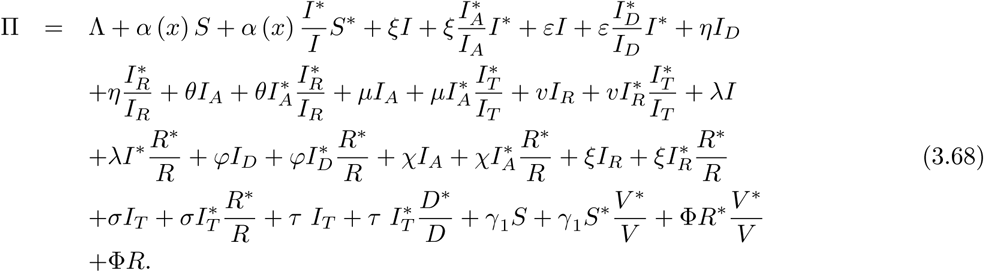

and

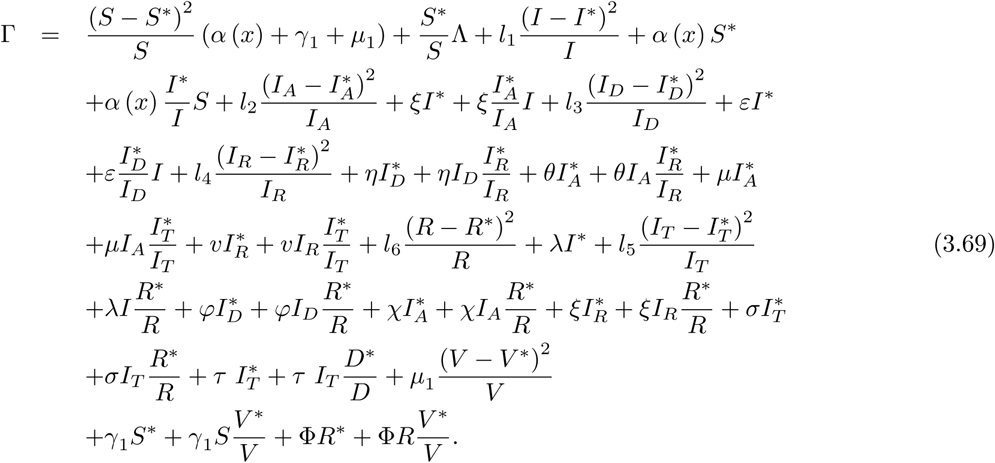

Therefore having П < Г, this implies 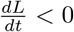, however

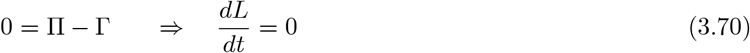

if

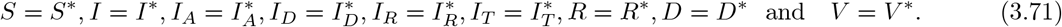

We can now conclude that the largest compact invariant set for COVID-19 model in

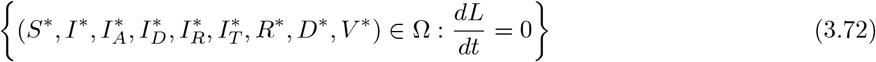

is the point {*E*_*_} the endemic equilibrium of the COVID-19 model. Therefore using the Lasalle’s invariance principle, we conclude that *E*_*_ is globally asymptotically stable in Ω if П < Г.

## 4 Modeling with non-local operators

Due to complexities around the spread of COVID-19, it is really hard to produce predictions. Especially, when multi-scenarios are requested. Indeed it has been reported that local operators including can not provide nonlocal processes for example change in processes. In this section, we present an analysis of COVID-19 model with local operators including Caputo-Caputo-Fabrizio, Atangana-Baleanu and the new introduced fractal-fractional operators. We first present the definition of each operator. We start with the definition of Caputo fractional derivative

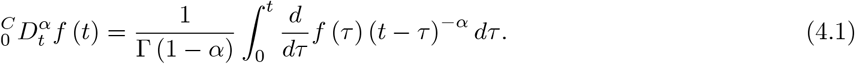

Caputo-Fabrizio fractional derivative

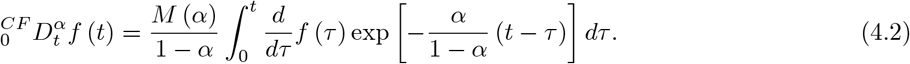

Atangana-Baleanu fractional derivative

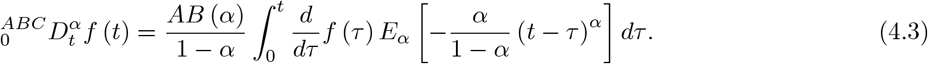

The fractal-fractional derivative with power-law kernel

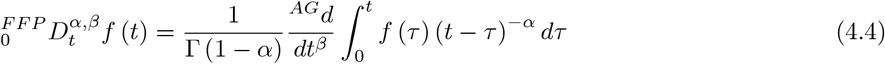

where

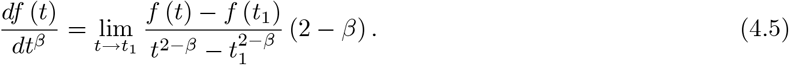

The fractal-fractional derivative with exponential decay kernel

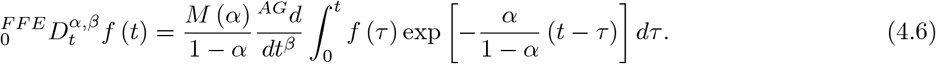

The fractal-fractional derivative with Mittag-Leffler kernel

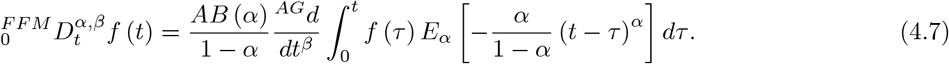

The associated integral operators of the last three operators are given as

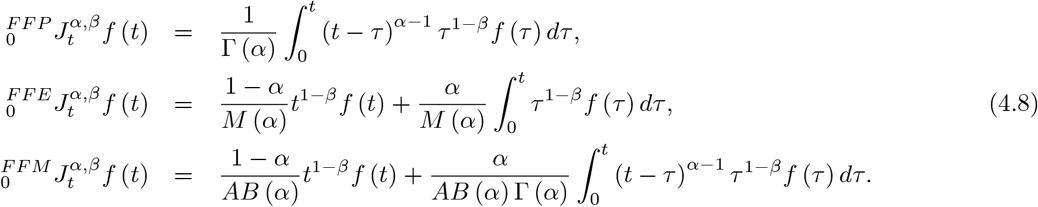

### 4.1 Positive solutions with non-local operators

In this subsection, we present a detailed analysis of positiveness of the solutions for COVID-19 model with non-local operators. We start with ABC derivative case

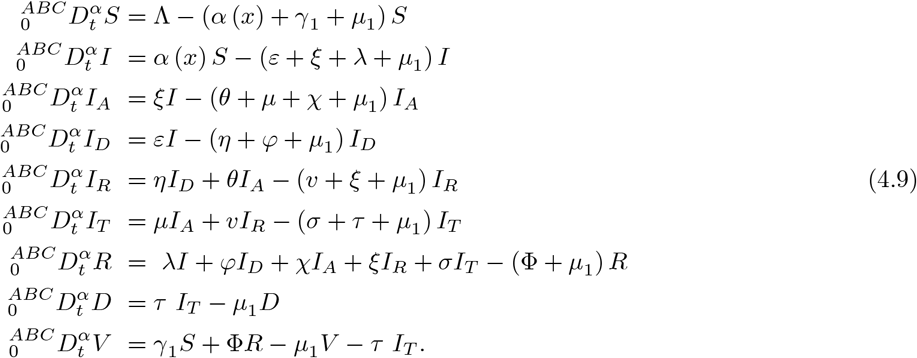

The norm and all hypothesis of the classical are valid here also

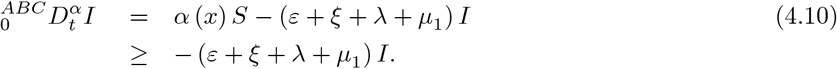

This produces

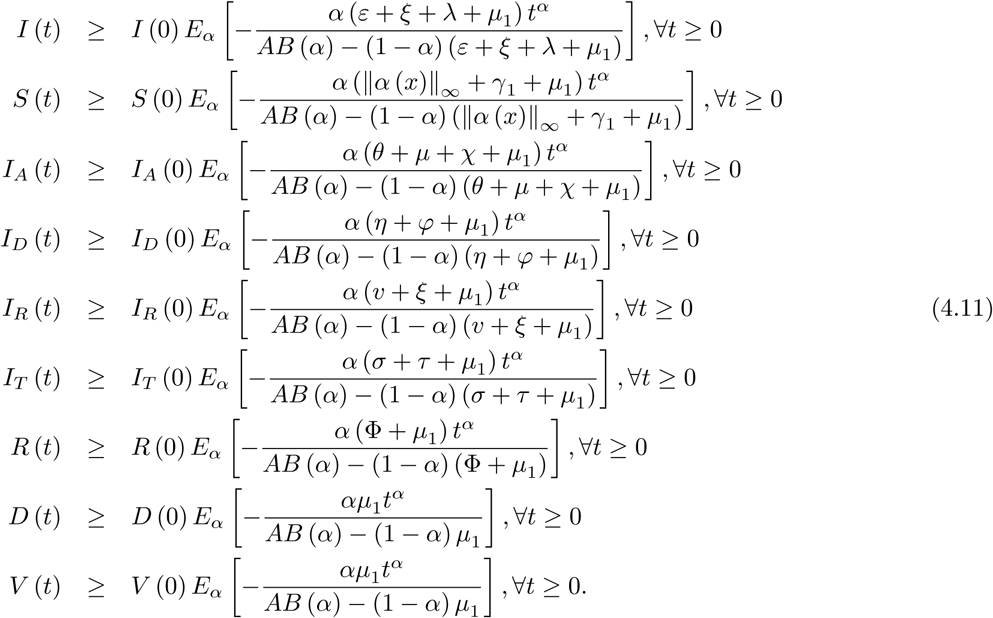

This shows that if all the initial conditions are positive then all solutions are positive with the Atangana-Baleanu derivative. With Caputo-Fabrizio, we have

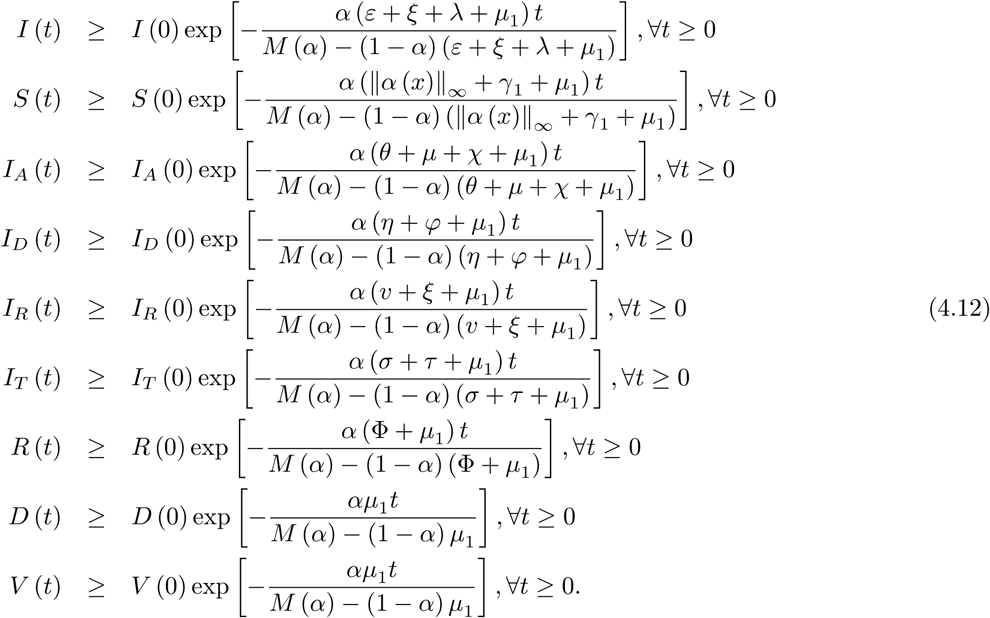

This shows that all solutions are positive if all the initial conditions are positive with Caputo-Fabrizio. With Caputo derivative, we have

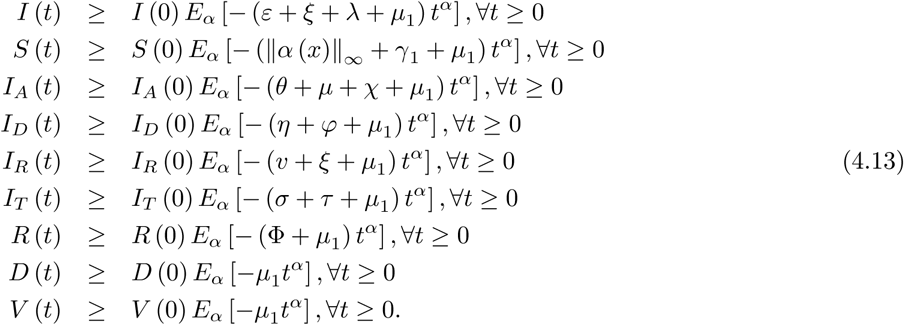

This shows that all solutions are positive if all the initial conditions are positive with Caputo.

For fractal-fractional case, without loss of generality, we present the proof for *I* class and the rest can be deduced similarly. We start with power-law case

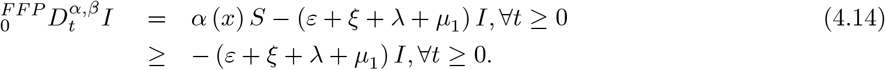

and

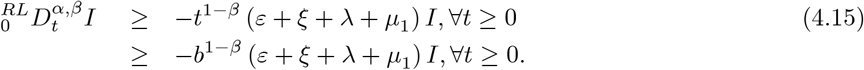

Thus, we have

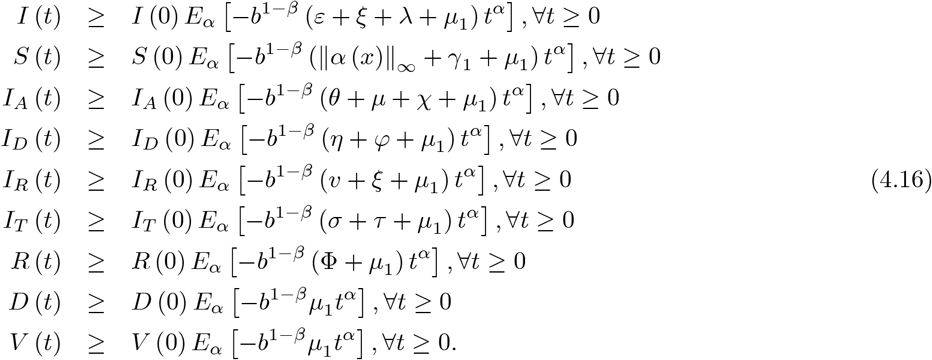

With exponential kernel, we have

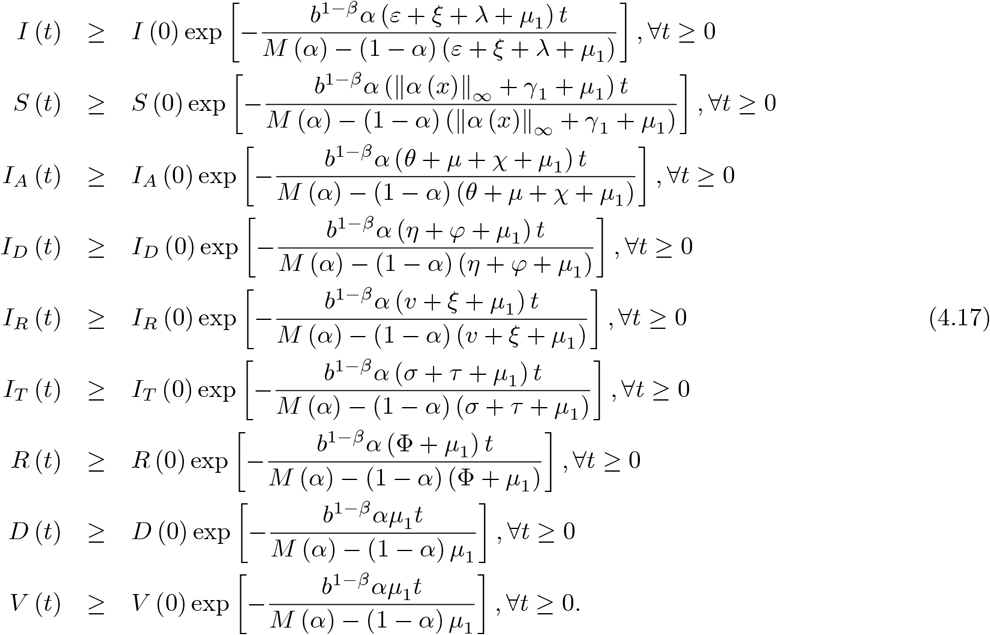

With Mittag-Leffler kernel, we obtain

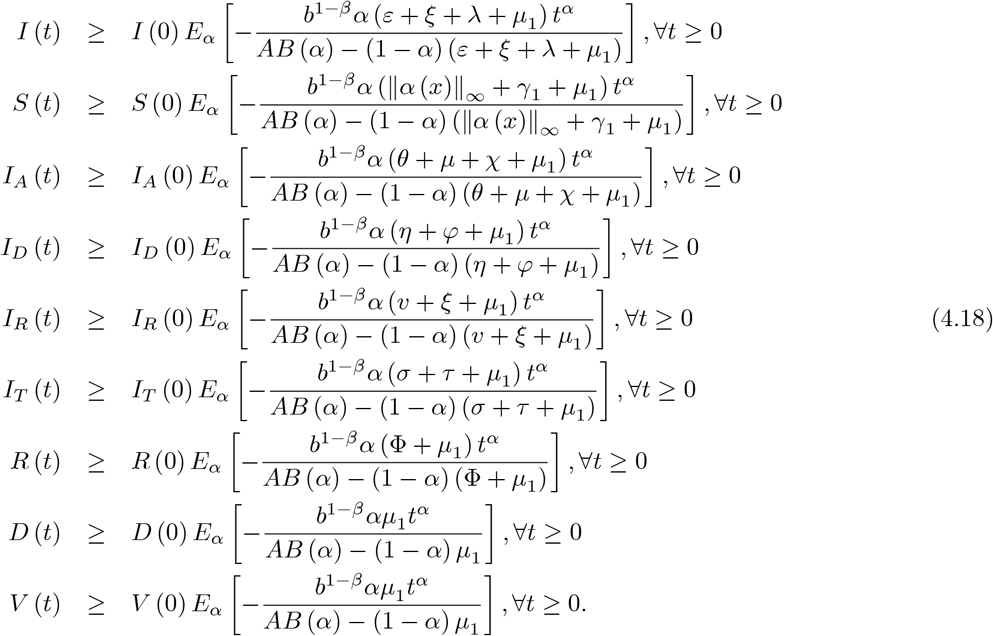

## 5 Numerical analysis of COVID-19 models from classical to nonlocal operators: Application of Atangana-Seda numerical scheme

While analytical methods are adequate to provide exact solution of a giving equation, or systems of equations, it is important to note that when dealing with nonlinear equations, analytical methods cannot be used. In particular, the model of COVID-19 suggested in this work either with classical or non-local operators contains nonlinear components therefore analytical methods is ineffective. Very recently, Atangana and Seda [10] made use of Newton polynomial to introduce an alternative numerical scheme that can be used to solving nonlinear equations arising in many field of science, technology and engineering, the method has been recognized to be very efficient and accurate. In this section, we will make use of the Atangana-Seda scheme to solve the suggested mathematical model for COVID-19 for different differential operators.

We start with classical case for numerical solution of COVID-19 model

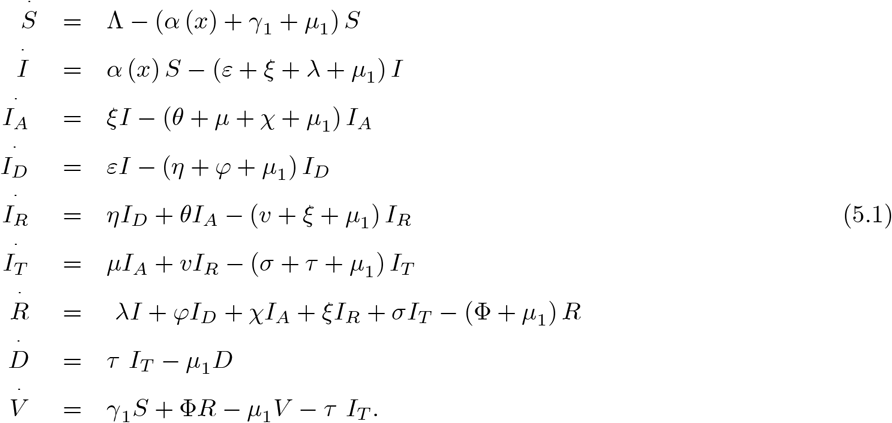

For simplicity, we write above equation as follows;

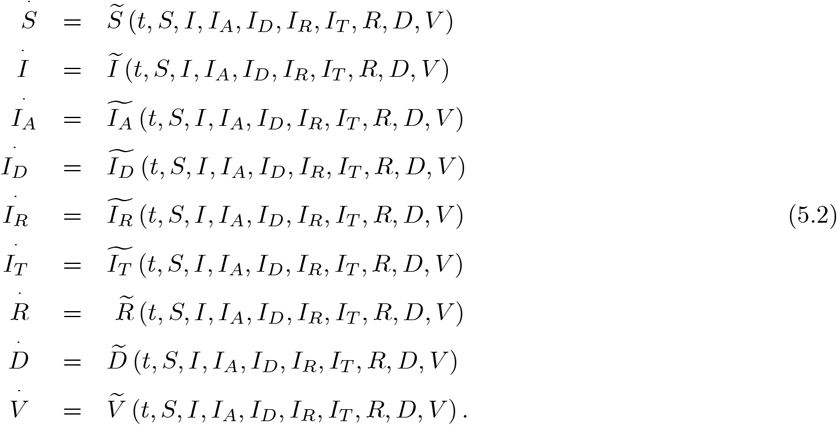

where

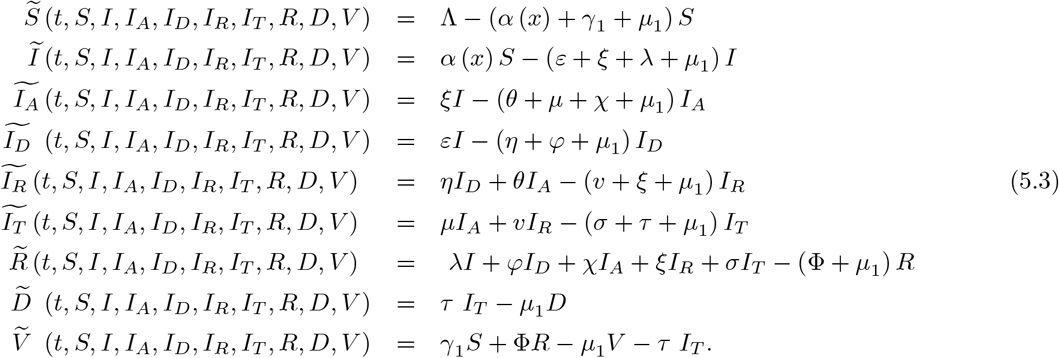

After applying fractal-fractional integral with exponential kernel, we have the following

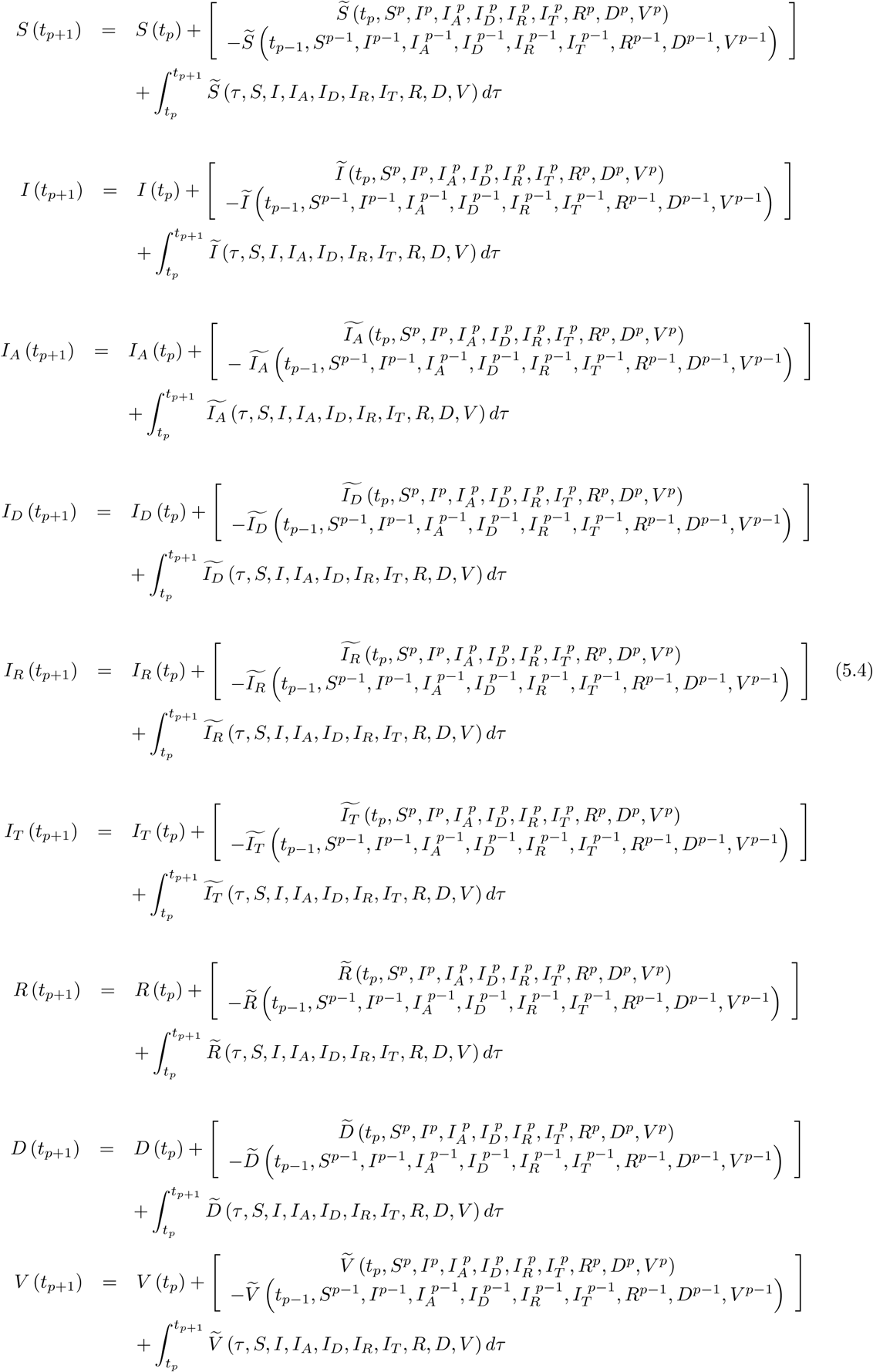

We can have the following scheme for this model

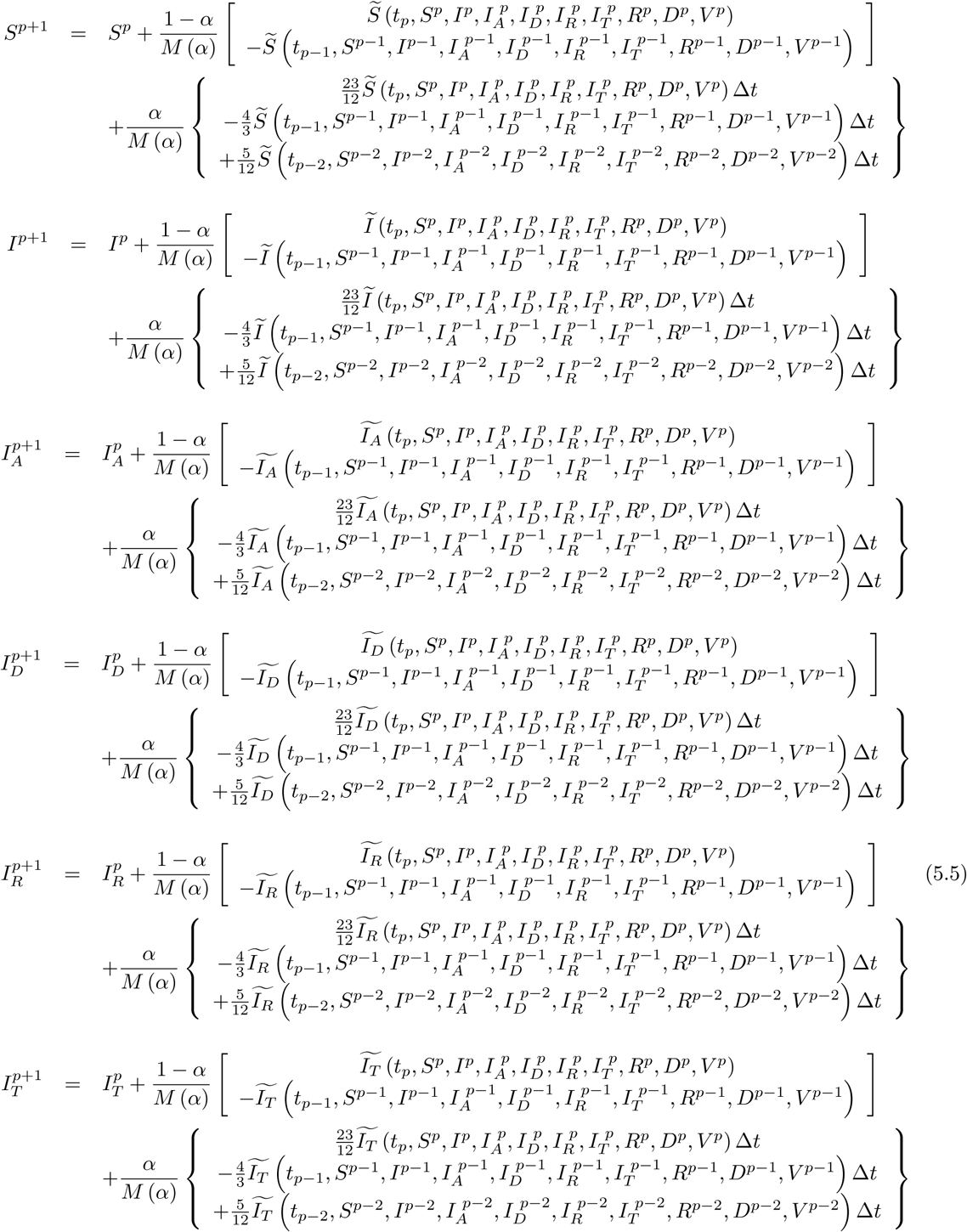

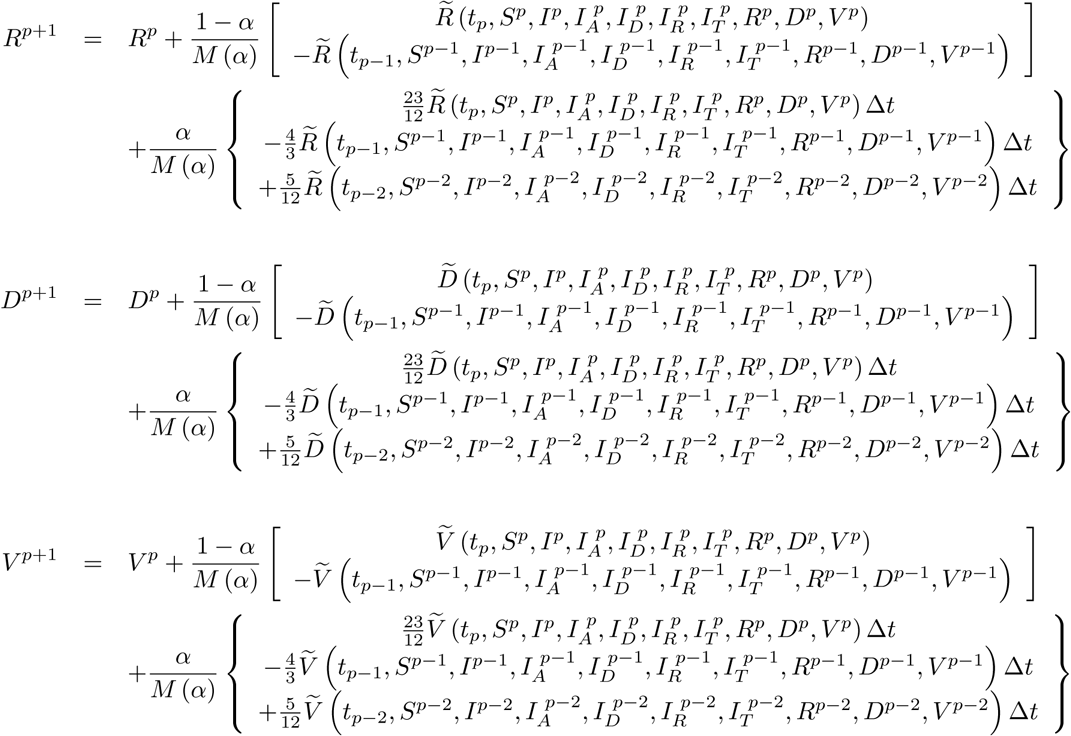

Now, we handle the following model with classical derivative

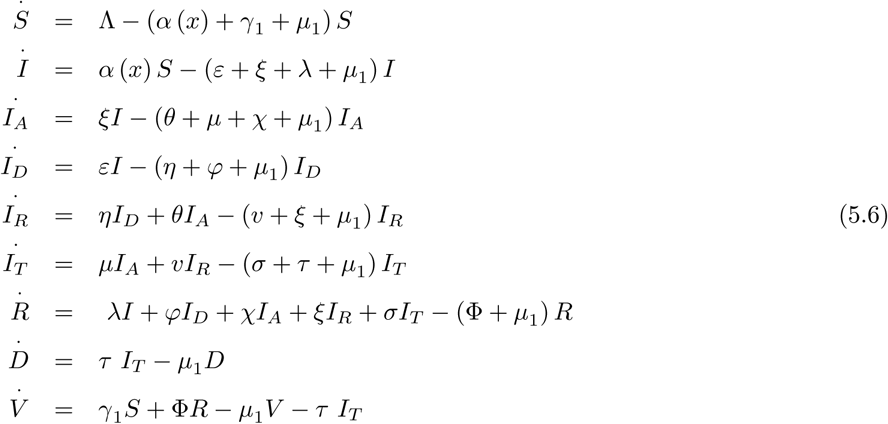

where initial conditions are

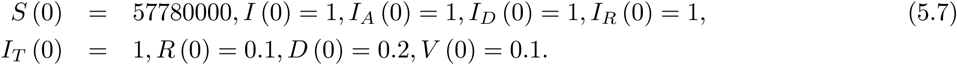

Also the parameters are chosen as

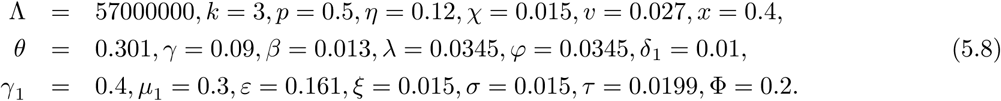

We present numerical simulation for COVID-19 model in Figure 33 and 34.

**Figure 33.**
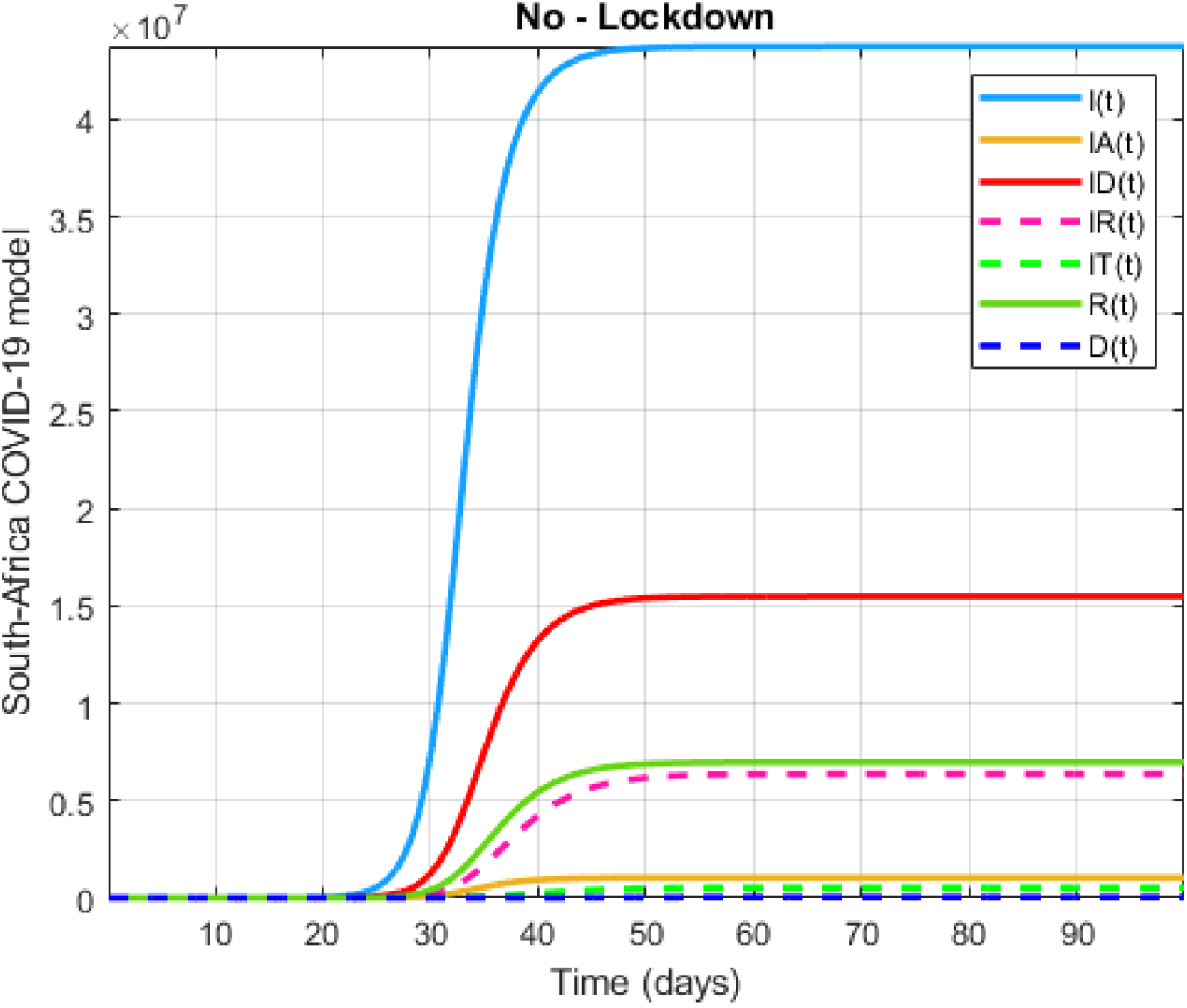
Numerical visualization for COVID-19 model in South Africa.

**Figure 34.**
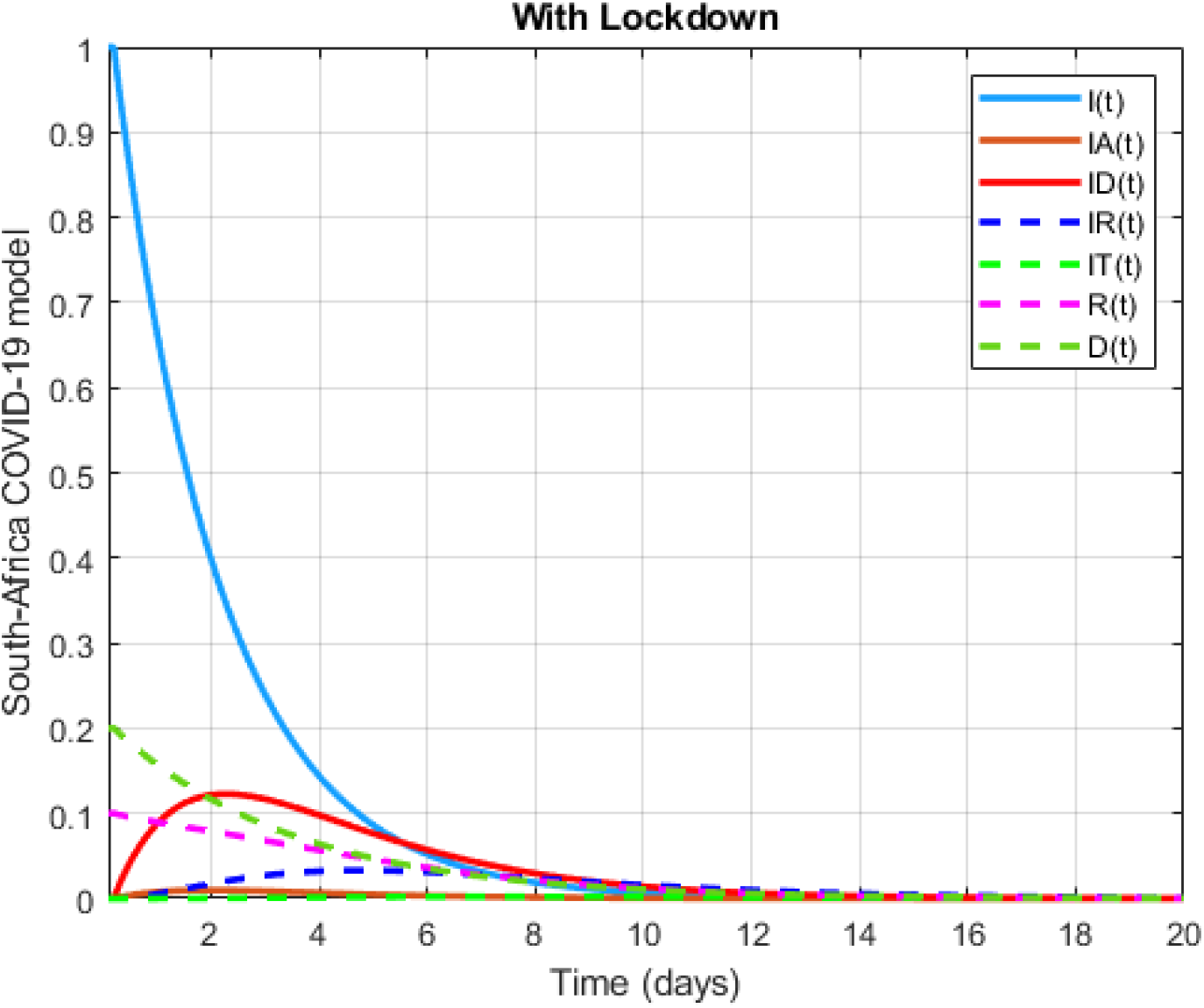
Numerical visualization for COVID-19 model in South Africa.

For same model, initial conditions are chosen

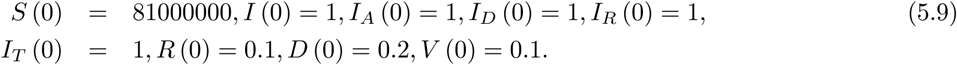

Also the parameters are

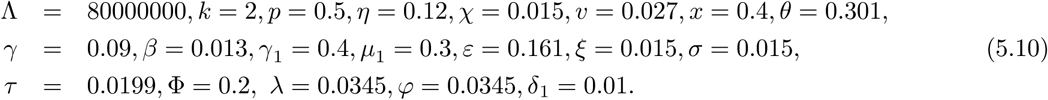

We present numerical simulation for COVID-19 model in Figure 35 and 36.

**Figure 35.**
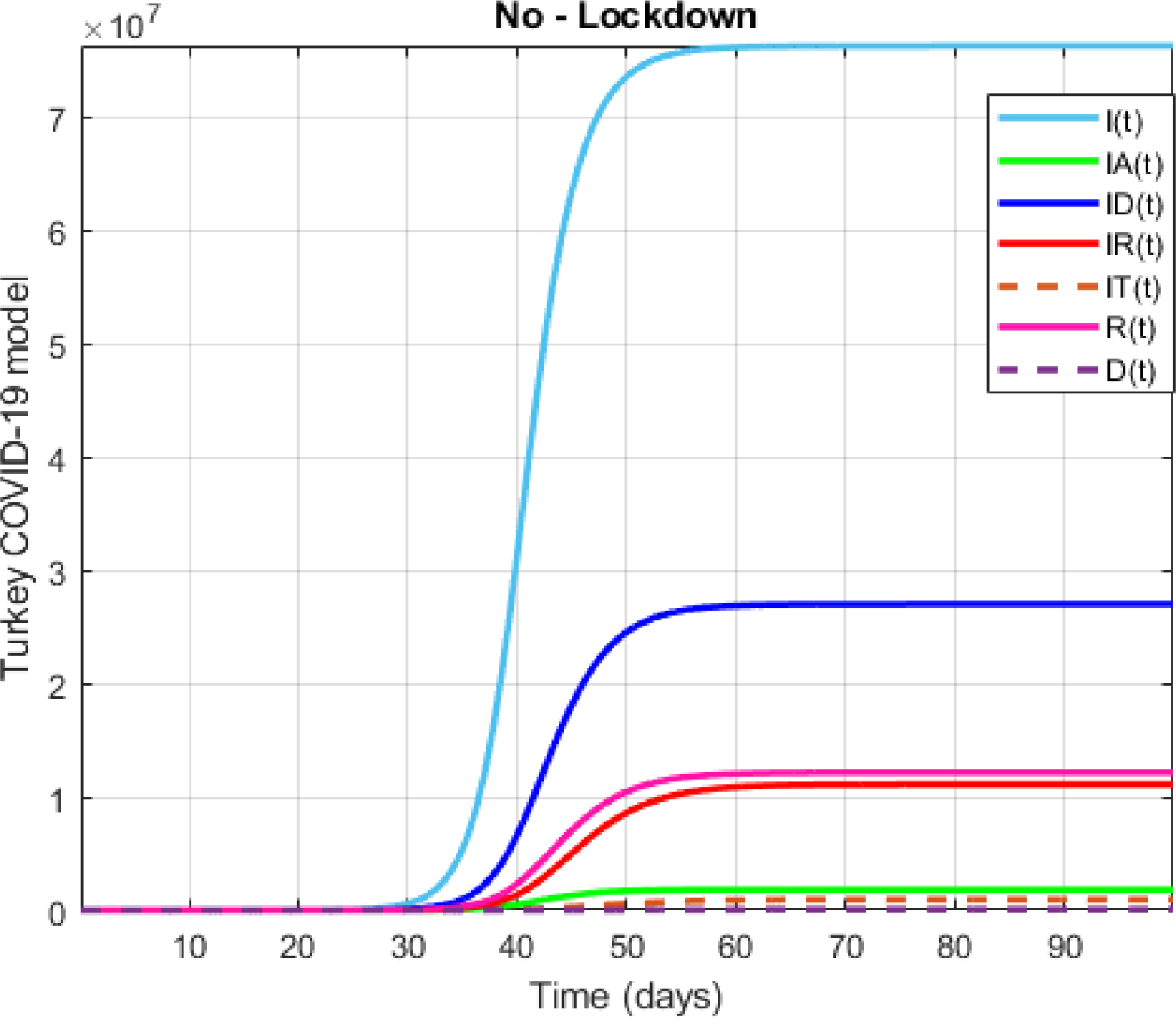
Numerical visualization for COVID-19 model in Turkey.

**Figure 36.**
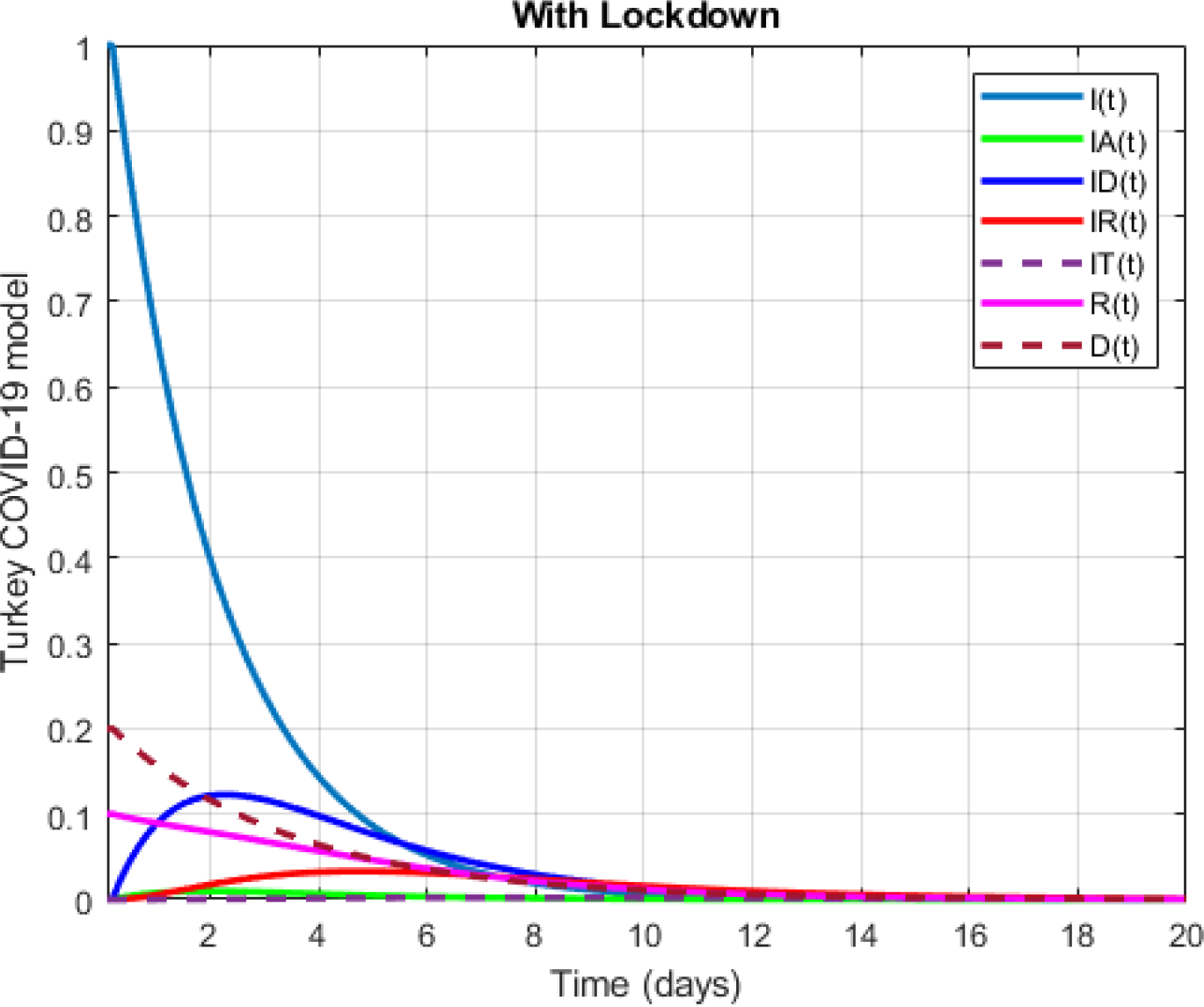
Numerical visualization for COVID-19 model in Turkey.

Now, we replace the classical differential operator will be replaced by the operator with power-law, exponential decay and Mittag-Leffler kernels. We start with exponential decay kernel.

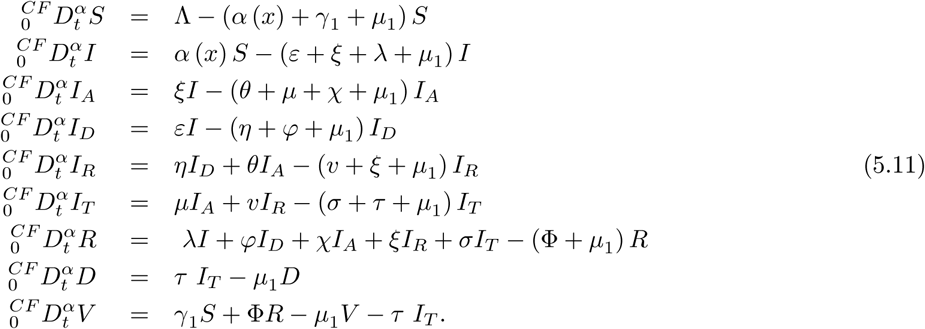

For simplicity, we write above equation as follows;

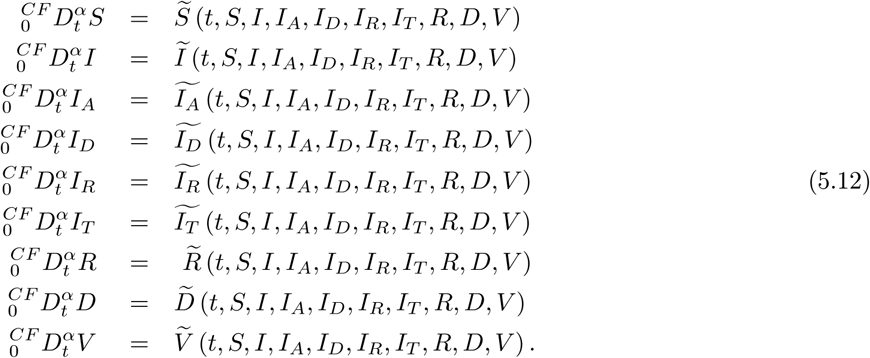

After applying fractal-fractional integral with exponential kernel, we have the following

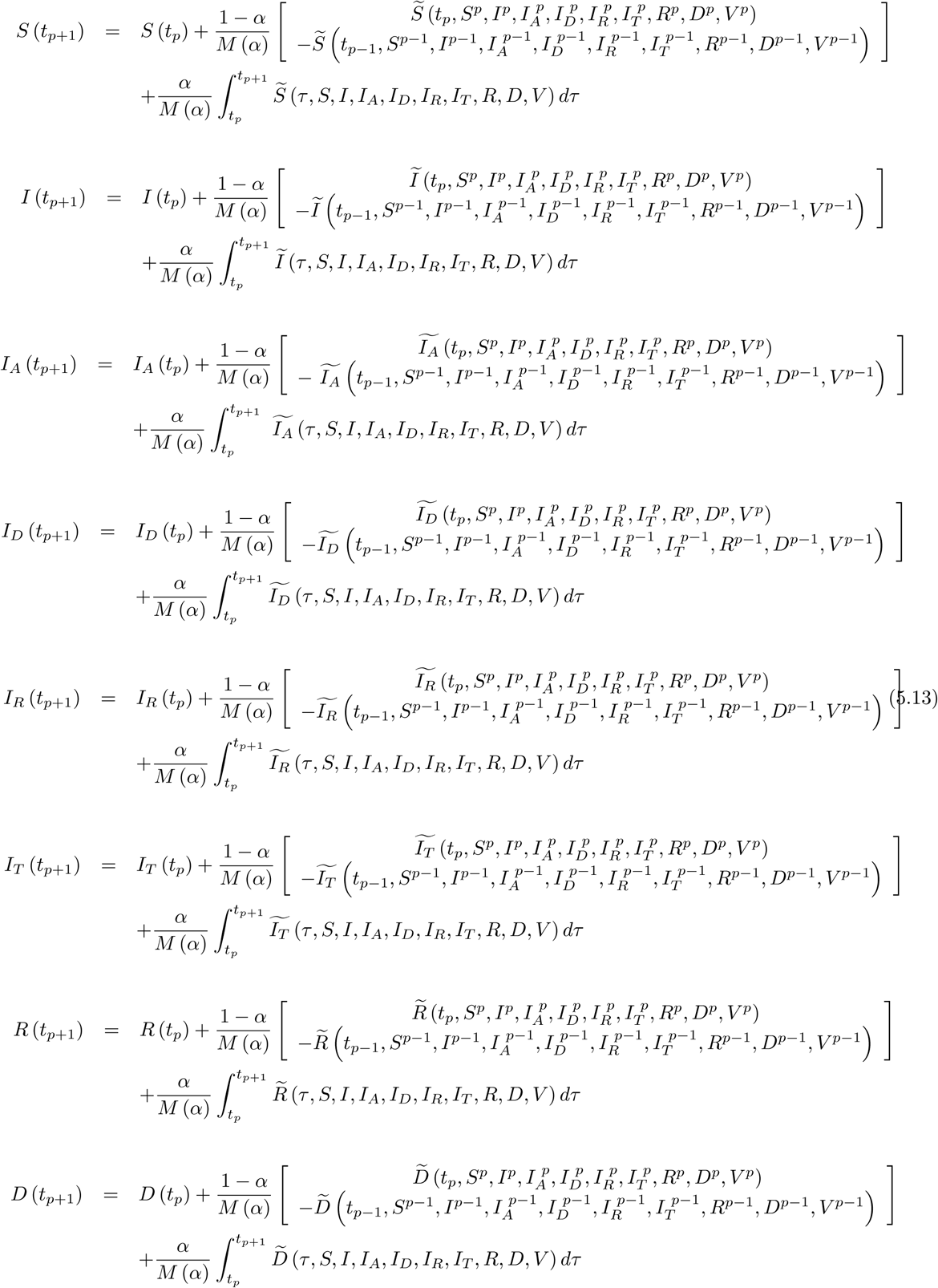

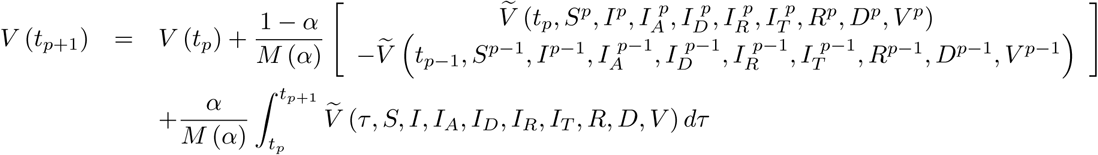

We can have the following scheme for this model

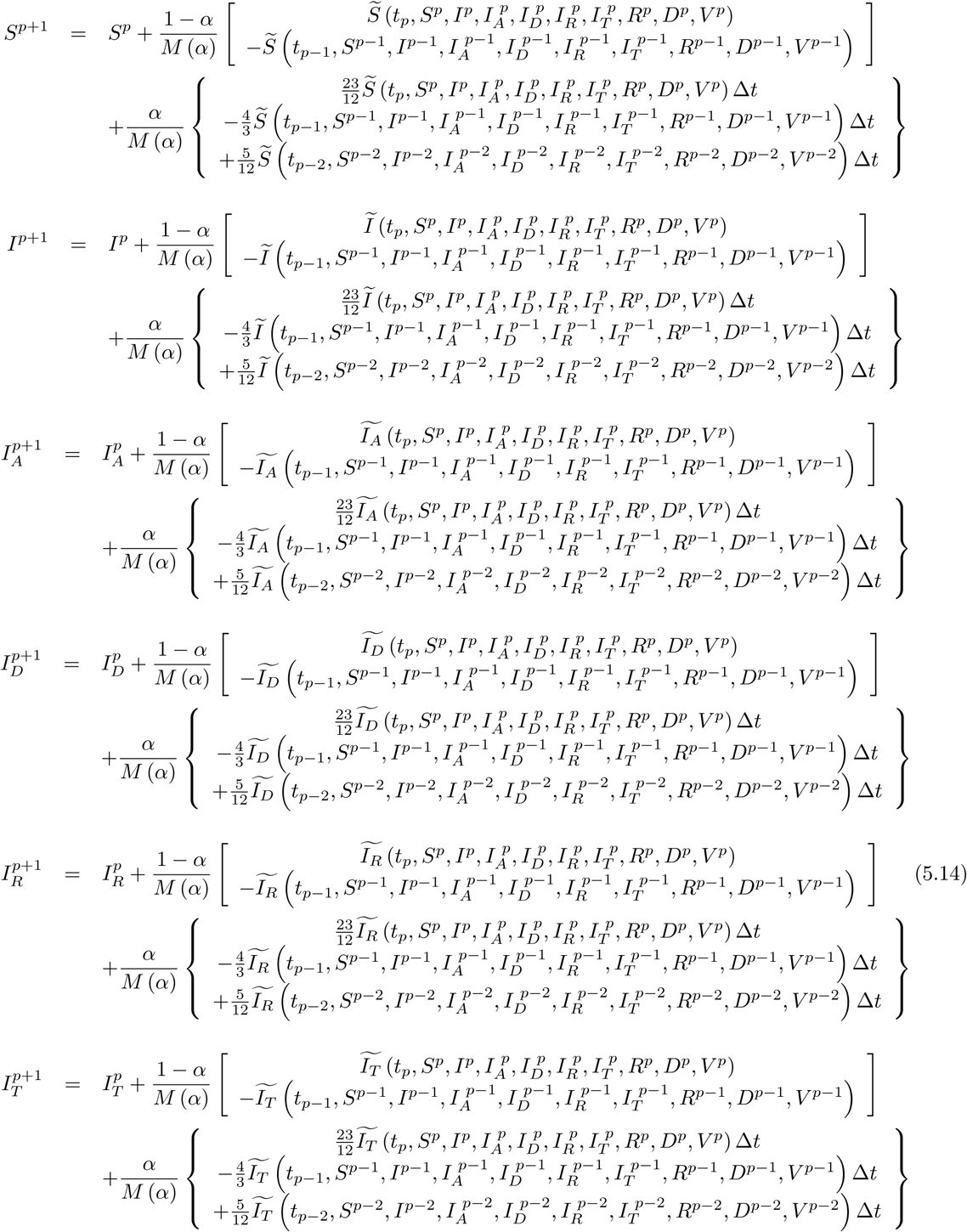

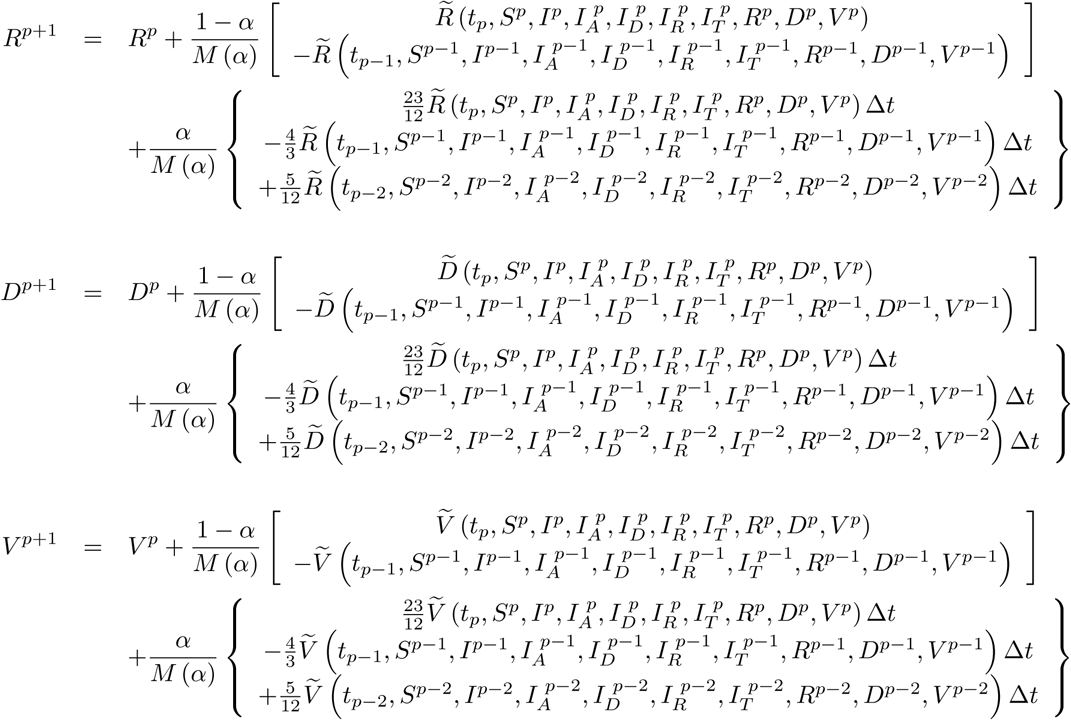

For Mittag-Leffler kernel, we can have the following

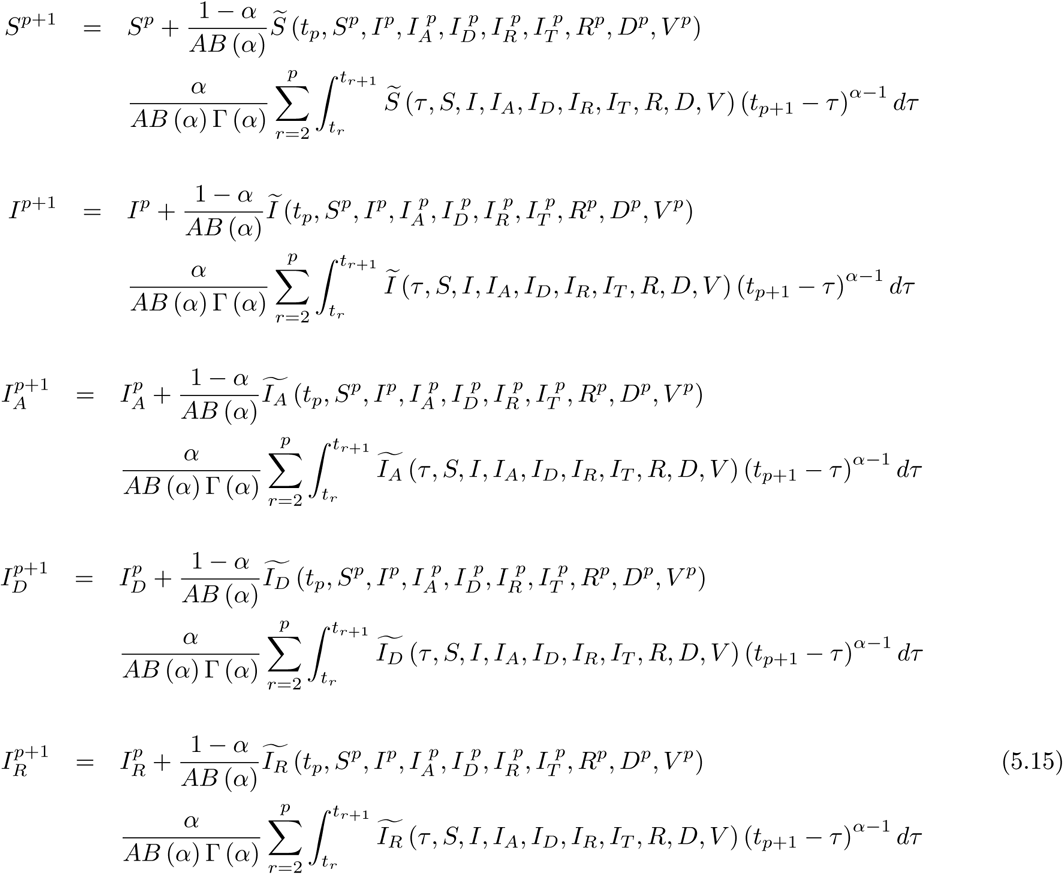

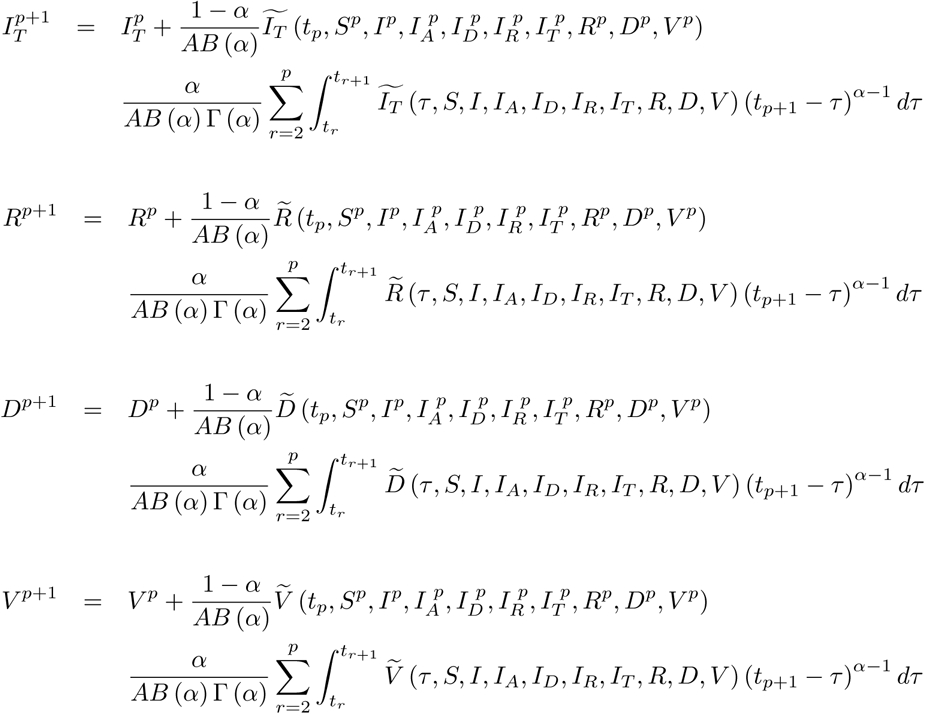

We can get the following numerical scheme

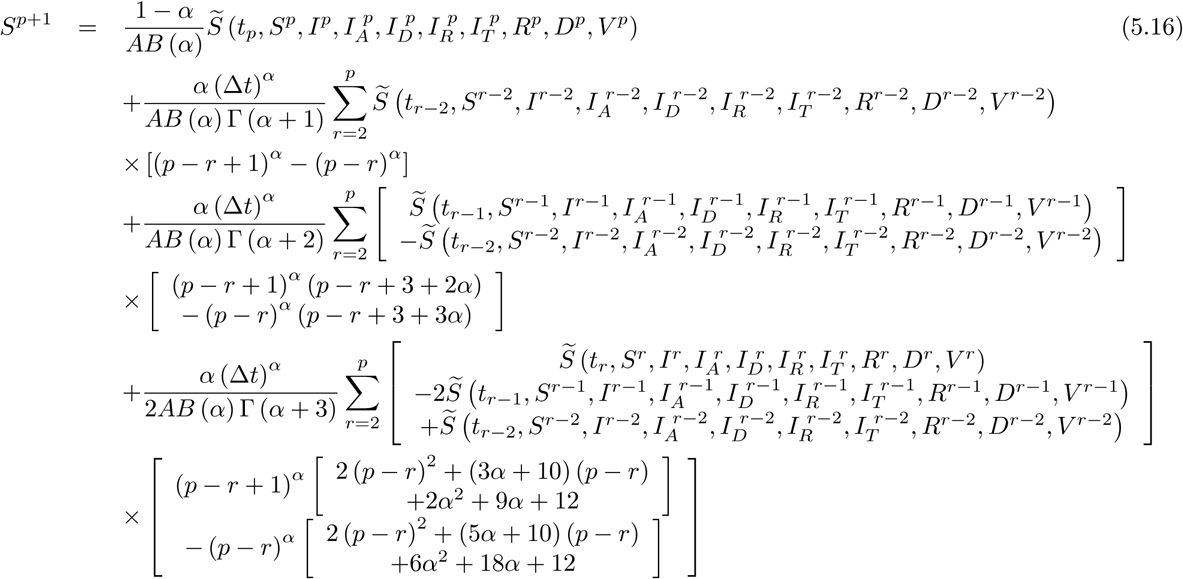

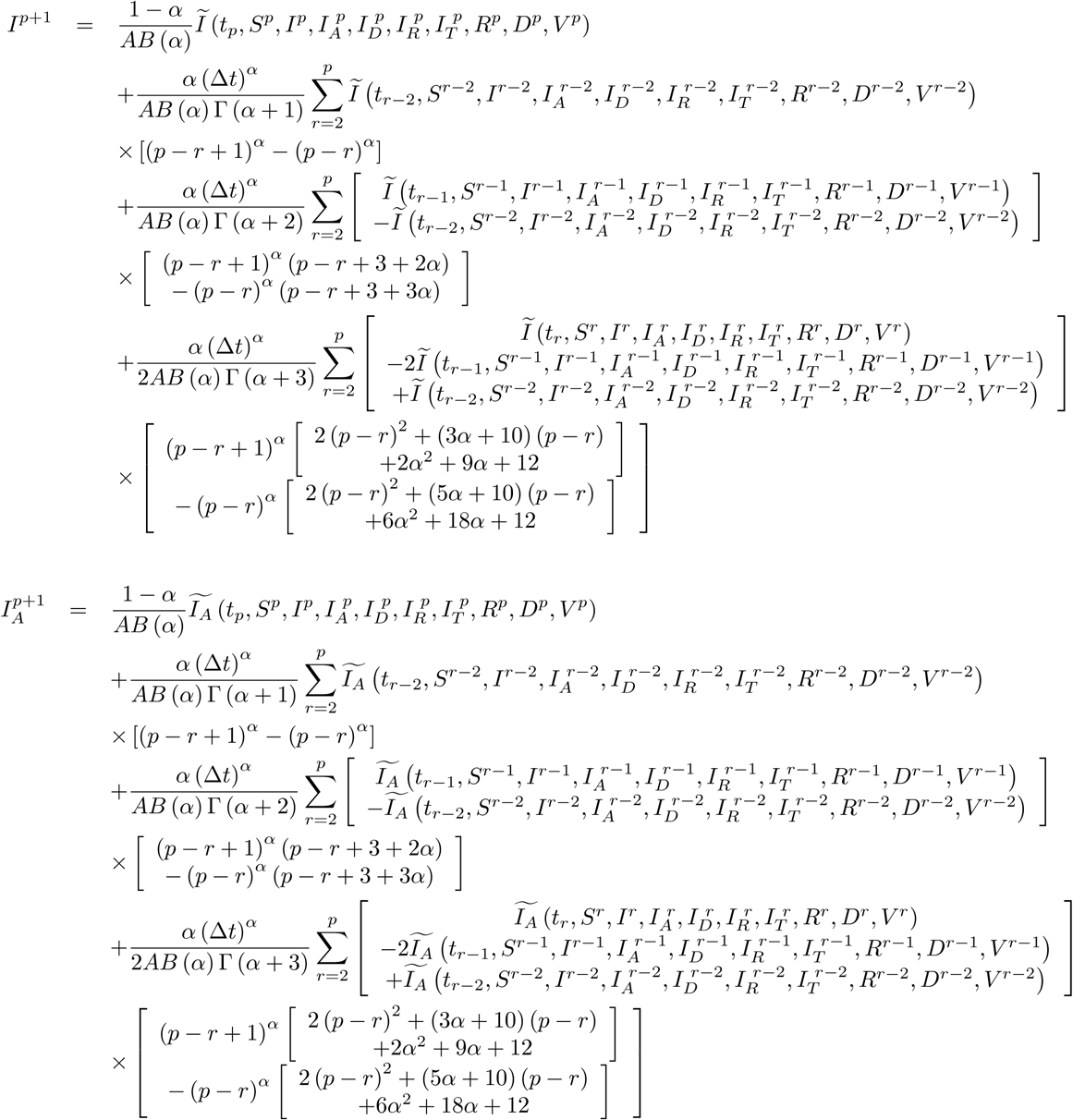

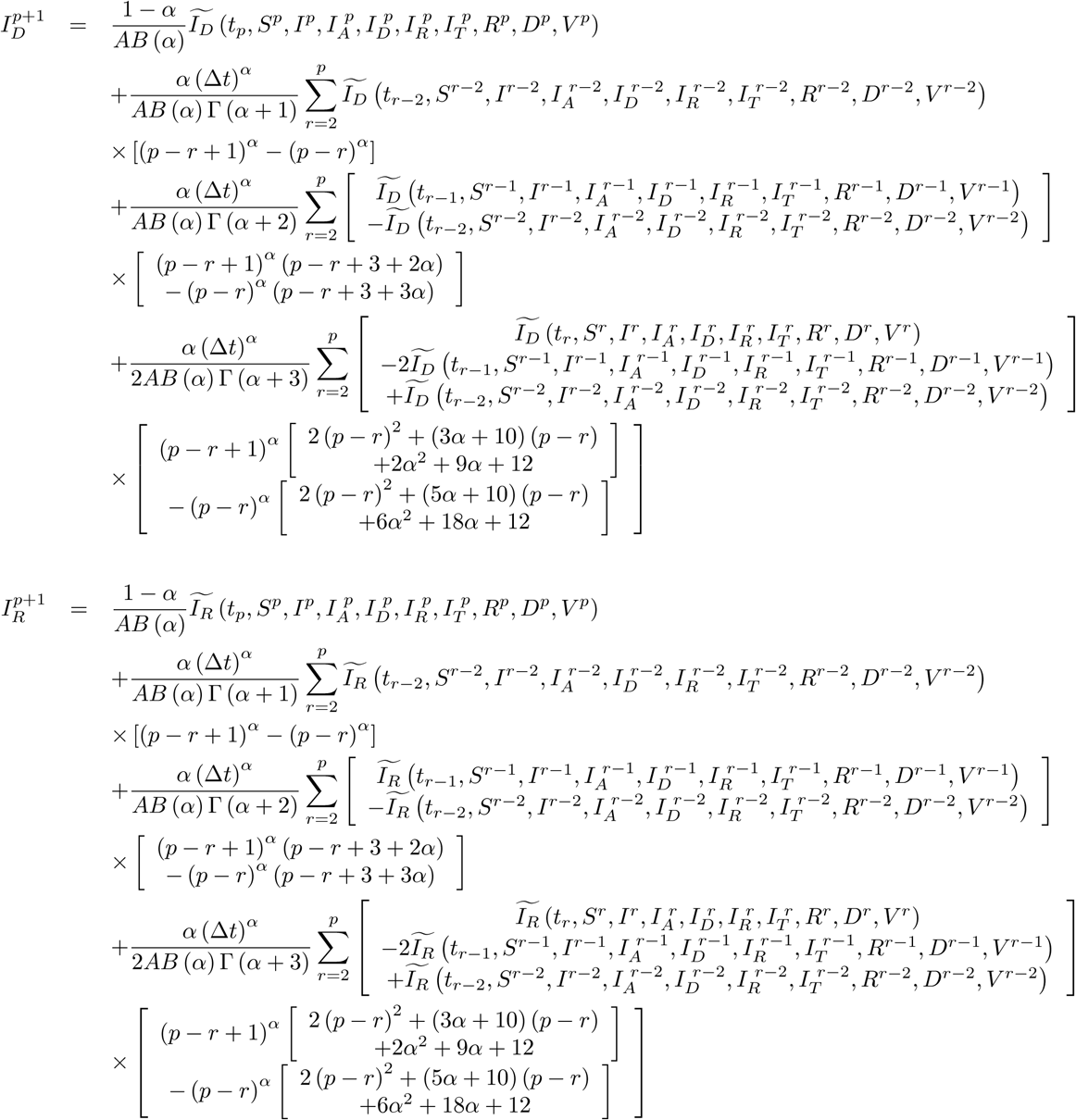

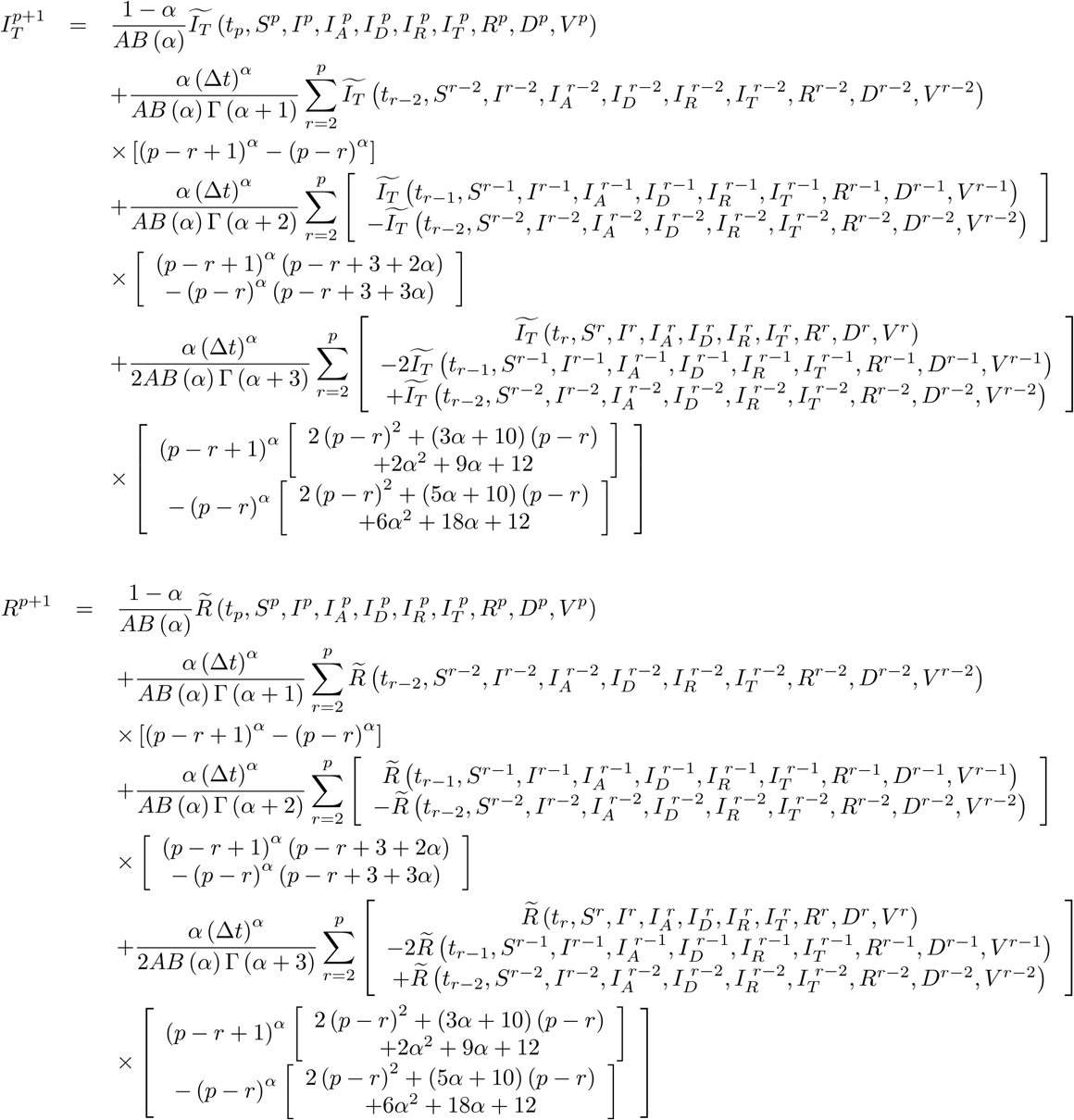

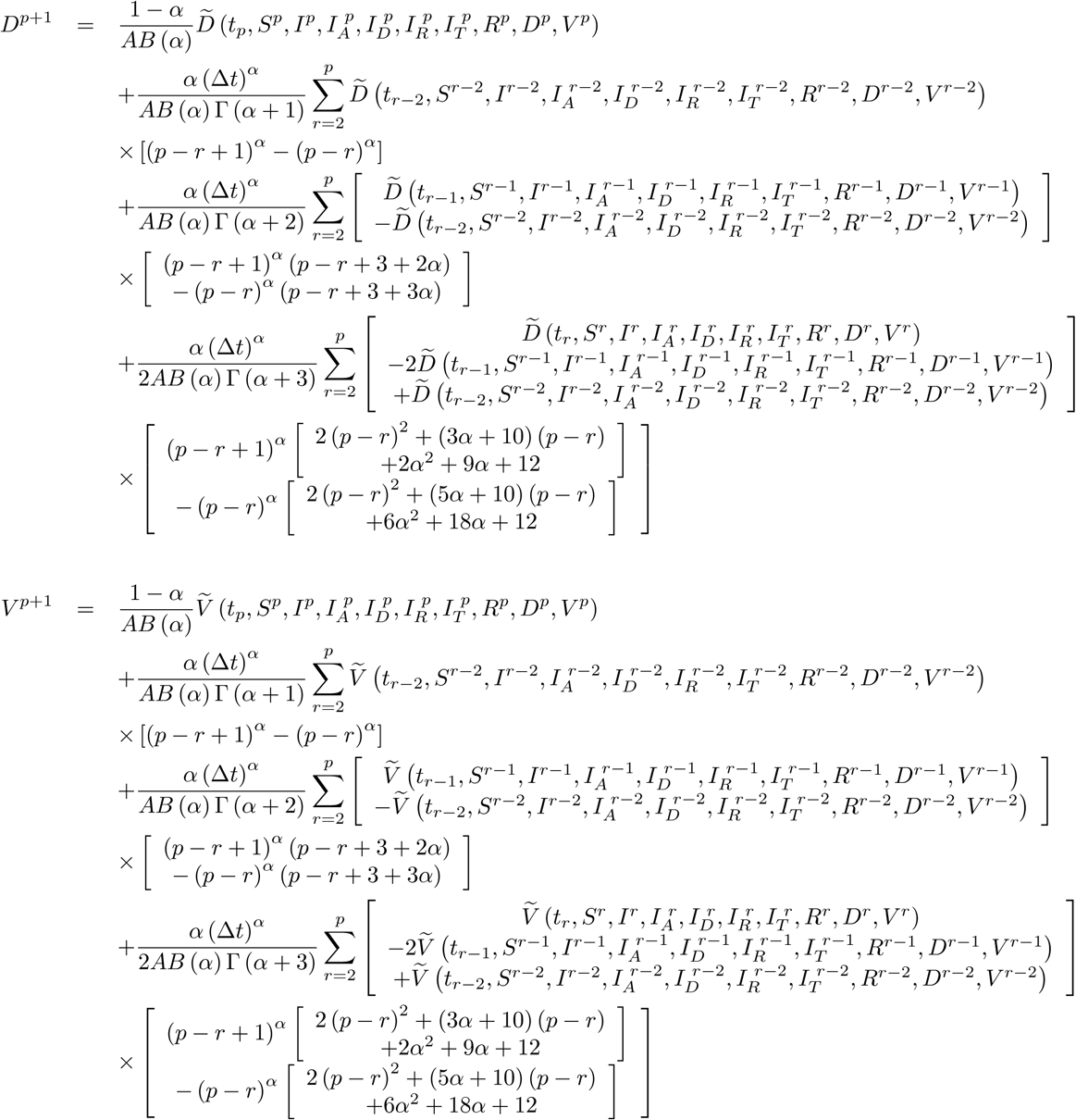

For power-law kernel, we can have the following

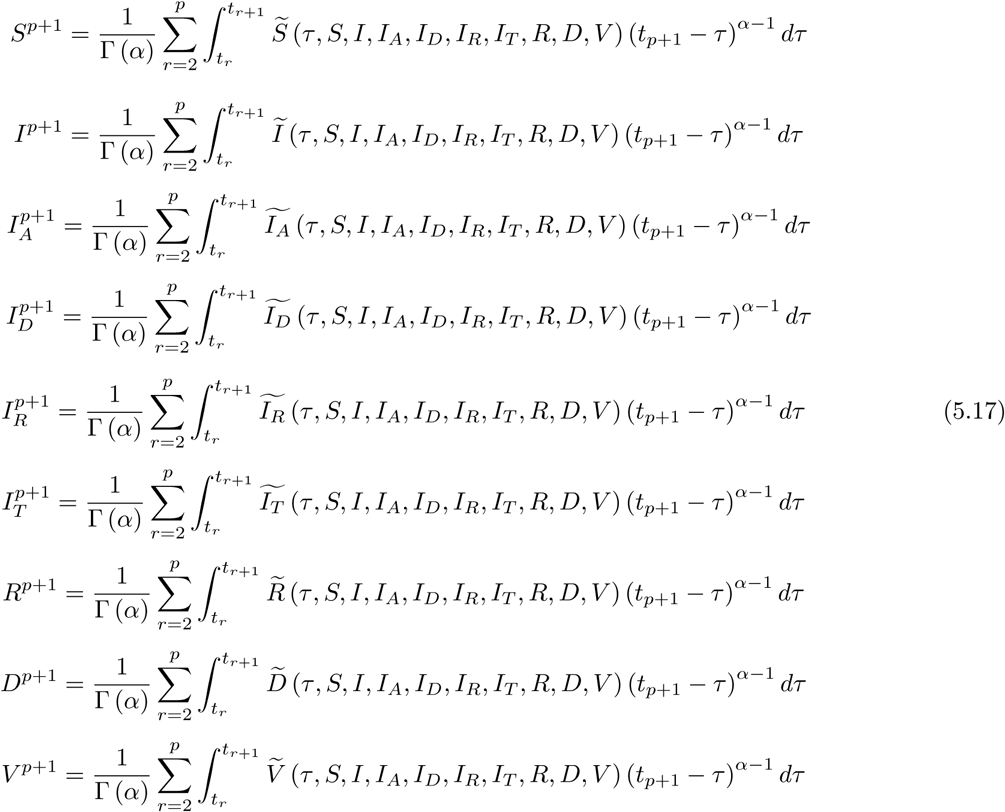

We can get the following numerical scheme

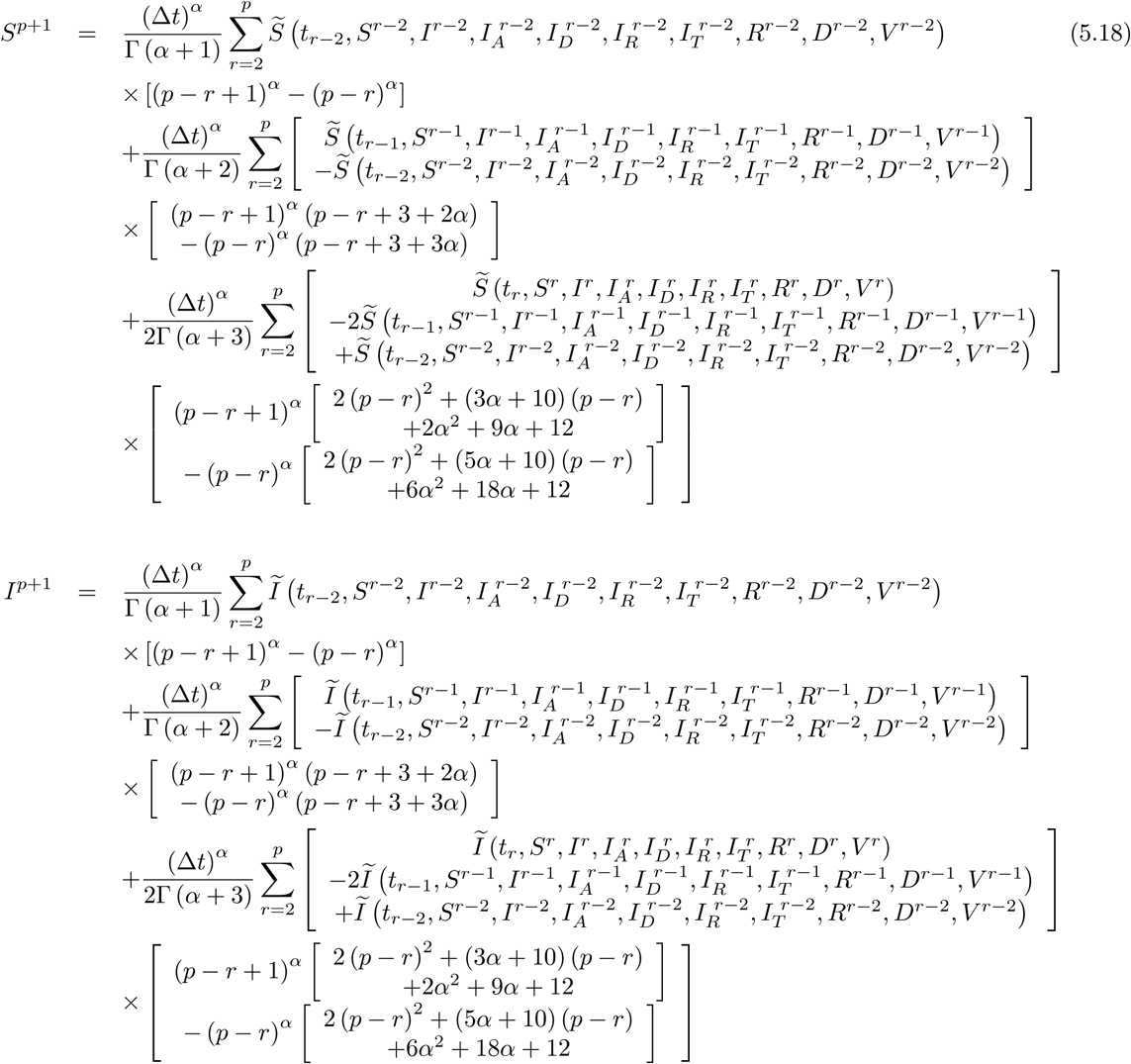

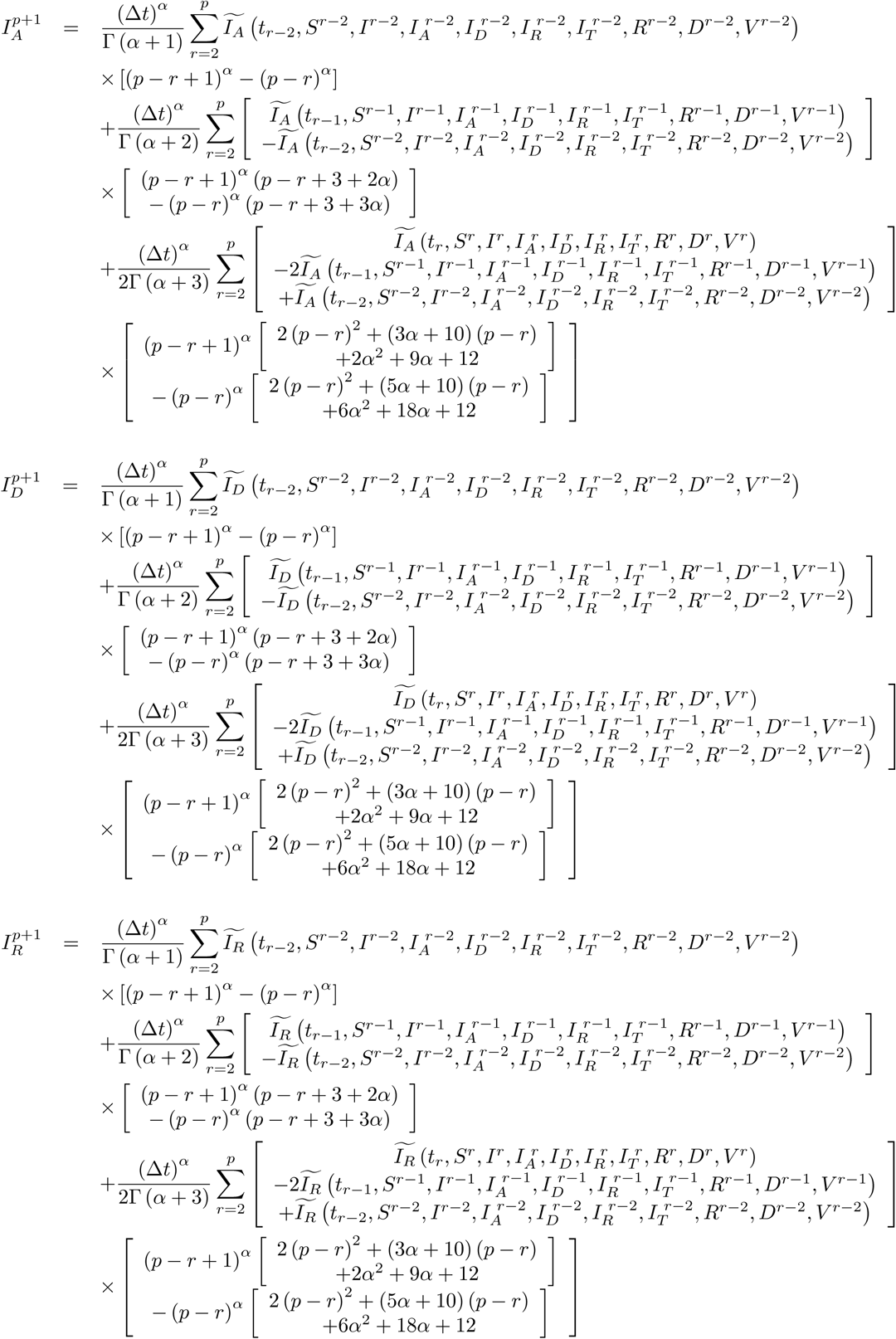

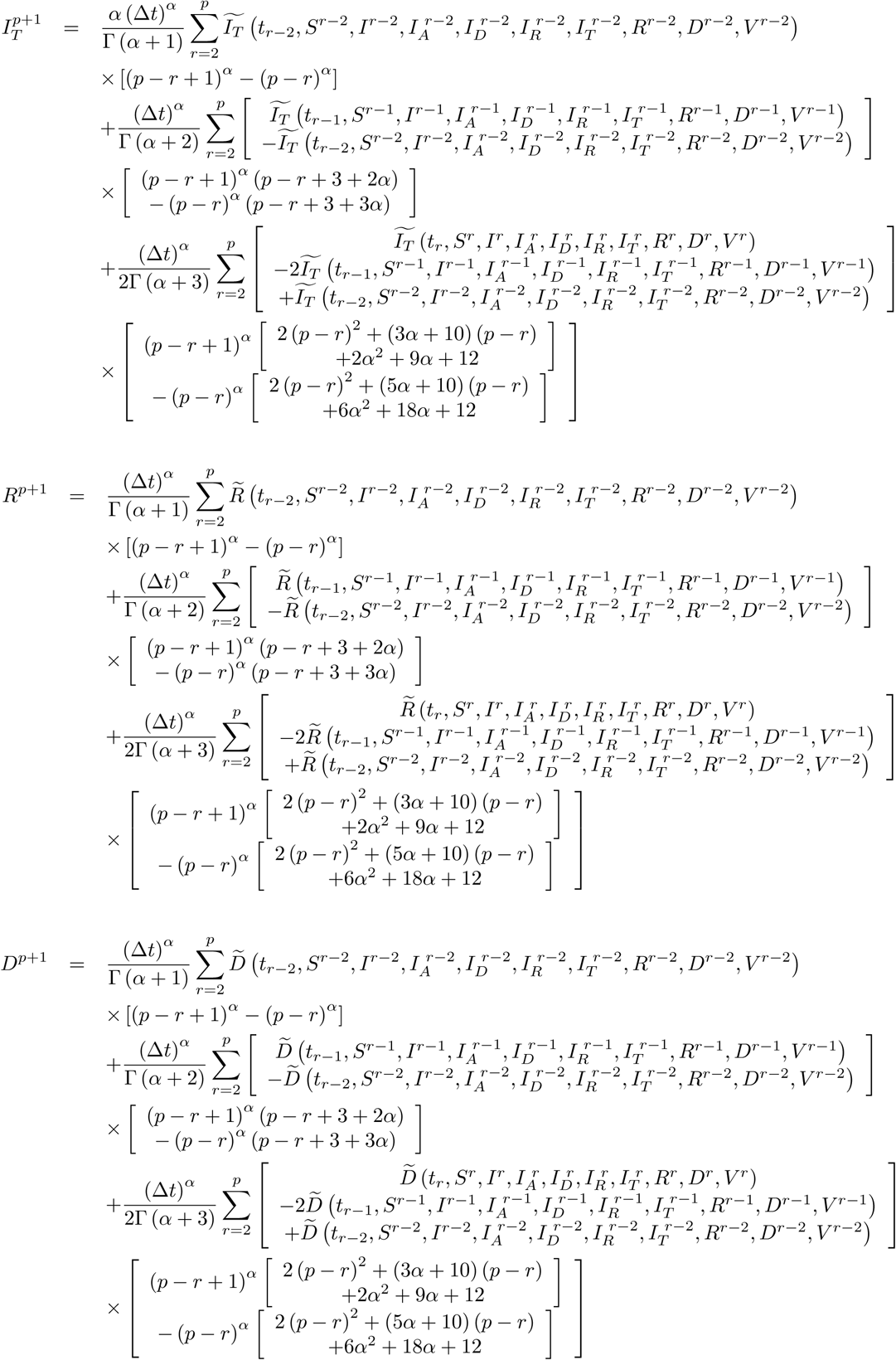

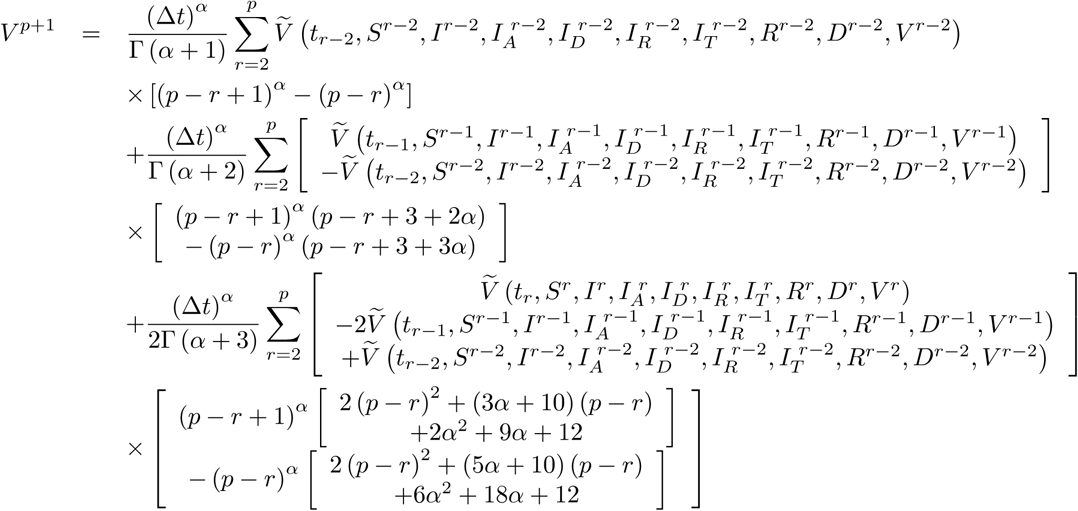

Now, we handle the following model

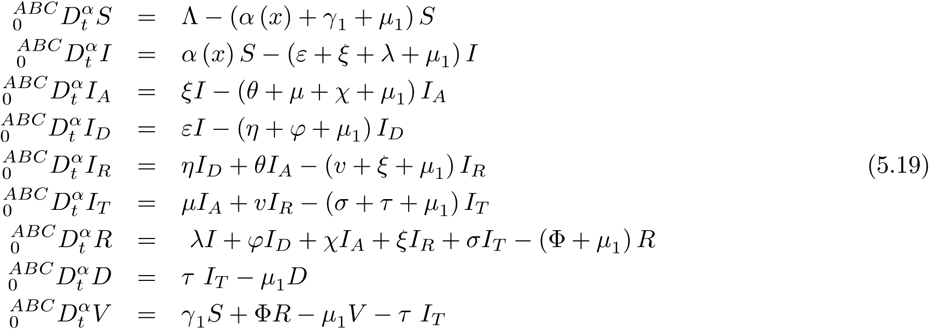

where initial conditions are

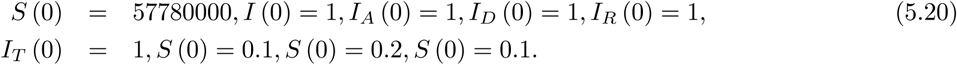

Also the parameters are chosen as

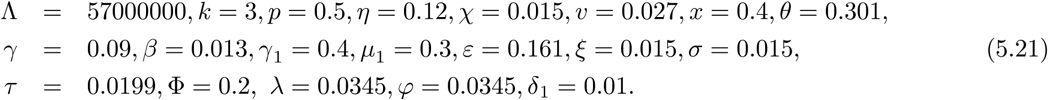

We present numerical simulation for COVID-19 model in Figure 37 and 38.4

**Figure 37.**
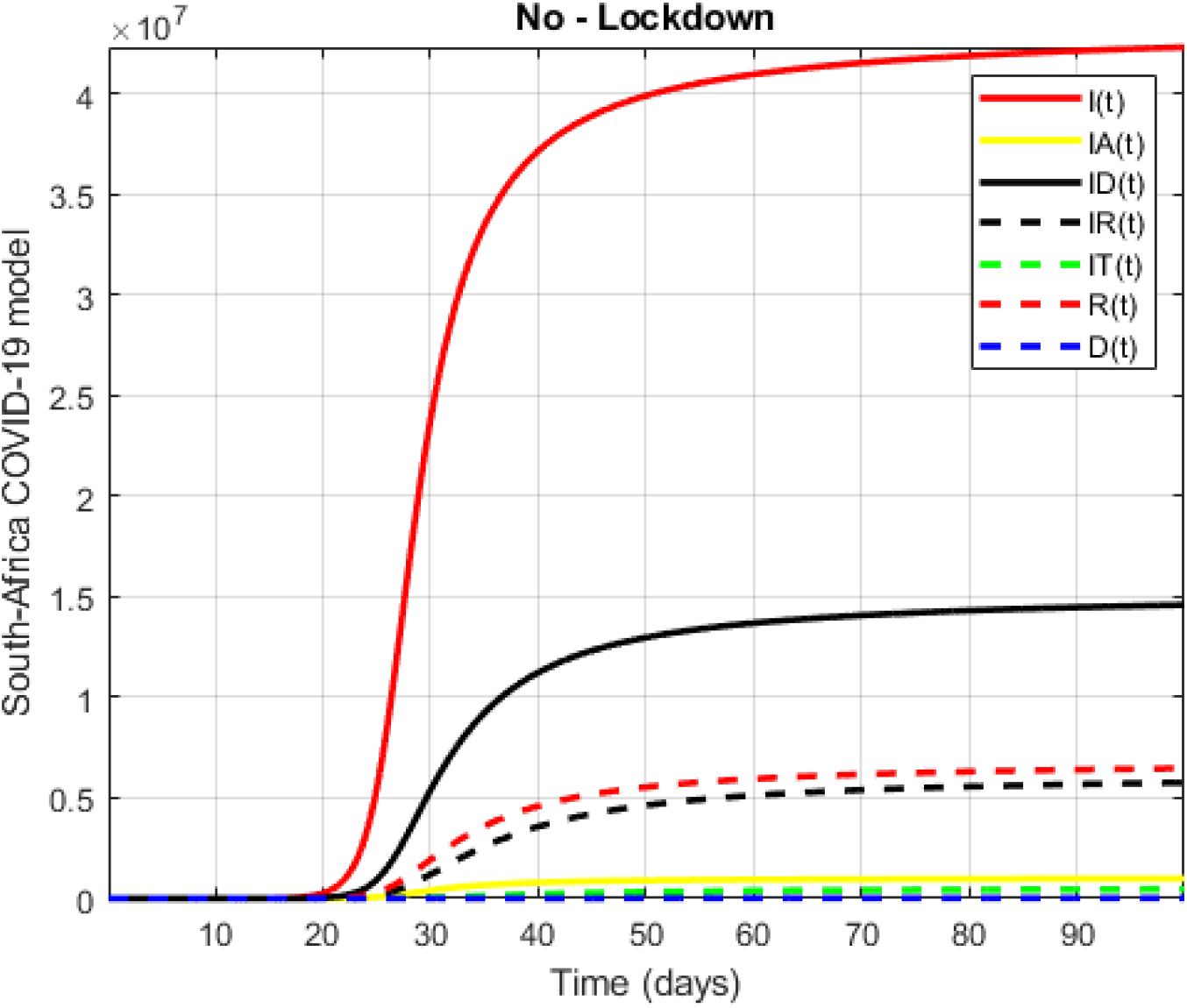
Numerical visualization for COVID-19 model in South Africa for *α* = 0.75.

**Figure 38.**
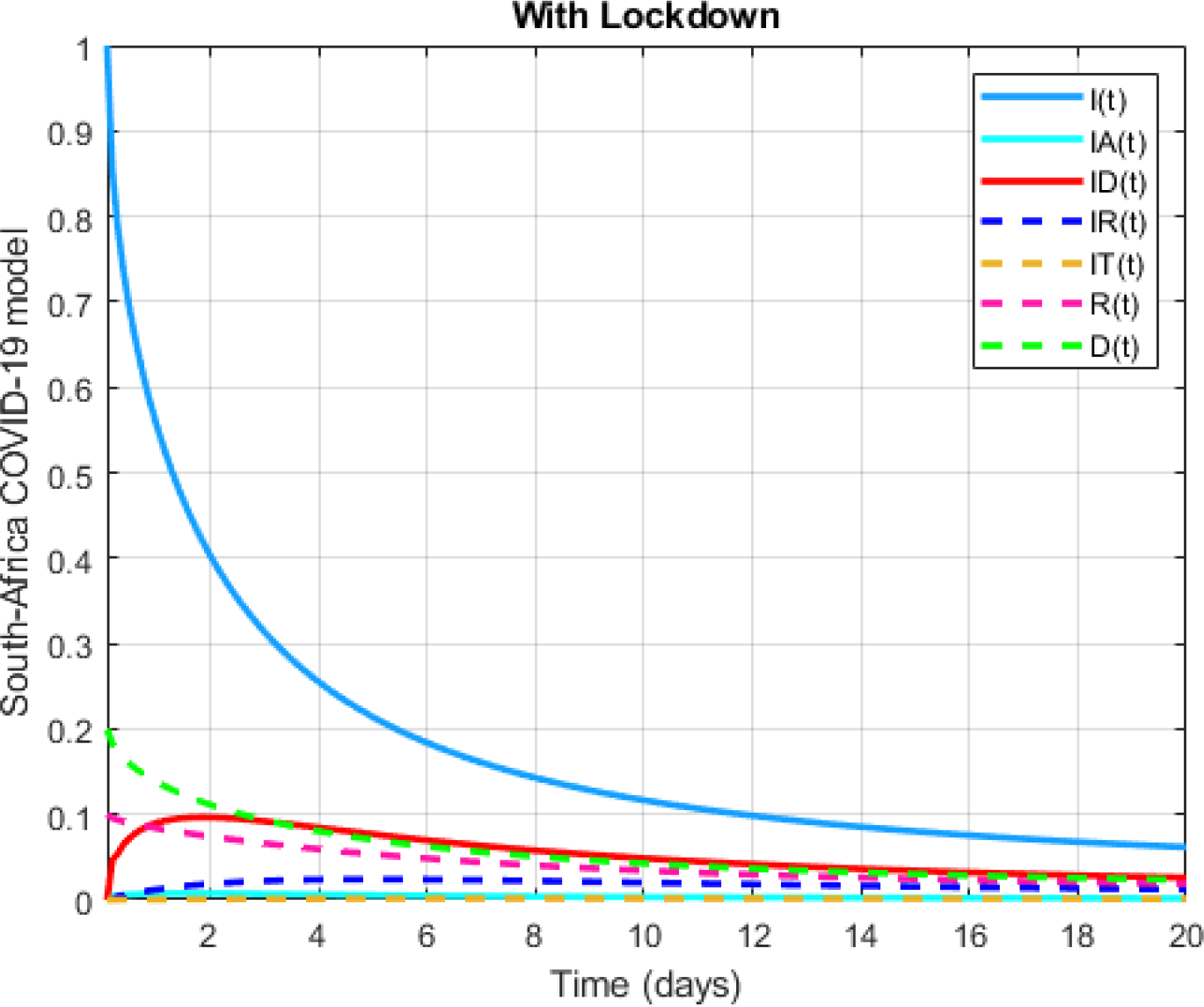
Numerical visualization for COVID-19 model in South Africa for *α* = 0:75:

For same model, initial conditions are chosen

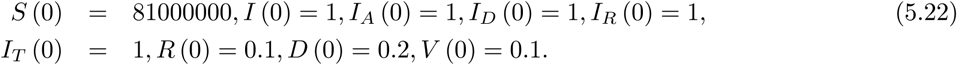

Also the parameters are

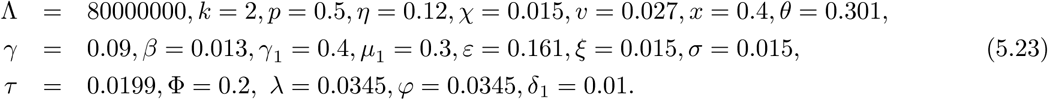

We present numerical simulation for COVID-19 model in Figure 39 and 40.

**Figure 39.**
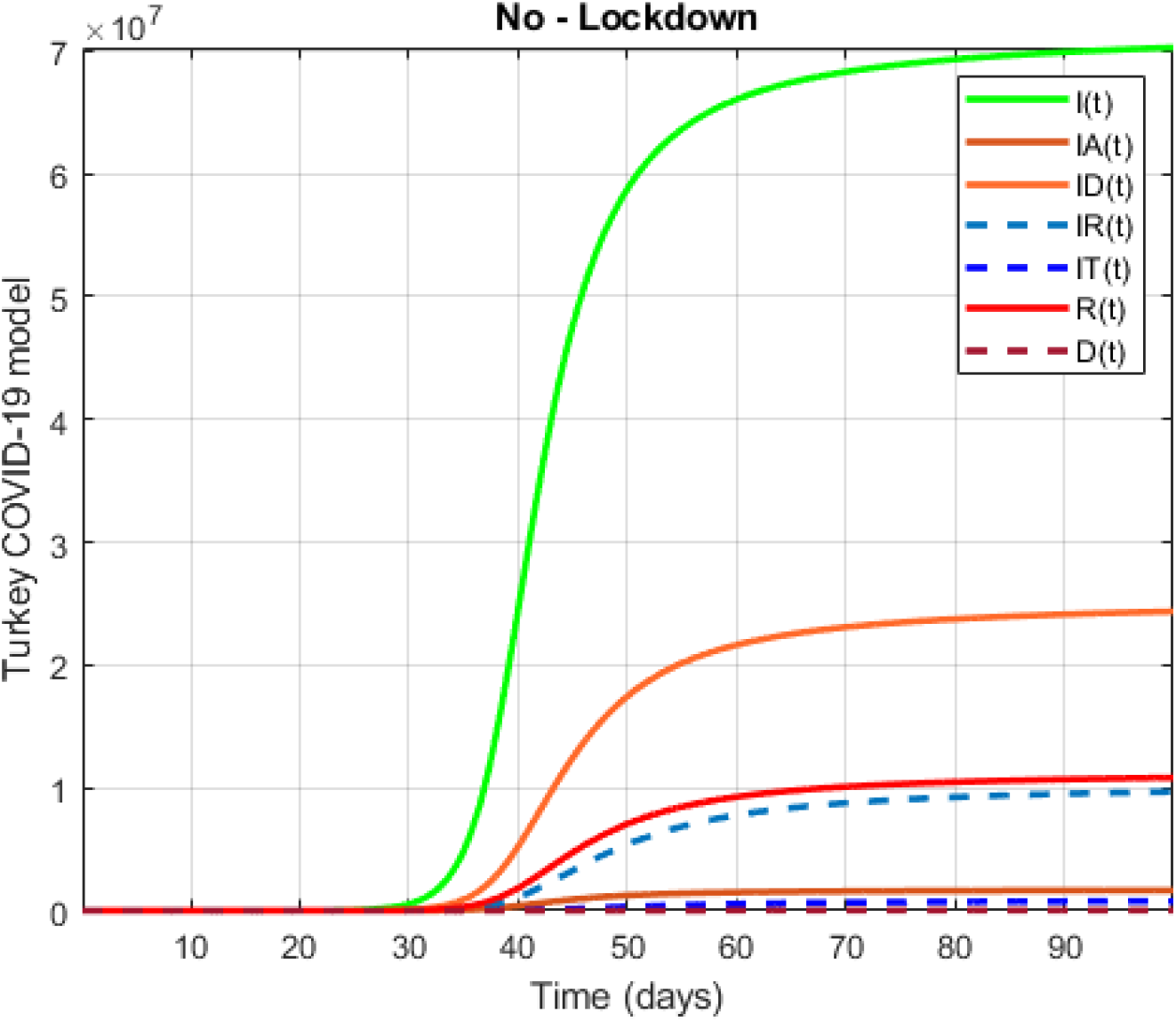
Numerical visualization for COVID-19 model in Turkey for *α* = 0.8.

**Figure 40.**
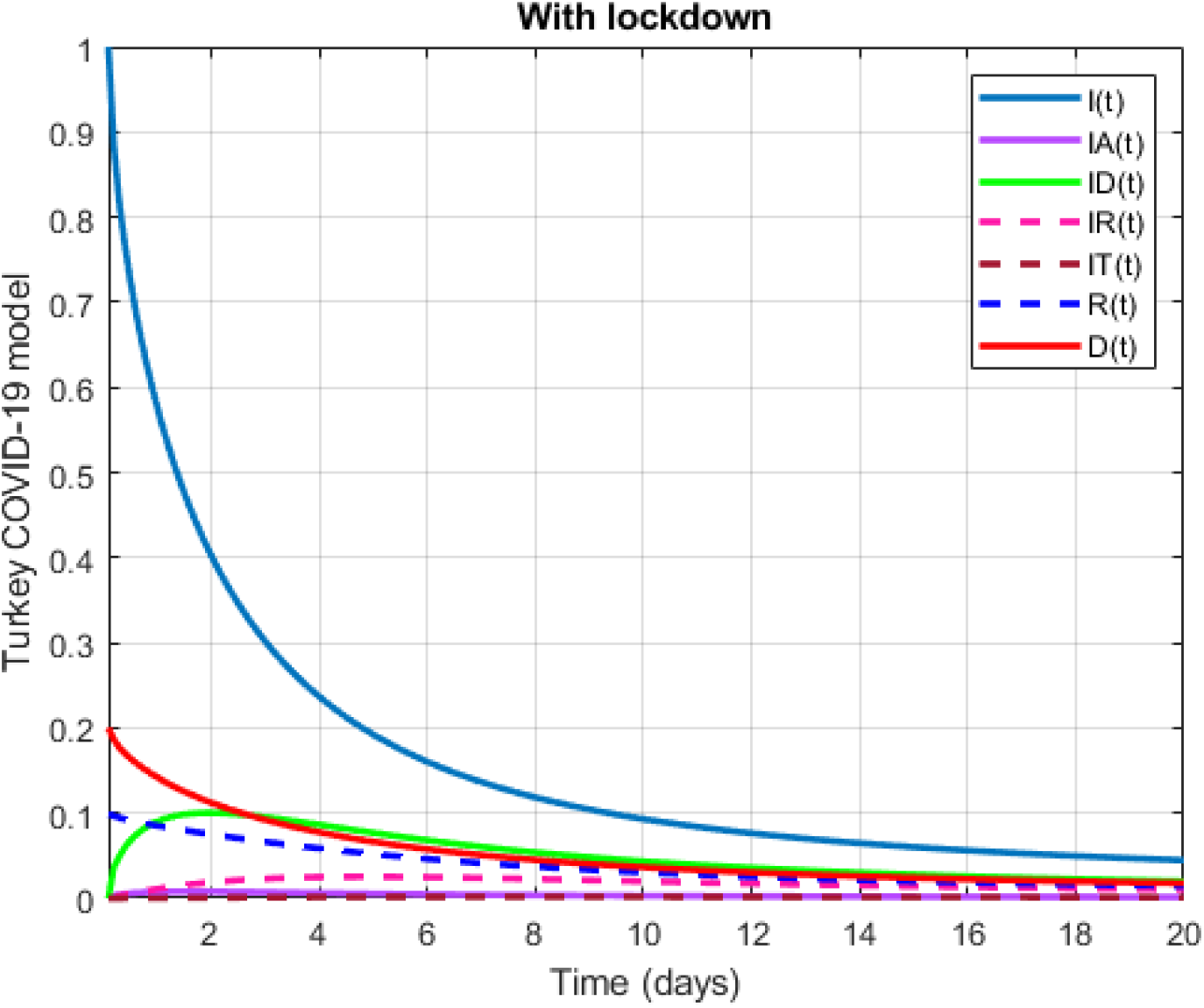
Numerical visualization for COVID-19 model in Turkey for *α* = 0.8.

Now, we replace the classical differential operator will be replaced by the operator with power-law, exponential decay and Mittag-Leffler kernels. We start with exponential decay kernel

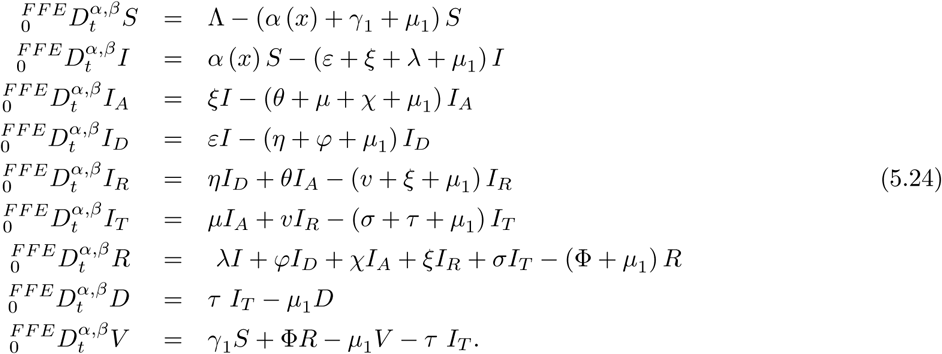

For simplicity, we write above equation as follows;

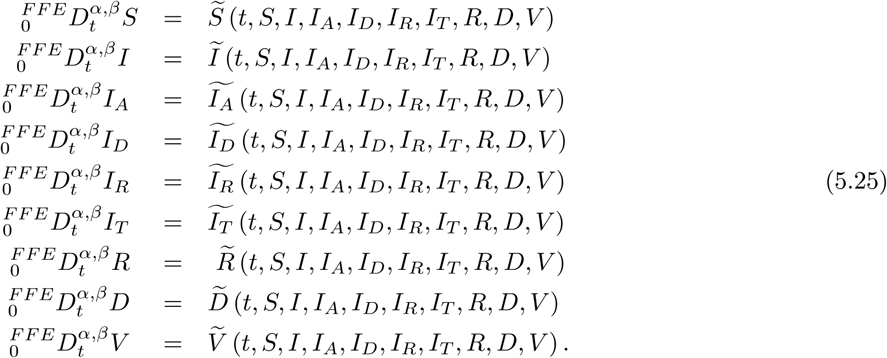

After applying fractal-fractional integral with exponential kernel, we have the following

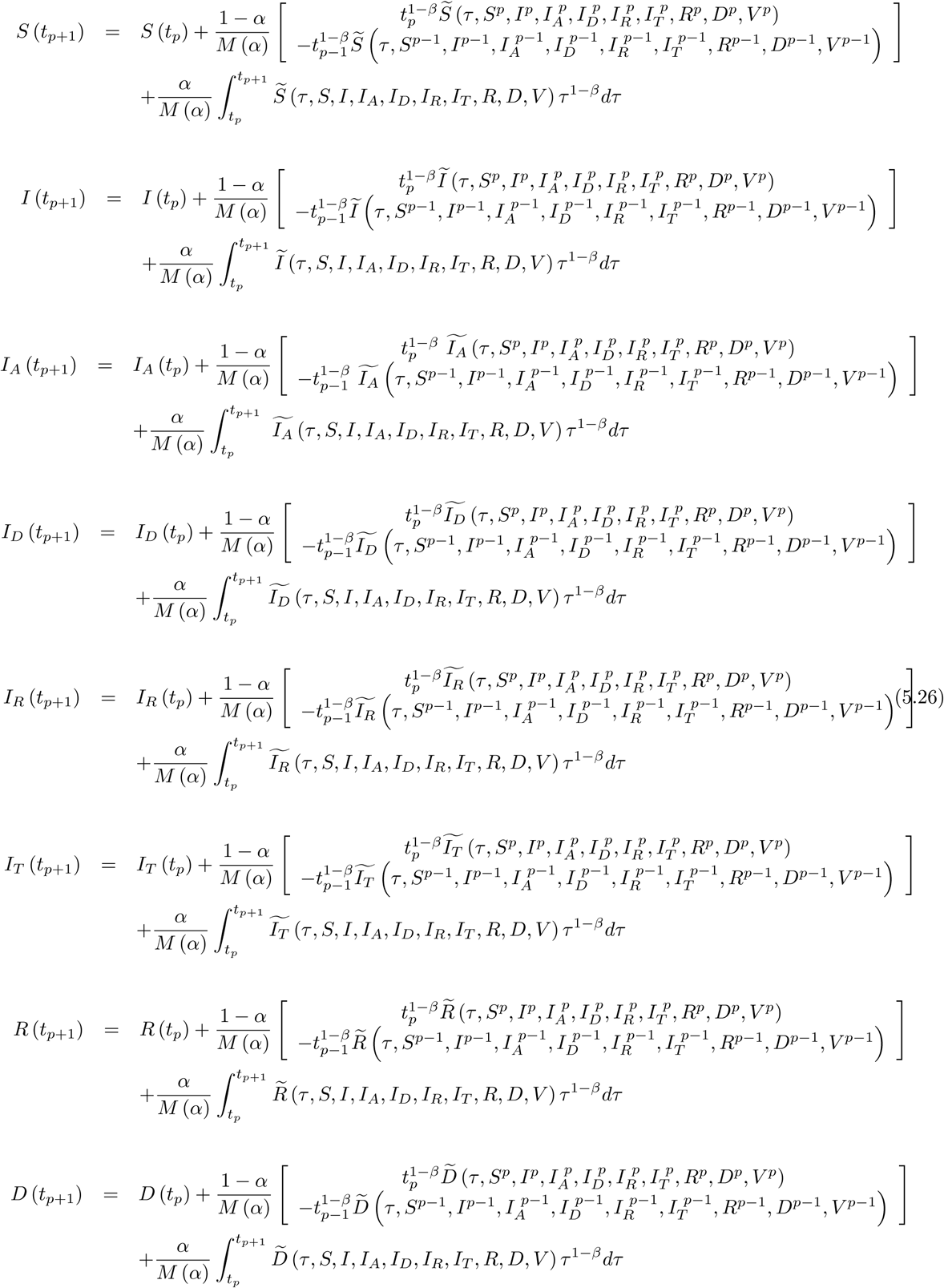

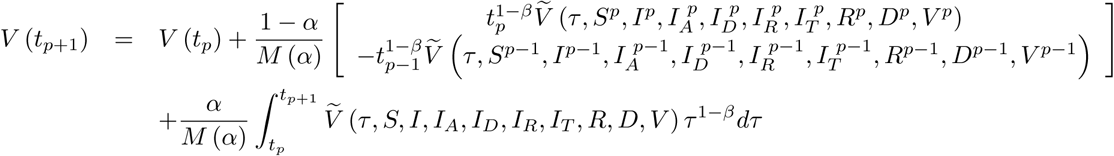

We can have the following scheme for this model

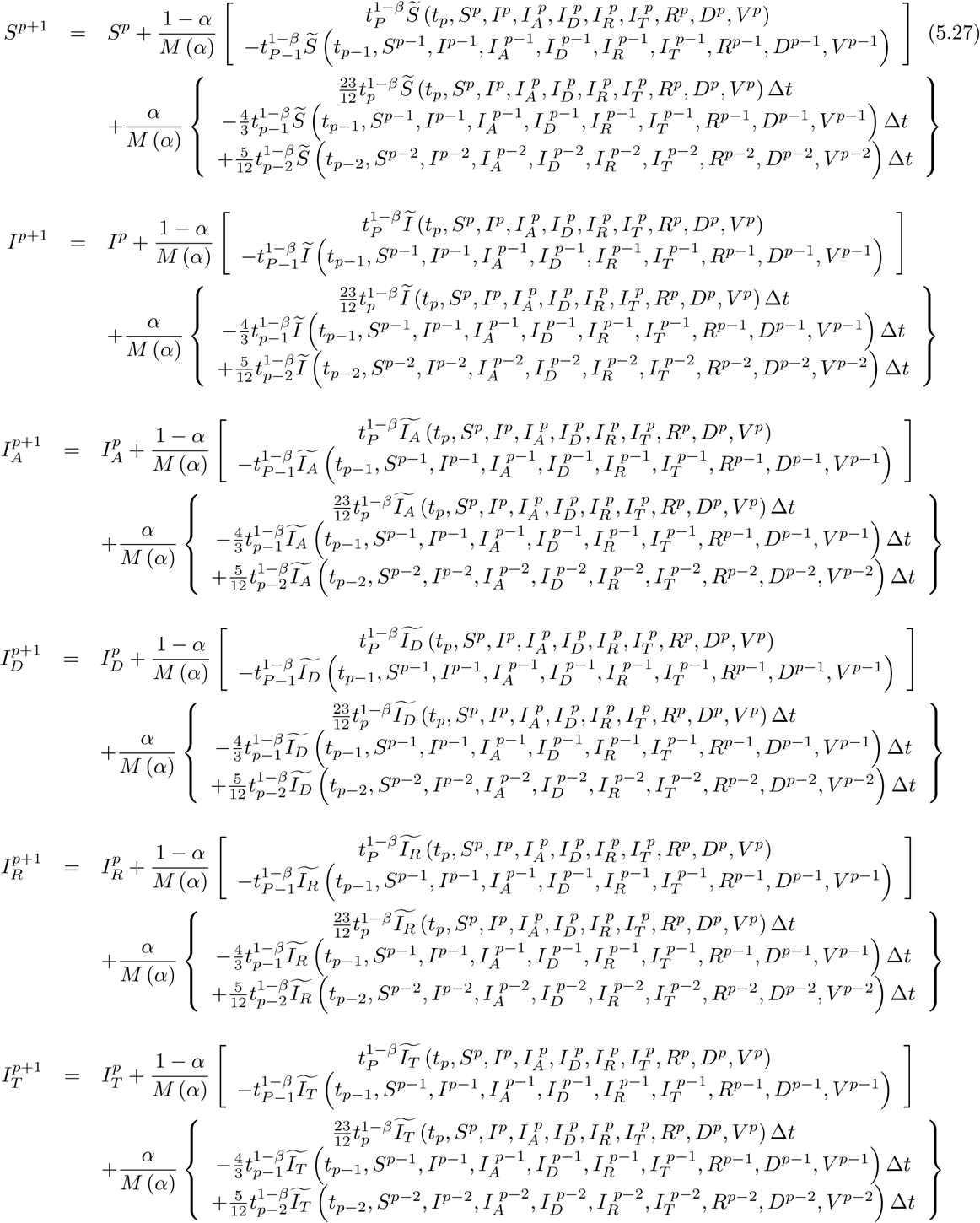

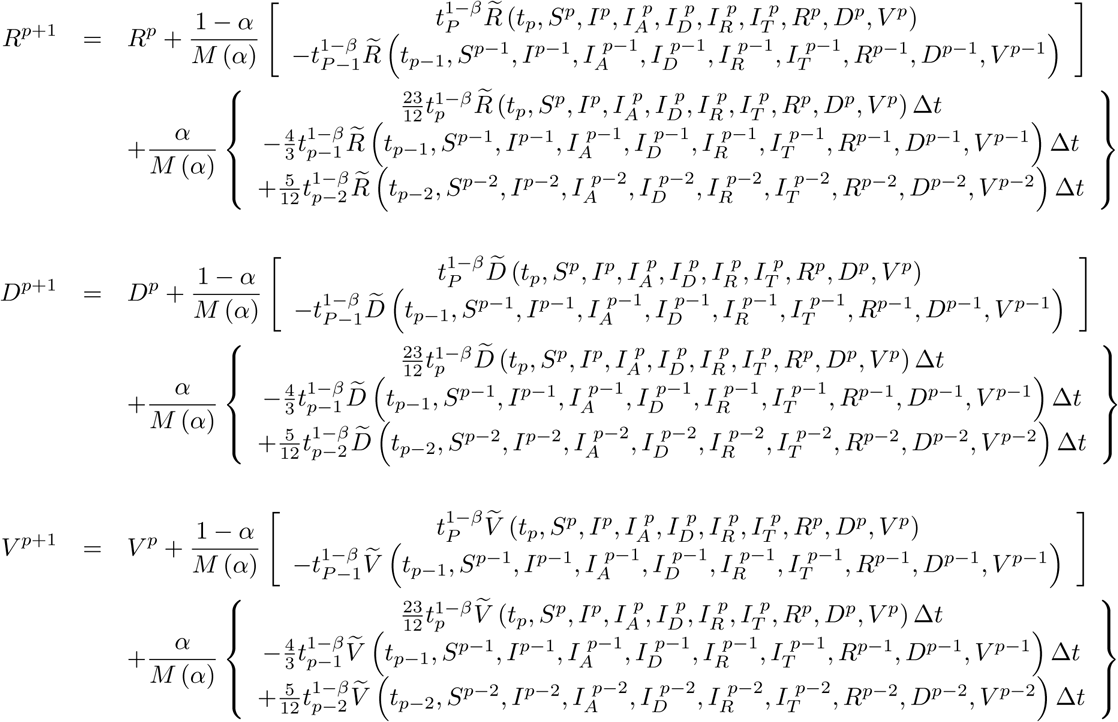

For Mittag-Leffler kernel, we can have the following

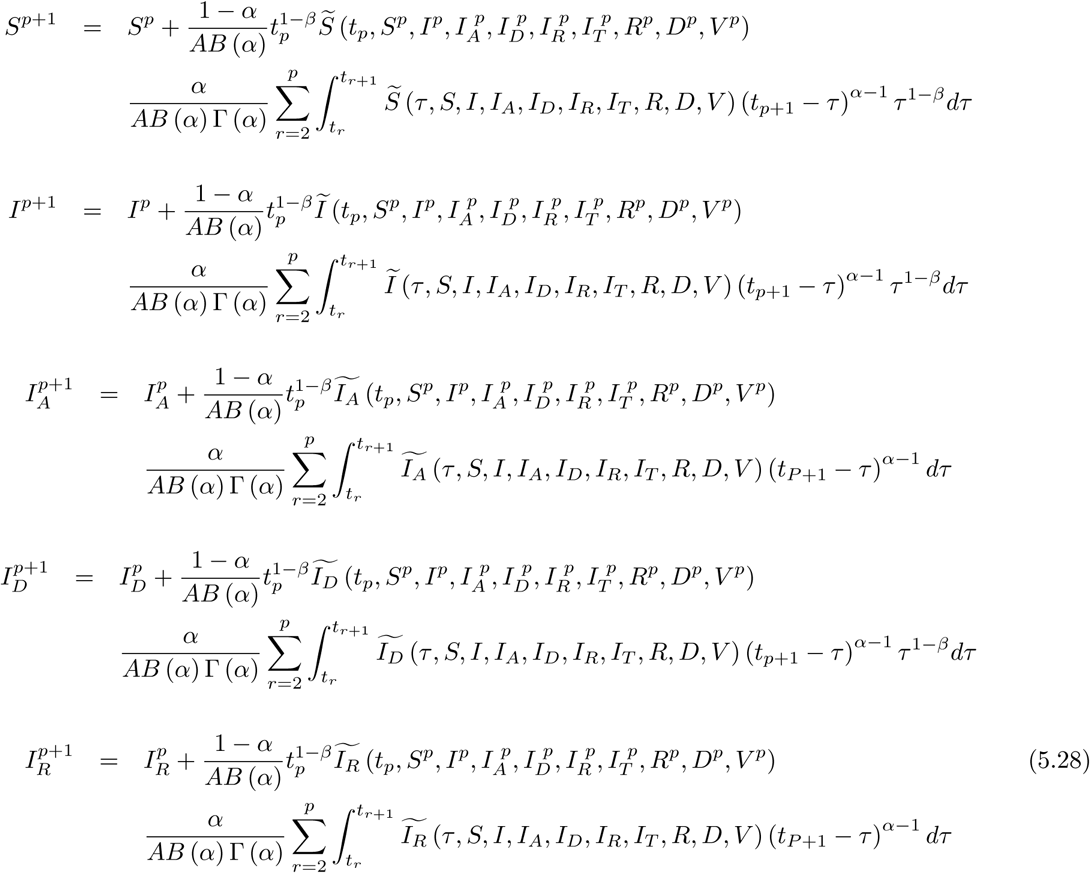

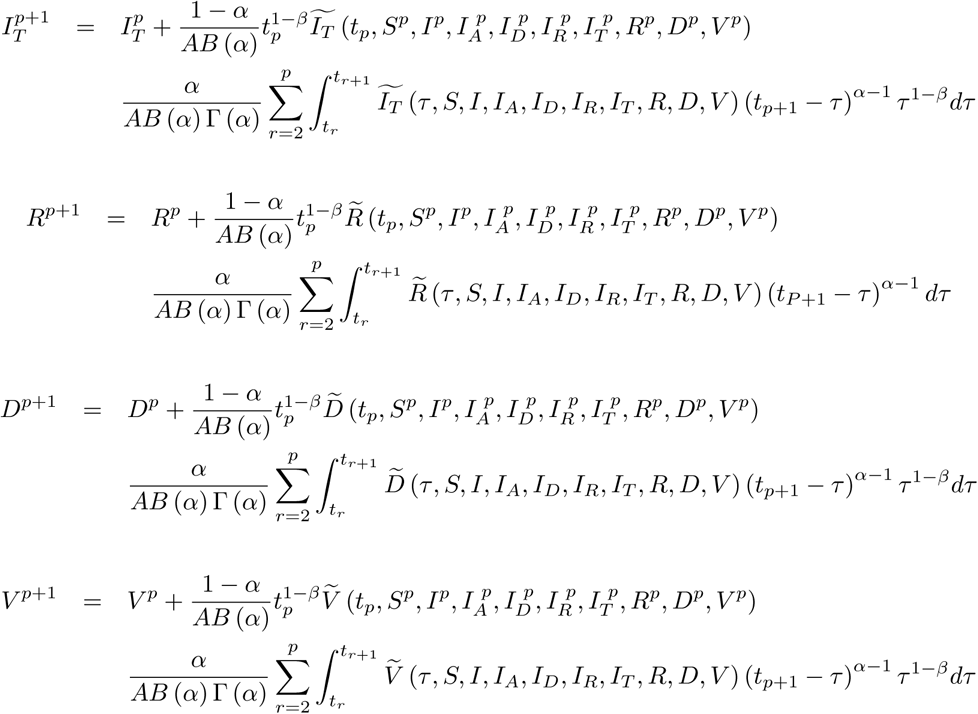

We can get the following numerical scheme

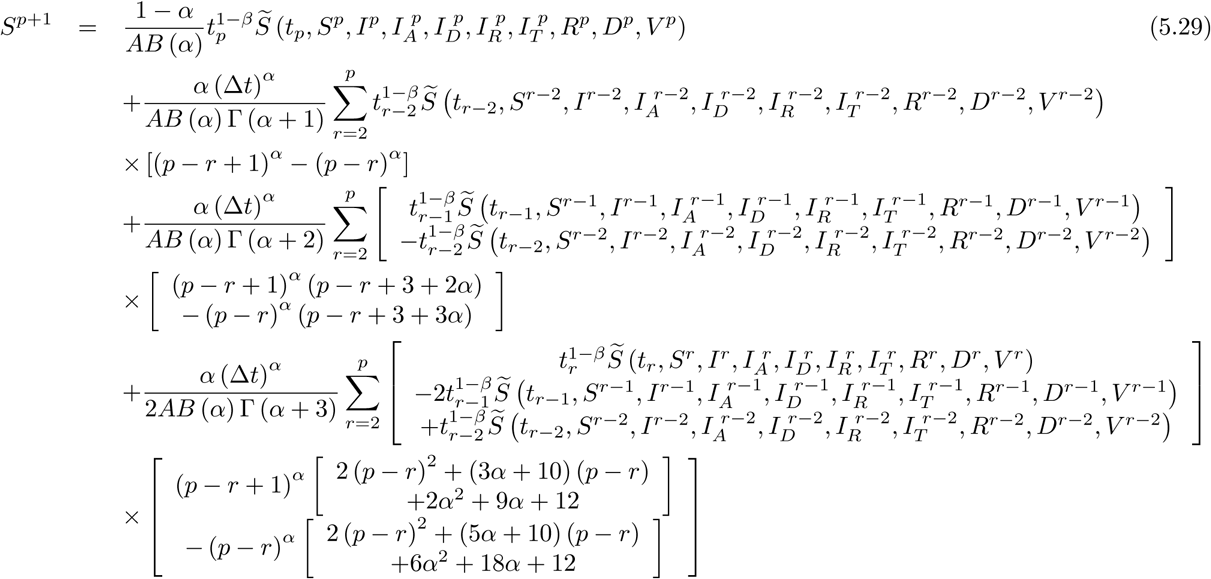

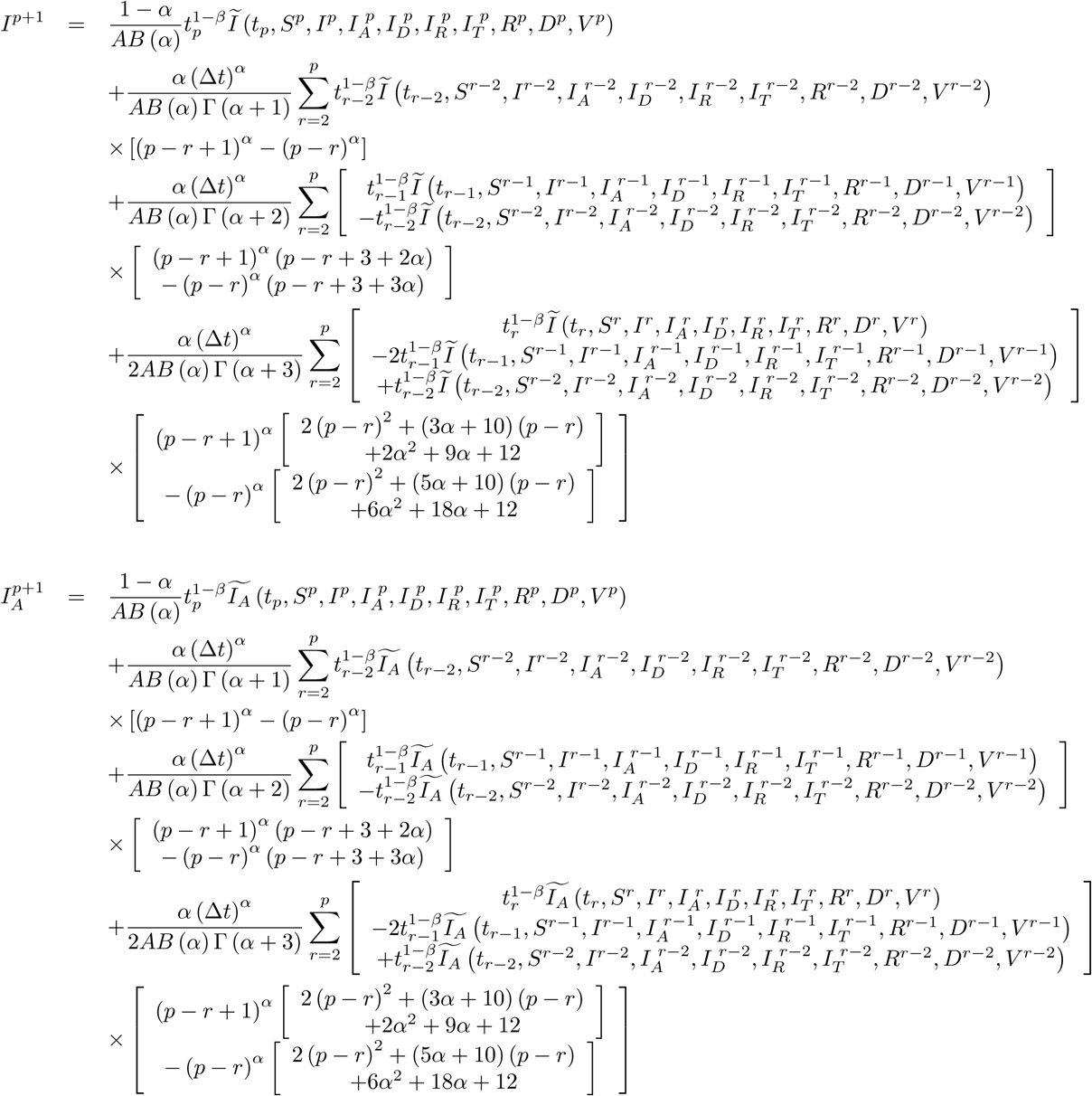

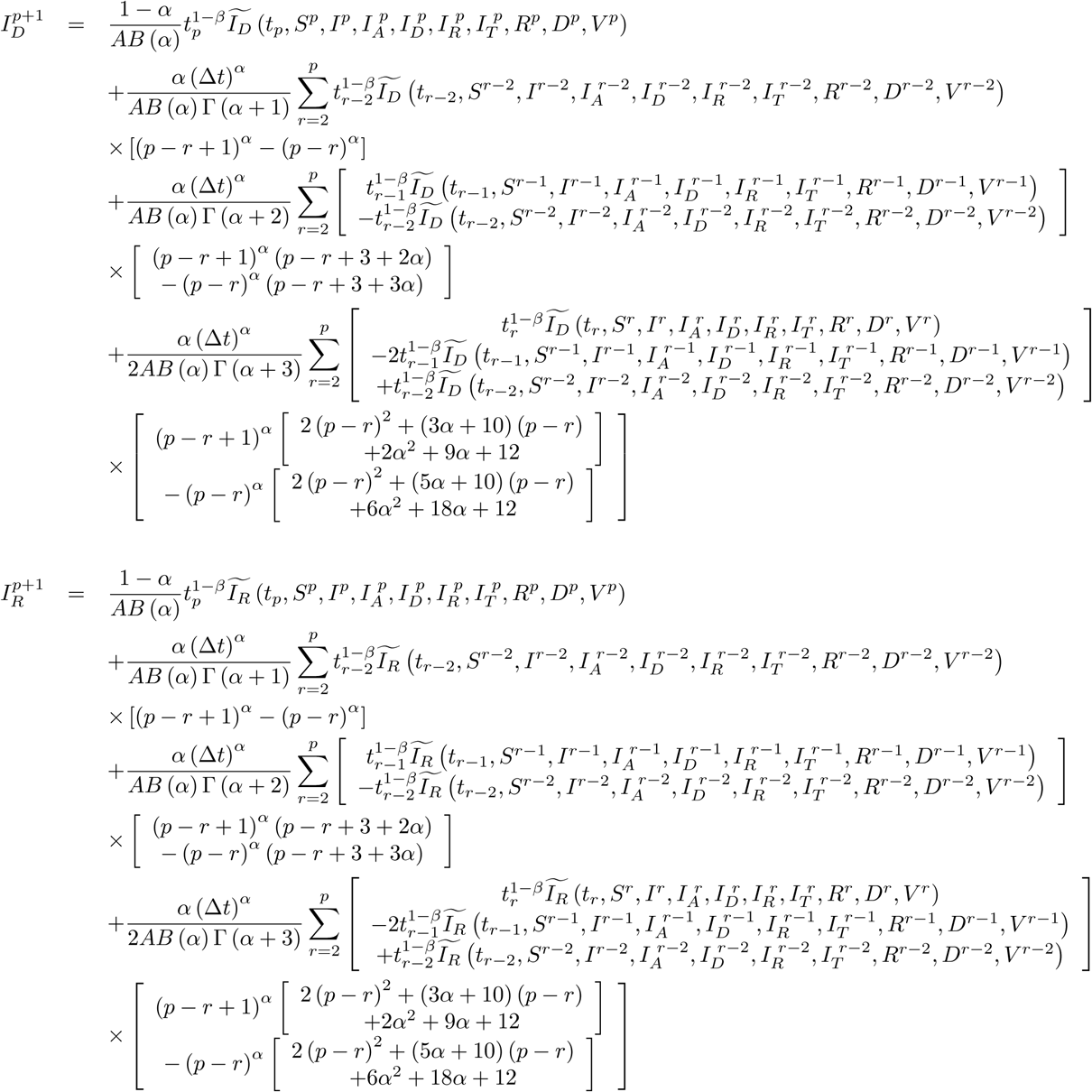

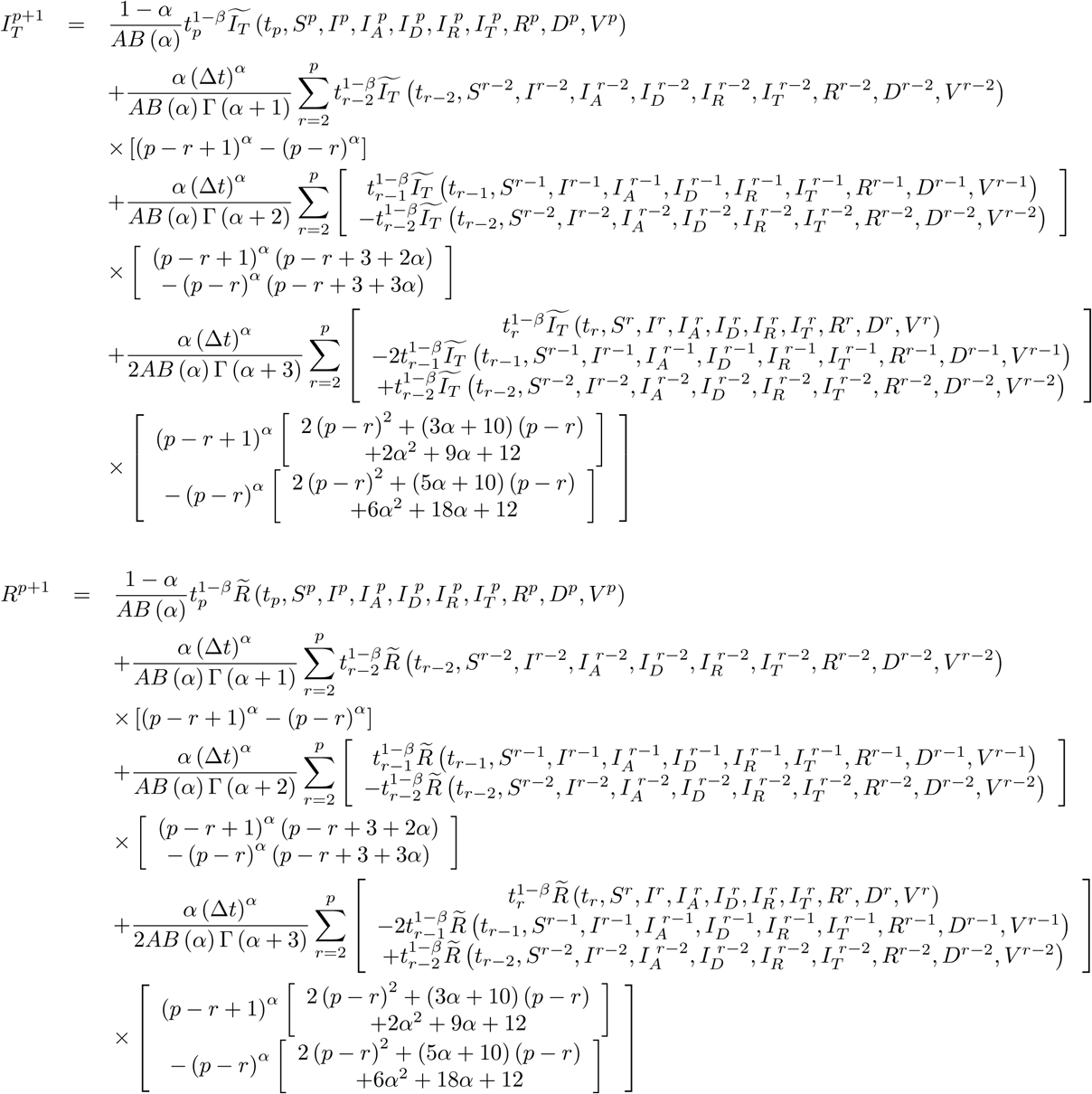

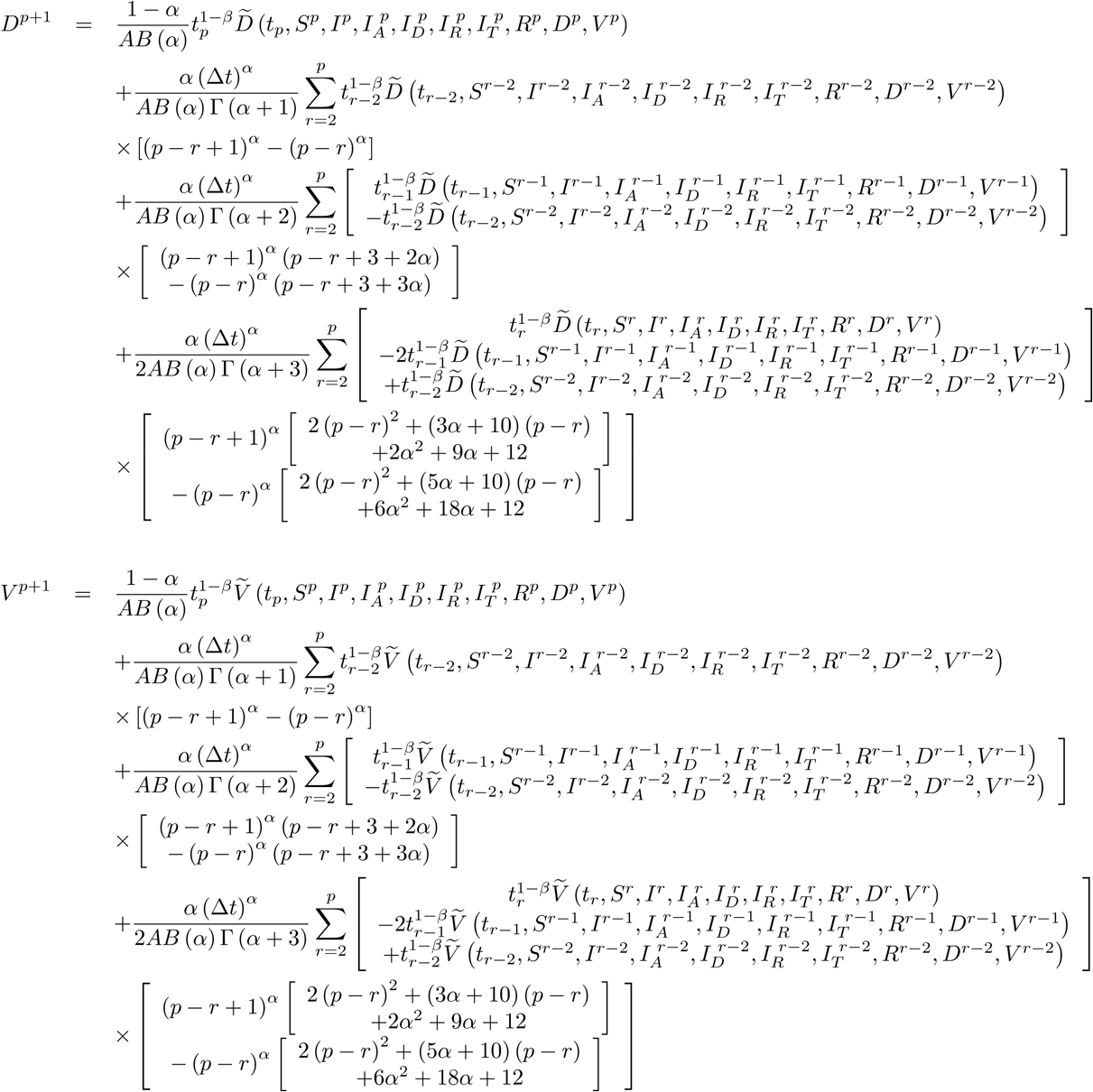

For power-law kernel, we can have the following

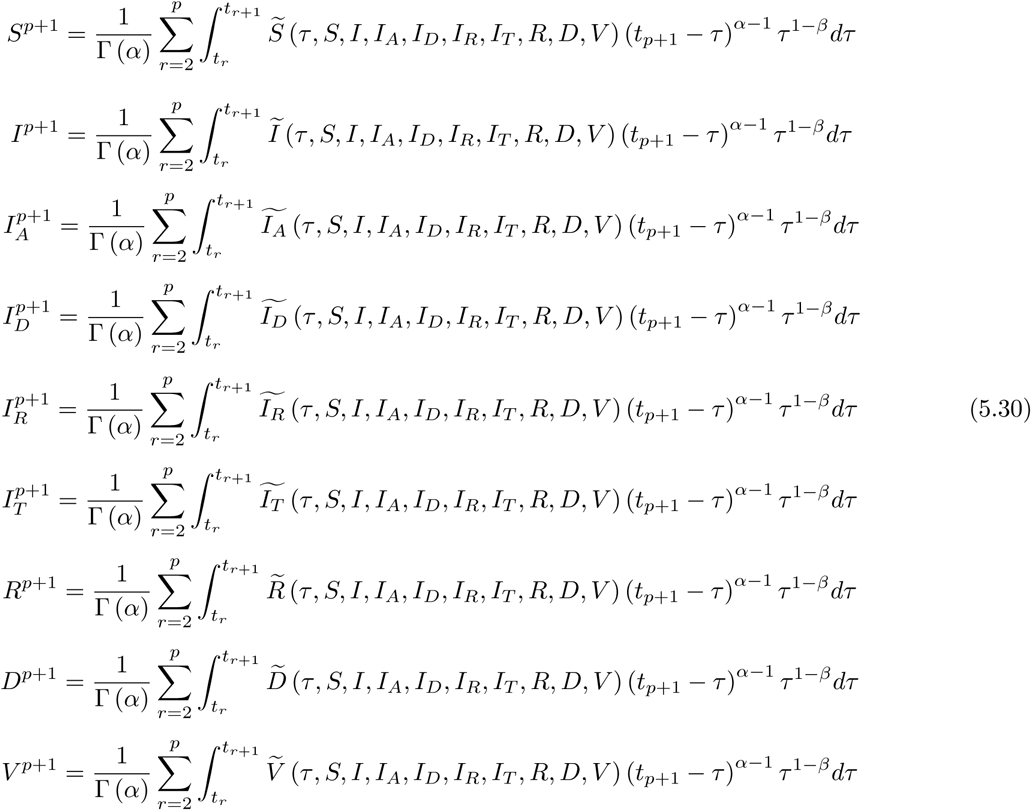

We can get the following numerical scheme

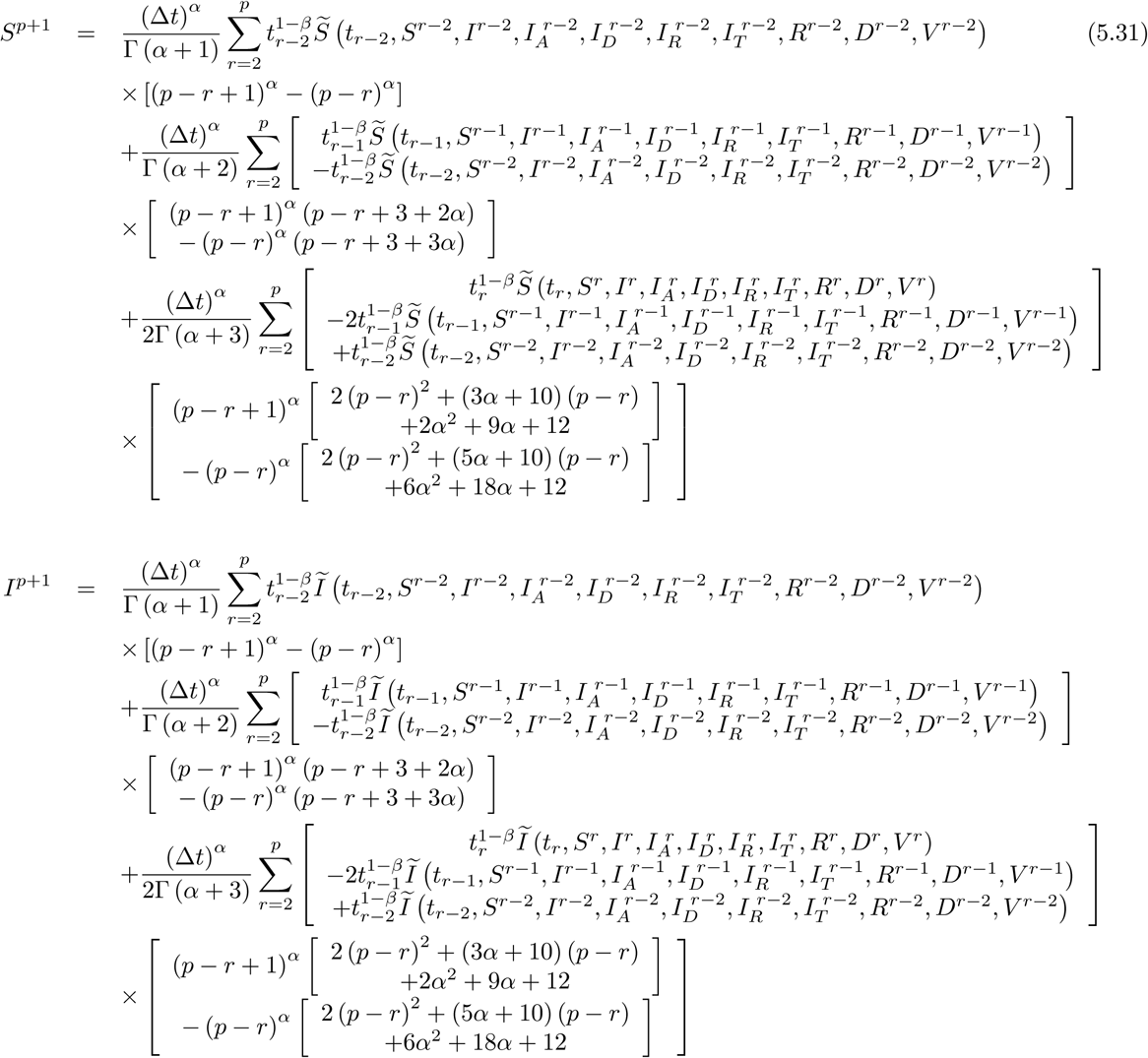

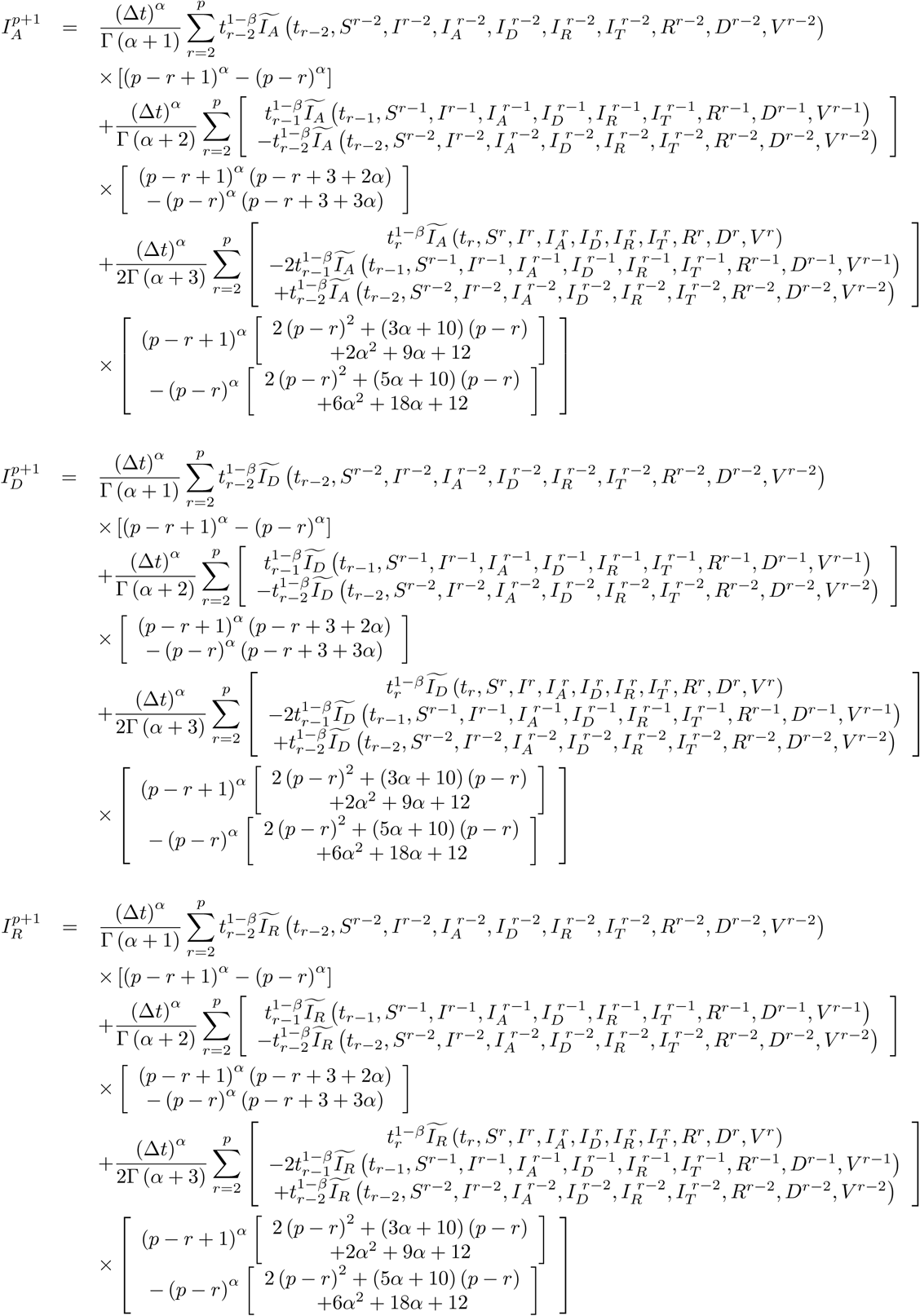

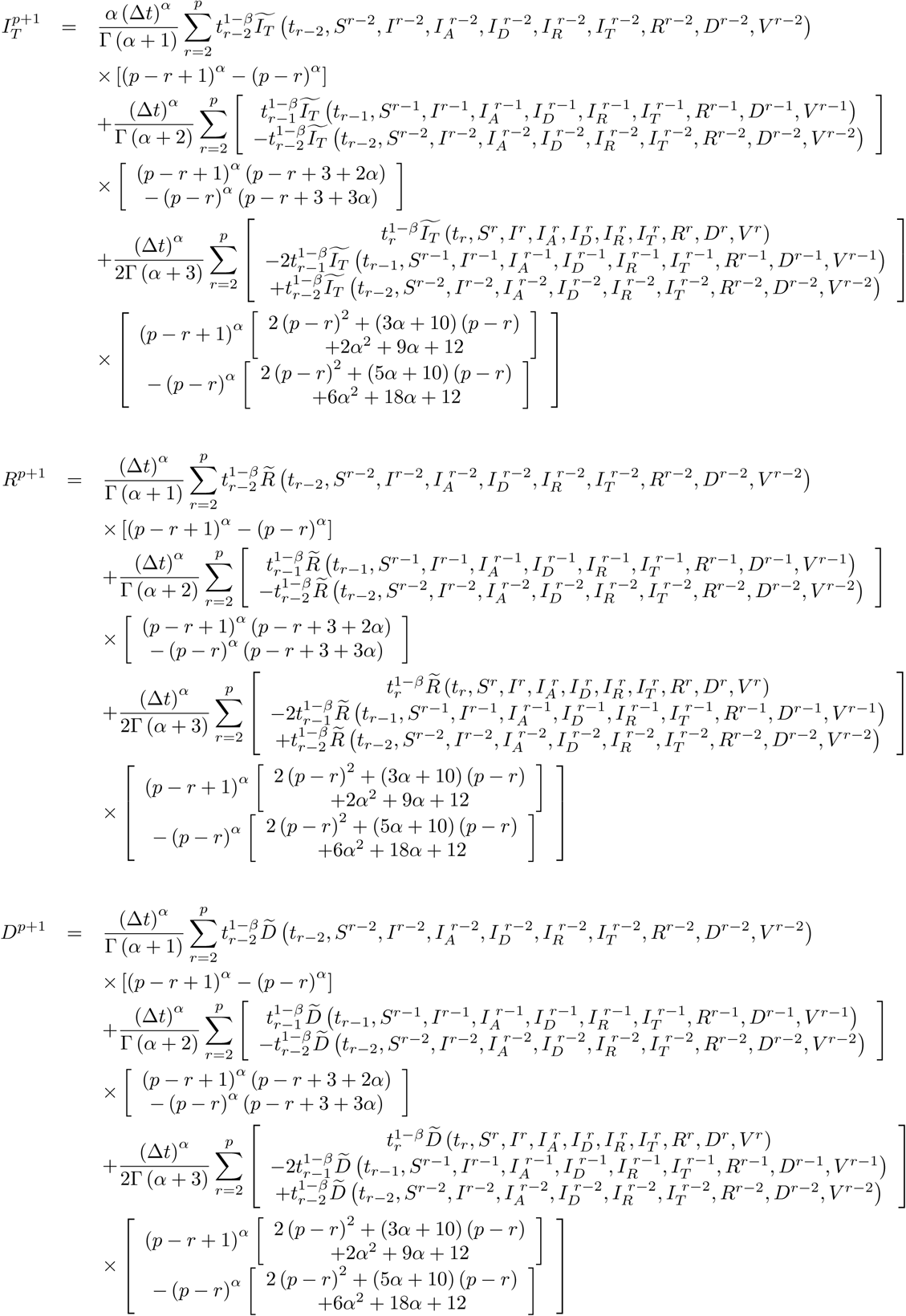

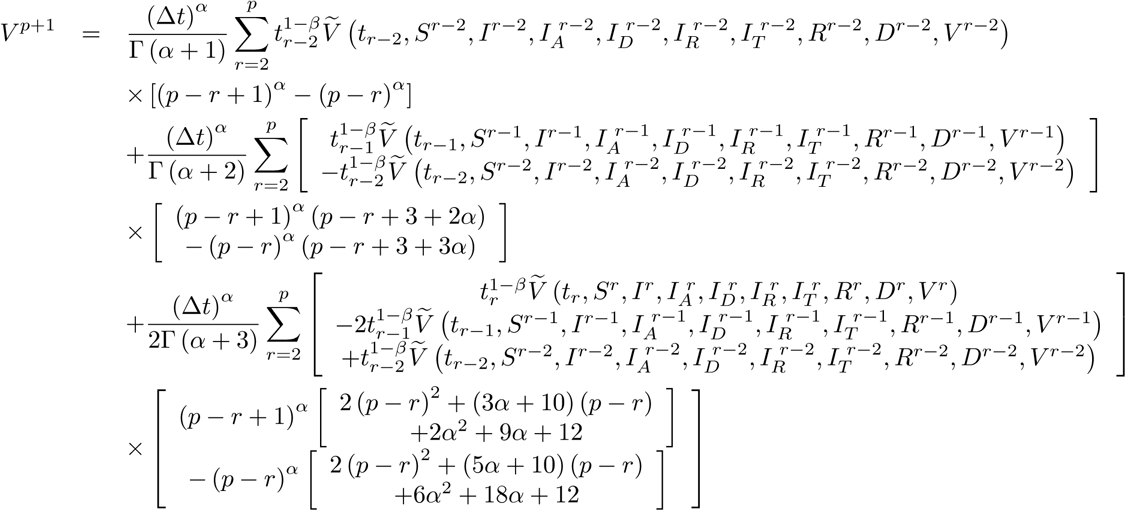

Now, we handle the following model

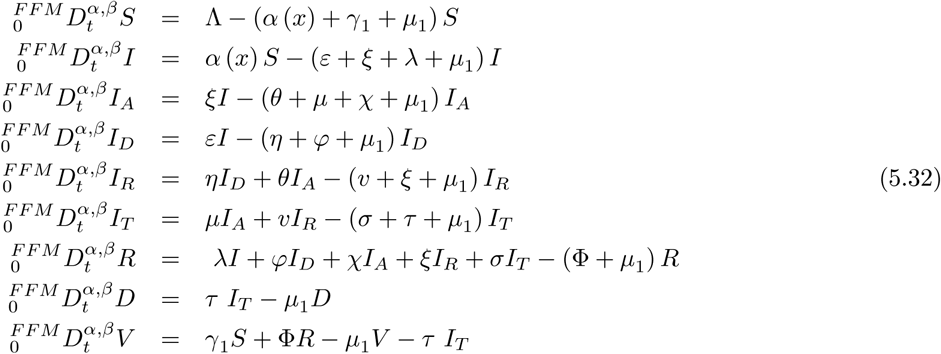

where initial conditions are

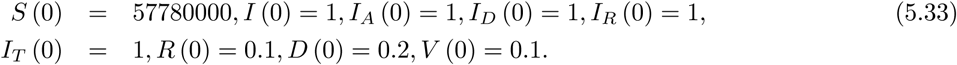

Also the parameters are chosen as

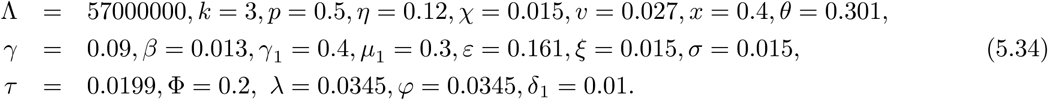

We present numerical simulation for COVID-19 model in Figure 41 and 42.

**Figure 41.**
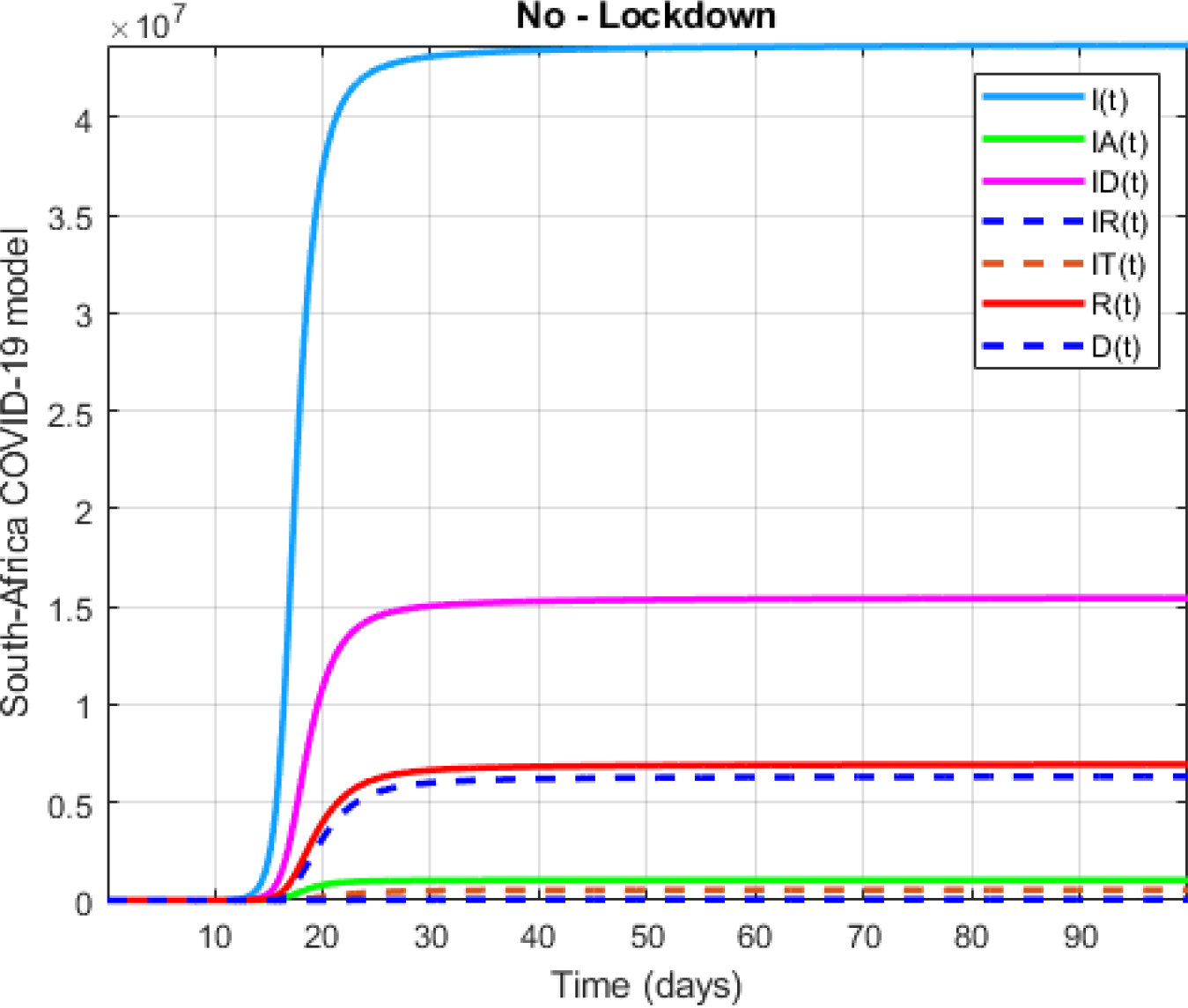
Numerical visualization for COVID-19 model in South Africa for *α* = 0.9, *β* = 0.75.

**Figure 42.**
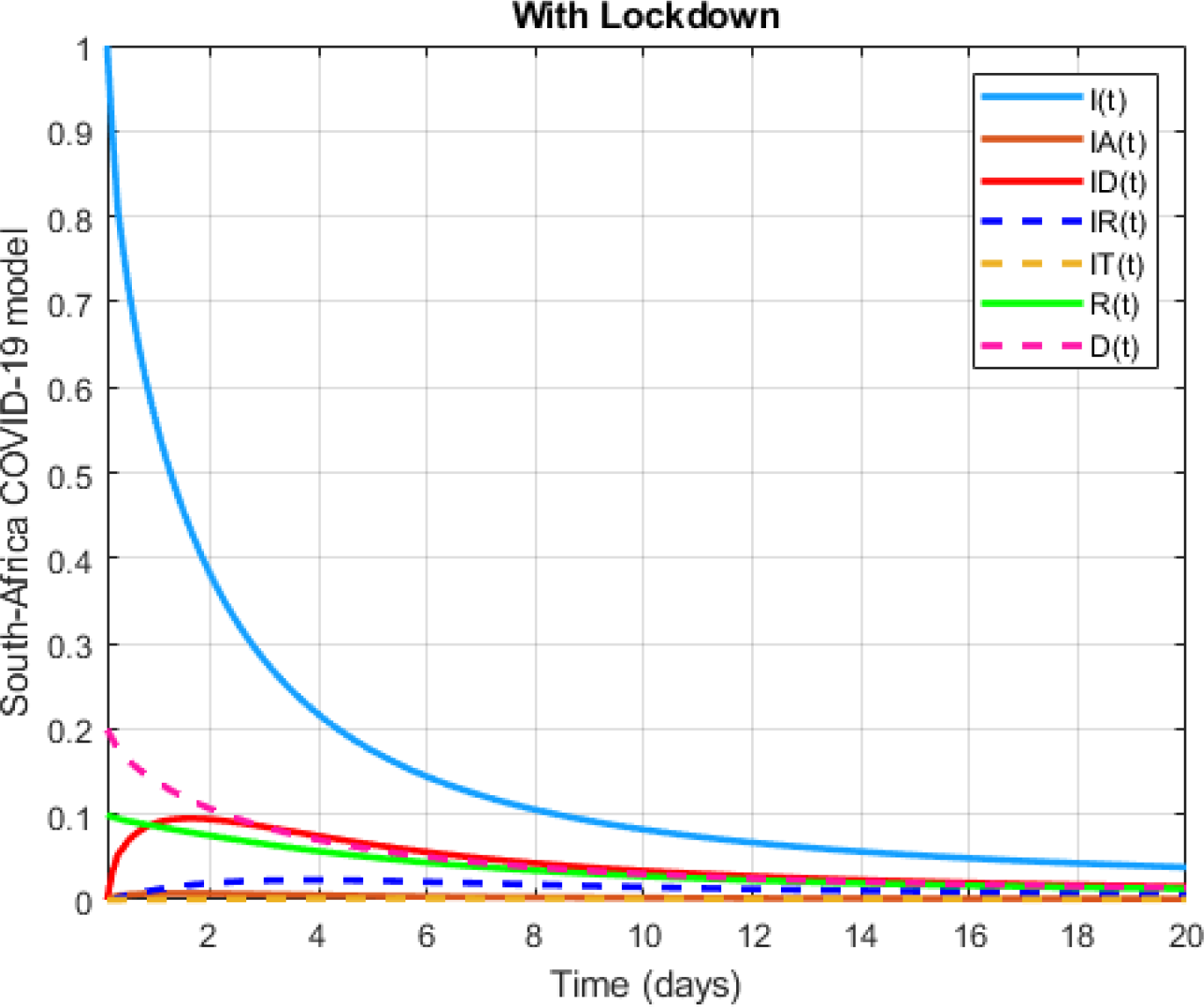
Numerical visualization for COVID-19 model in South Africa for *α* = 0:7; *β* = 0:75:

For same model, initial conditions are chosen

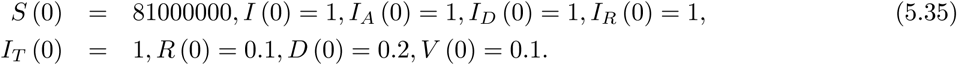

Also the parameters are

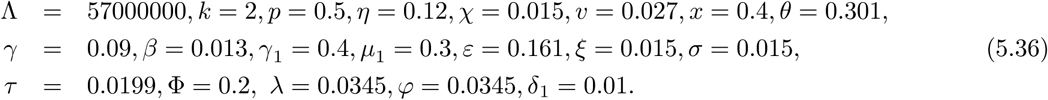

We present numerical simulation for COVID-19 model in Figure 43 and 44.

**Figure 43.**
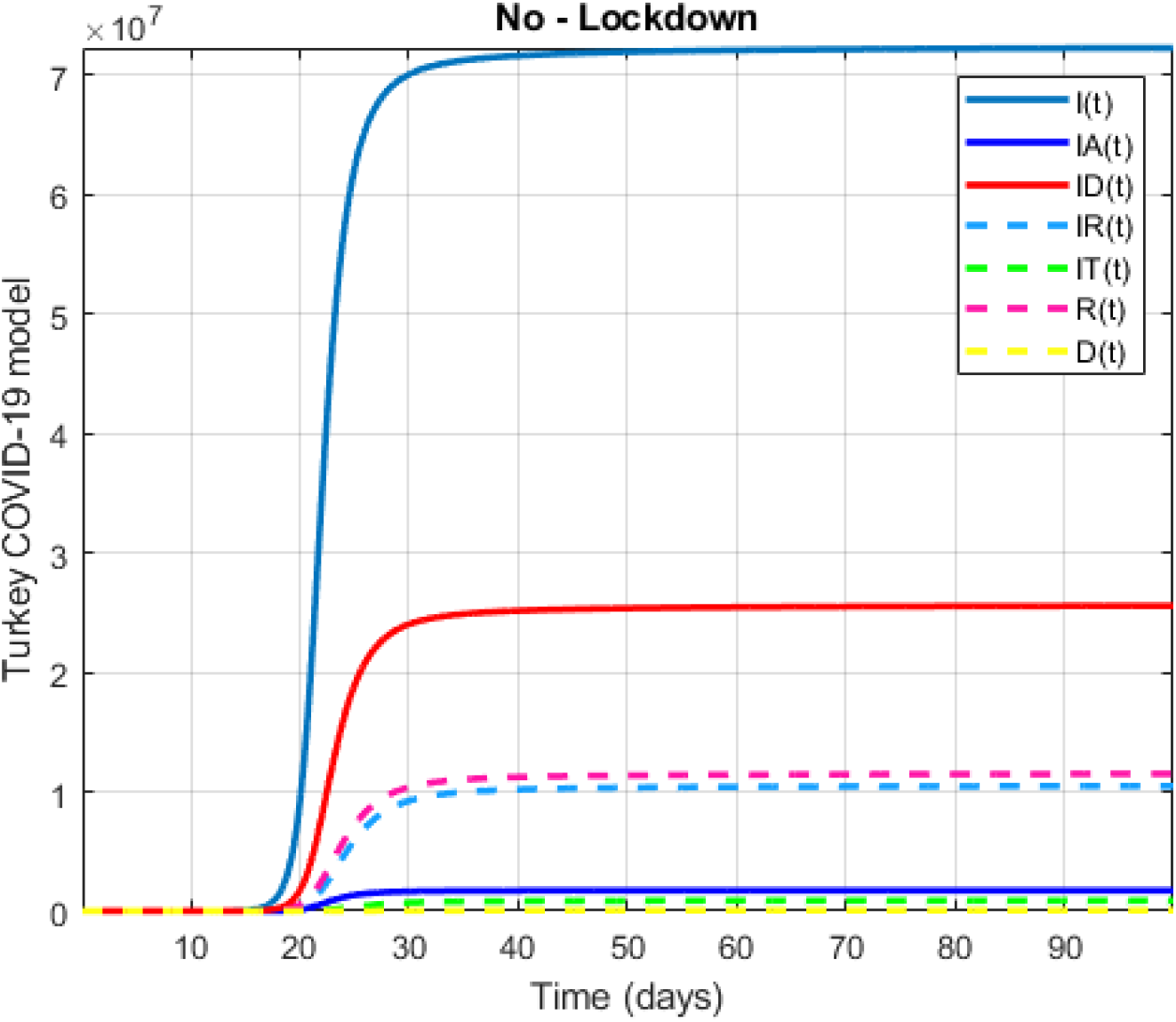
Numerical visualization for COVID-19 model in Turkey for *α* = 0.9, *β* = 0.75.

**Figure 44.**
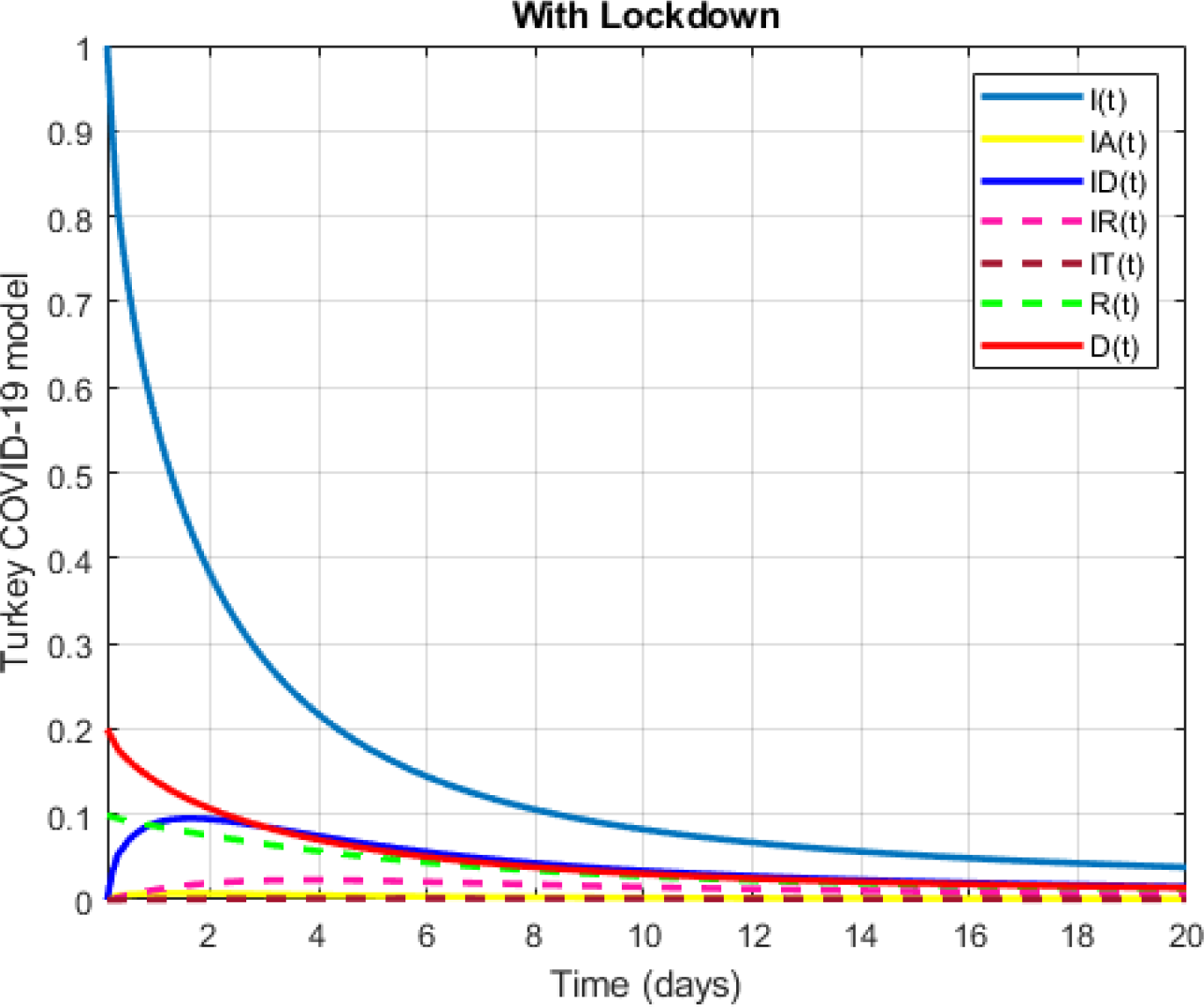
Numerical visualization for COVID-19 model in Turkey for *α* = 0.7, *β* = 0.75.

## 6 Optimal control for COVID-19 model

Optimal control theory provides us important contributions in controlling COVID-19 outbreak. In this section, we will use 7 control variables as 7 possible control strategies to perform our aim. The control variable *u*_1_ is the partial lockdown of schools, travels, universities, some businesses in Turkey. Also government apply partial lockdown by age of people and sometimes states where spread of virus is high. The control variable *u*_2_ is the vaccination which is applied to susceptible individuals. The control variable *u*_3_ is the information campaign to people that have symptoms but not have been tested. The control variable *u*_4_ is the treatment for the infected individuals. The control variable *u*_5_ is the personal protection which is performed with masks, sanitizer and other stuffs.The control variable *u*_6_ is the self-quarantine exposed people. The control variable *u*_7_ is the isolation of infected people.

We modify our model by adding these control variables such as;

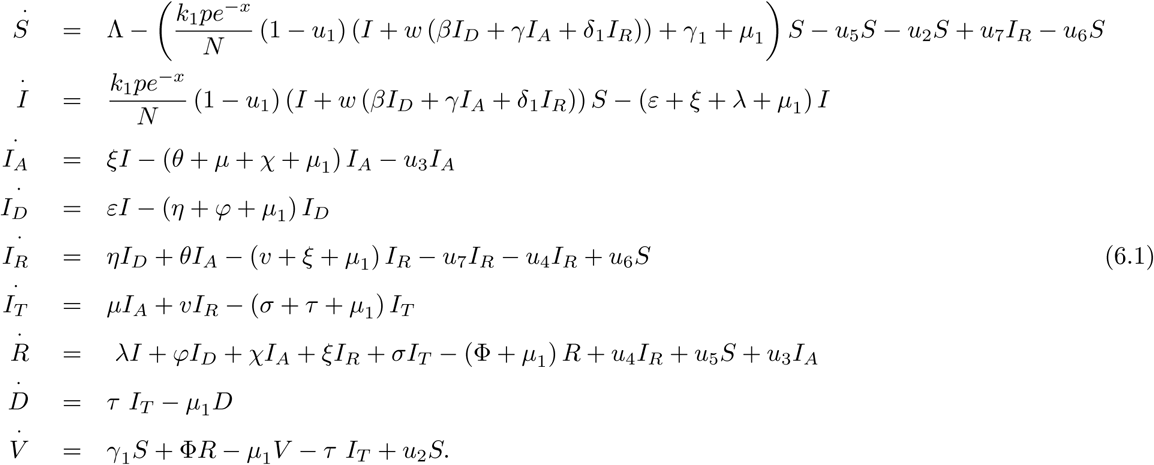

In this paper, we aim to minimize susceptible, infected, critically infected, asymptomatic people and to maximize recovered people while minimizing the costs caused by the partial lockdown, vaccination, information campaign, treatment, personal protection, self-quarantine and isolation. Thus, we construct the cost functional as follows;

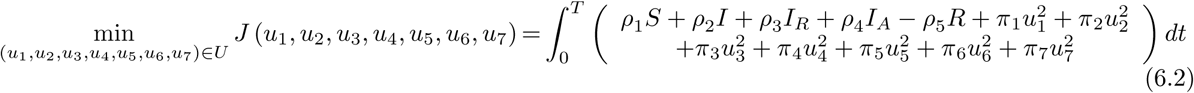

on the set of admissible controls

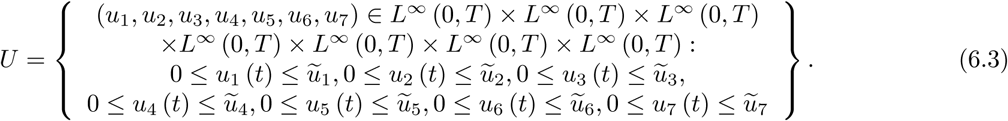

The parameters *ρ*_1_, *ρ*_2_, *ρ*_3_, *ρ*_4_, *ρ*_5_, *π*_1_, *π*_2_, *π*_3_, *π*_4_, *π*_5_, *π*_6_, *π*_7_ represent the weighted parameters.

To show the existence of the optimal control for the problem under consideration, we notice that the set of admissible controls *U* is, by definition, closed and bounded. It is obvious that there is an admissible pair (*u*_1_, *u*_2_, *u*_3_, *u*_4_, *u*_5_, *u*_6_, *u*_7_) for the problem. Hence, the existence of the optimal control comes as a direct result from the Filippove-Cesari theorem [11,12]. We therefore, have the following result:

We prove that the existence of an optimal control of an optimal control is guaranteed by providing the following conditions.

- The set of admissible controls is convex, bounded and closed.
- The set of controls and corresponding state variables is nonempty.
- The right-hand side of the state ODE system is bounded by a linear function in the state and control variables.
- The convexity of the integrand of cost functional with respect to *u* on the set *U*. The Hessian matrix of this functional is given by;

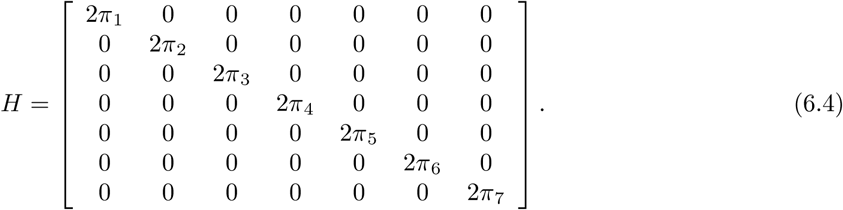

Since the Hessian of of this functional is everywhere positive definite, then the functional *J* (*u*_1_, *u*_2_, *u*_3_, *u*_4_, *u*_5_, *u*_6_, *u*_7_) is strictly convex.

There exist constants *π* = min {*π*_1_, *π*_2_, *π*_3_, *π*_4_, *π*_5_, *π*_6_, *π*_7_} > 0 such that the integrand of the cost functional holds

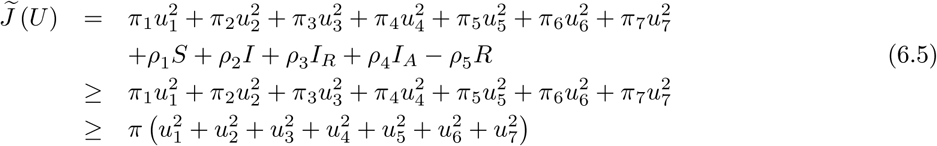

under the condition *ρ*_1_*S* + *ρ*_2_*I* + *ρ*_3_*I_R_* + *ρ*_4_*I_A_* > *ρ*_5_*R*. Applying the Pontryagin’s maximum principle, we present the first order necessary conditions for optimal solution for the considered optimal control problem. To achieve this, we construct the Hamiltonian *H* which is given as

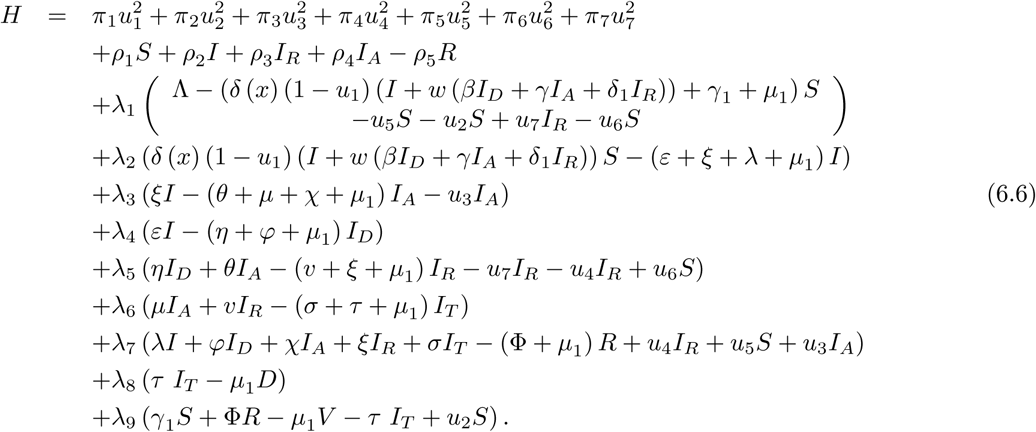

Then there exists λ ∈ ℝ^9^ such that the first order necessary conditions for the existence of optimal control are given by the equations

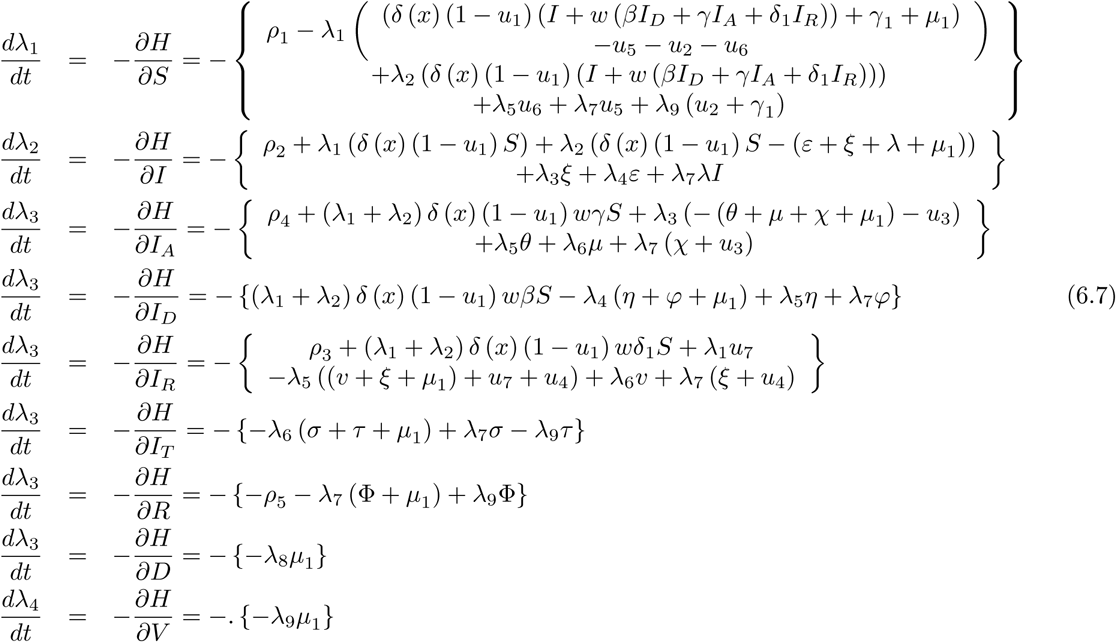

Hence the optimal controls are given as

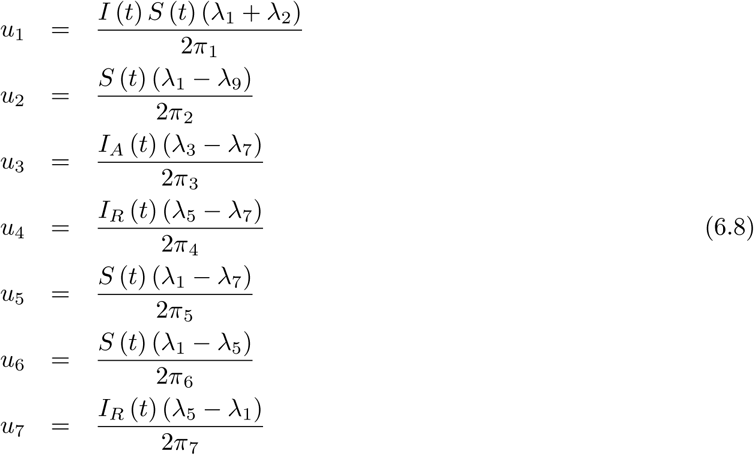

and optimality conditions are given by

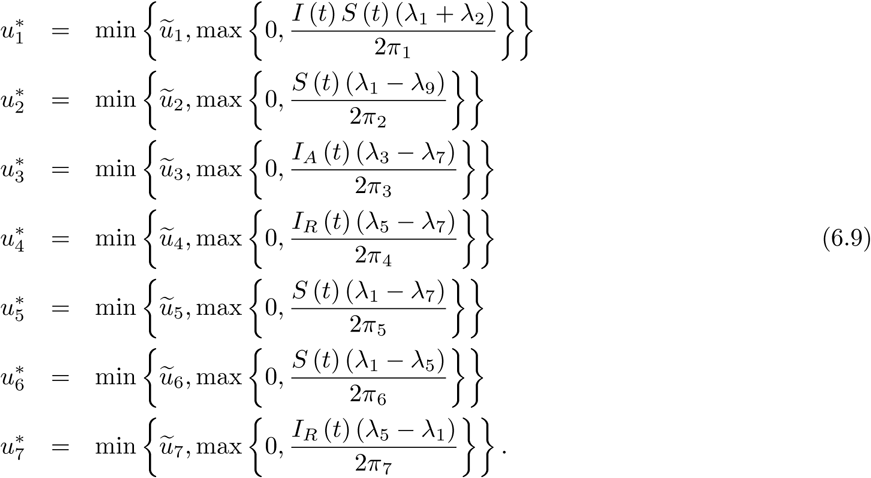

## 7 Discussion, recommendations and conclusion

The COVID-19 fatality on mankind prompted them to undertake serious investigations covering various aspects within several fields of science, technology and engineering in the last 4 months. While researchers have obtained some successful results, they are still struggling to get an effective vaccine that could prevent the spread of the deadly COVID-19 among human beings. From December 2019 to 30 April 2020, there are 3441767 confirmed infected cases, 1097858 recovered and 243922 deaths worldwide. Among which, 6336 confirmed cases, 2549 recovered and 123 deaths are recorded in South Africa; and 124 375 confirmed cases, 58259 recovered and 3336 deaths in Turkey. South Africa registered its first confirmed case of COVID-19 before Turkey on 5 March 2020; while Turkey witnessed its first case on 11 March. The unfolding of the spread of COVID-19 in both countries has defeated the general expectations that South Africa would record more infections and deaths comparing to Turkey. As a result, endless scientific questions were asked within different fields of science; which impelled the compilation of this paper to present critical and comprehensive studies with cases studied in South Africa and Turkey in particular. Although both countries have put in place severe measures to protect their citizens; the statistical predictions from the suggested mathematical models and predictions from statistical analysis show two different patterns for both countries. For instance, in Turkey, a high and exponential growth in new infected numbers from 11 March to 11 April 2020 was observed due to late implementation of the lockdown regulations; however, from 12 April to 02 May 2020 this country has observed an exponential decay in the daily numbers of new infected cases. Thus, Turkey curve seems to follow a lognormal distribution, which of course could mean that they are winning the COVID-19 war; or they took control of the situation. As a result, it is possible that Turkey in the next few months could end the spread of COVID-19, if they maintain the energy and adhere to the measures in place to combat this virus. However, if they relax, the prediction from reliability level method indicated that Turkey could see a very rapid exponential growth in numbers of daily deaths and new infections. Furthermore, it is observed that the exponential decay in the daily number of new infected and death cases corresponds to the period of lockdown implementation and the stringent rules put in place by the Turkish government; by which the contravening of the rule is punishable with a monetary fine. On the other hand in South Africa, although the numbers are not as high as those of Turkey, three phases are observed from statistical results. The first phase goes from 5 March 2020 to 27 March 2020, where the country witnessed an exponential growth in numbers of new infected and deaths daily; and it corresponds to a pre-lockdown period. The second phase began on 28 March until 18 April 2020, in which the country observed a slowly increase of new infected and death daily; a period corresponding to lockdown period enforced with the presence of South Africa Defence Force. While the last phase ranges from 19 April to 02 May 2020, in which the country observed an exponential growth in numbers of new infected and deaths per day. This exponential growth is attributed to the relaxation and disobedience of lockdown regulations; probably due to economic breakdown, increasing poverty effects among the larger population and also due to migration from level five to level four on the 1 st of May 2020. Therefore, as the provision of a suitable vaccine to save and protect human beings the wrath and the fatality of the deadly COVID-19, which breakout in Wuhan, China, December 2019 is delayed; it is clearly evident that the exponential growth in numbers of new infected can only be stopped or halted by enforcing the implementation of social distancing and ensuring that people do frequently wash their hands upon touching any object or even animals; whether infected with COVID-19 or not. Additionally, the wearing of masks should be adopted in public places to avoid the spreading of the virus, in case the social distancing rules is not being kept. It is further paramount that the medical workers in charge of COVID-19 patients are well protected to minimize the contraction of the virus from the patients and passing it on to the general public. In addition to the prohibition of alcohol sales and usage in public places; public smoking should also be prohibited in the effort to combat the spread of COVID-19. Moreover, the statistical analysis results specifically the reliability level prediction and the results obtained from suggested mathematical models; indicated that without social distancing restrictions or clear implementation of lockdown regulations, it will be impossible for countries to control the spread of COVID-19. This implies that the number of new infected and deaths per day would be difficult to contain, resulting in the fight against the virus to get out of hand. These outcomes from reliability level are therefore indicated in blue lines in Figures 18, 19, 20, 29 and 30 those from reliability level. The suggested mathematical models with different differential operators including classical and nonlocal operators in the last 12 Figures as case with lockdown and no-lockdown presented for different fractional orders also confirmed the results obtained from the reliability level. In consideration of all prediction results, it is concluded that South Africa has not yet won the war against COVID-19 and serious outbreak are expected in the near future as the climate season changes to winter. Cold seasons are scientifically proven to be thriving climate for the survival of corona virus. Therefore to avoid this foreseen crisis, social distancing must be a responsibility of each person living within the Republic of South Africa, and the transition from level 5 to level 1 should be implemented very wisely.

## Data Availability

https://en.wikipedia.org/wiki/COVID-19_pandemic_in_South_Africa

## Acknowledgment

The authors of this paper will like to thank Dr. M. Altaf Khan and Prof. Dr. Zakia Hammouch for their valuable comments to enhance the stability part of our paper.

## References

[1] Giordano, G., Blanchini, F., Bruno, R. et al. Modelling the COVID-19 epidemic and implementation of population-wide interventions in Italy. Nat Med (2020).

[2] MA Khan, A Atangana, Modeling the dynamics of novel coronavirus (2019-nCov) with fractional derivative, Alexandria Engineering Journal, 2020.

[3] KM Owolabi, A Atangana, Mathematical analysis and computational experiments for an epidemic system with nonlocal and nonsingular derivative, Chaos, Solitons & Fractals 126, 2019, 41–49.

[4] Tuite AR., Fisman DN., Greer LA., Mathematical modelling of COVID-19 transmission and mitigation strategies in the population of Ontario, Canada, CMAJ, 2020.

[5] Anastassopoulou C., Russo L., Tsakris A., Siettos C., Data-Based Analysis, Modelling and Forecasting of the COVID-19 outbreak, 2020, medRxiv.

[6] Kucharski AJ., Russell TW., Diamond C., Liu Y., Edmunds J., Funk S., Eggo RM., Early dynamics of transmission and control of COVID-19: a mathematical modelling study, Lancet Infect Dis. 2020.

[7] COVID-19 pandemic in South Africa, https://en.wikipedia.org/wiki/COVID-19_pandemic_in_South_Africa, 2020.

[8] LaSalle JP., The stability of dynamical systems, SIAM Press, 1976.

[9] Driessche P., Watmough J., (2002), Reproduction numbers and sub-threshold endemic equilibria for compartmental models of disease transmission, Mathematical Biosciences, 180 (1), 29–48.

[10] Atangana A, Araz S·I., New numerical method for ordinary differential equations: Newton polynomial, Journal of Computational and Applied Mathematics 372, 2020, 112622.

[11] S. Nababan; A. ippov-type lemma for functions involving delays and its application to time delayed optimal control problems, Optim. Theory appl. 27 3 (1979), 357–376.

[12] Alvarez FE., Argente D., Lippi F., A Simple Planning Problem for COVID-19 Lockdown, National Bureau of Economic Research, 2020002E

